# Delayed bystander CD8 T cell activation, early immune pathology and persistent dysregulation characterise severe COVID-19

**DOI:** 10.1101/2021.01.11.20248765

**Authors:** Laura Bergamaschi, Federica Mescia, Lorinda Turner, Aimee Hanson, Prasanti Kotagiri, Benjamin J. Dunmore, Hélène Ruffieux, Aloka De Sa, Oisín Huhn, Michael D Morgan, Pehuen Pereyra Gerber, Mark R. Wills, Stephen Baker, Fernando J Calero-Nieto, Rainer Doffinger, Gordon Dougan, Anne Elmer, Ian G Goodfellow, Ravindra K. Gupta, Myra Hosmillo, Kelvin Hunter, Nathalie Kingston, Paul J. Lehner, Nicholas J. Matheson, Jeremy K. Nicholson, Anna M. Petrunkina, Sylvia Richardson, Caroline Saunders, James E.D. Thaventhiran, Erik J. M. Toonen, Michael P. Weekes, Cambridge Institute of Therapeutic Immunology and Infectious Disease-National Institute of Health Research (CITIID-NIHR) COVID BioResource Collaboration, Berthold Göttgens, Mark Toshner, Christoph Hess, John R. Bradley, Paul A. Lyons, Kenneth G.C. Smith

## Abstract

In a study of 207 SARS-CoV2-infected individuals with a range of severities followed over 12 weeks from symptom onset, we demonstrate that an early robust bystander CD8 T cell immune response, without systemic inflammation, is characteristic of asymptomatic or mild disease. Those presenting to hospital had delayed bystander responses and systemic inflammation already evident at around symptom onset. Such early evidence of inflammation suggests immunopathology may be inevitable in some individuals, or that preventative intervention might be needed before symptom onset. Viral load does not correlate with the development of this pathological response, but does with its subsequent severity. Immune recovery is complex, with profound persistent cellular abnormalities correlating with a change in the nature of the inflammatory response, where signatures characteristic of increased oxidative phosphorylation and reactive-oxygen species-associated inflammation replace those driven by TNF and IL-6. These late immunometabolic inflammatory changes and unresolved immune defects may have clinical implications.

## Introduction

The immune pathology associated with COVID-19 is complex (Wang et al., 2020; Zhou et al., 2020). Most infected individuals mount a successful anti-viral response, resulting in few if any symptoms. In a minority of patients there is evidence that ongoing cytokine production develops, associated with persistent systemic inflammation, end-organ damage and often death (Del Valle et al., 2020; Lucas et al., 2020). The relationship between the initial immune response to SARS-CoV-2, viral clearance, and development of the ongoing inflammatory disease which drives severe COVID-19 is not clearly established, nor have the kinetics of the immune changes seen in COVID-19 been fully assessed as disease progresses. Defective immune recovery might drive ongoing disease, and contribute to long-term disease sequelae (“long COVID”) and perhaps contribute to secondary immunodeficiency with an increased risk of subsequent infection.

Severe COVID-19 is associated with profound abnormalities in circulating immune cell subsets (Arunachalam et al., 2020; Hadjadj et al., 2020; Kuri-Cervantes et al., 2020; Laing et al., 2020; Mann et al., 2020; Mathew et al., 2020; Maucourant et al., 2020; Schulte-Schrepping et al., 2020; Su et al., 2020; Wen et al., 2020). There is a decrease in many peripheral blood subsets of both CD4 and CD8 T cells (Kuri-Cervantes et al., 2020; Laing et al., 2020; Mann et al., 2020; Mathew et al., 2020; Su et al., 2020), but an increase in activated and differentiated effector cells (Arunachalam et al., 2020; Hadjadj et al., 2020; Kuri-Cervantes et al., 2020; Laing et al., 2020; Mann et al., 2020; Mathew et al., 2020; Su et al., 2020). Cells expressing PD1 and other inhibitory molecules are increased, though whether these reflect genuine T cell exhaustion or changes accompanying T cell activation, has not been firmly established (Hadjadj et al., 2020; Laing et al., 2020; Mathew et al., 2020; Su et al., 2020; Zheng et al., 2020). There is, nonetheless, evidence of functional impairment in both CD8 and CD4 T cells in a number of studies (Chen and Wherry, 2020). Data on T helper cell subsets is variable, but there is evidence of increased T_H_17 cells and markedly reduced T follicular helper cells (T_FH_) (Chen and Wherry, 2020; Mathew et al., 2020; Su et al., 2020). There have been conflicting reports regarding B cell immunity (Laing et al., 2020; Mann et al., 2020; Mathew et al., 2020), but increased circulating plasmablasts (Arunachalam et al., 2020; Hadjadj et al., 2020; Laing et al., 2020; Mathew et al., 2020) and reduced germinal centre responses (Su et al., 2020) are consistently observed in severe COVID-19. A number of innate T cell subsets, including γδ T cells and MAIT cells, are also reduced (Kuri-Cervantes et al., 2020; Laing et al., 2020; Maucourant et al., 2020; Parrot et al., 2020), as are non-classical monocytes (Schulte-Schrepping et al., 2020; Su et al., 2020) and both plasmacytoid and myeloid dendritic cells (Kuri-Cervantes et al., 2020; Laing et al., 2020)

By analysing longitudinal samples from COVID-19 patients with a range of disease severities, for up to 3 months from symptom onset, we were able to address two important questions regarding the immune response to SARS-CoV-2: (i) How does the very early immune response in patients who cleared virus and recovered from disease with few or no symptoms, compare with those who progressed to severe inflammatory disease. This provided insight into what constitutes an effective versus an ineffective immune response, and whether systemic inflammation is an early or later development in those who progress to severe disease. (ii) How rapidly do the profound immune defects that accompany severe COVID-19 recover, and do the kinetics of recovery relate to ongoing inflammation and clinical status.

We recruited 207 patients with COVID-19, ranging from asymptomatic healthcare workers in whom SARS-CoV-2 was detected on routine screening, through to patients requiring assisted ventilation, and compared their results to 45 healthy controls. We performed detailed immune phenotyping at multiple time points up to 90 days from symptom onset, reporting absolute cell counts rather than proportions as the latter are difficult to interpret in many studies in the context of profound lymphopenia. Correlation of these data with clinical and other meta-data demonstrated that the immune response in patients with progressive COVID-19 is delayed compared to those with mild disease, and is inflammatory in nature from the outset. Early immune cellular changes predict severe disease course. Their variable recovery over 3 months is associated with marked changes in the nature of the systemic inflammation seen in severe COVID-19.

## Results

### Patient cohorts

SARS-CoV-2 PCR positive subjects were recruited for this study between 31^st^ March and 20^th^ July 2020 in three contexts. Those without symptoms, or with mild symptoms, were recruited from routine screening of healthcare workers (HCW) at Addenbrooke’s Hospital (Rivett et al., 2020). COVID-19 patients were recruited at presentation to Addenbrooke’s hospital if their symptoms were consistent with COVID-19, and then formally included for follow-up if subsequent swab results were positive for SARS-CoV-2. In addition, some patients were recruited having already been admitted, twenty-nine having developed COVID-19 in hospital after admission for another reason, with others recruited after transfer to intensive care at either Addenbrooke’s or Royal Papworth hospitals. After recruitment patients were bled approximately weekly, and then invited to an outpatient follow-up visit 4-12 weeks after study enrolment. HCWs were sampled at study entry, and then approximately 2 and 4 weeks later.

Study participants were divided into five categories according to clinical severity, which we use throughout this paper unless otherwise stated (**Figures 1A and Item S1**). These were:

**Figure 1:**
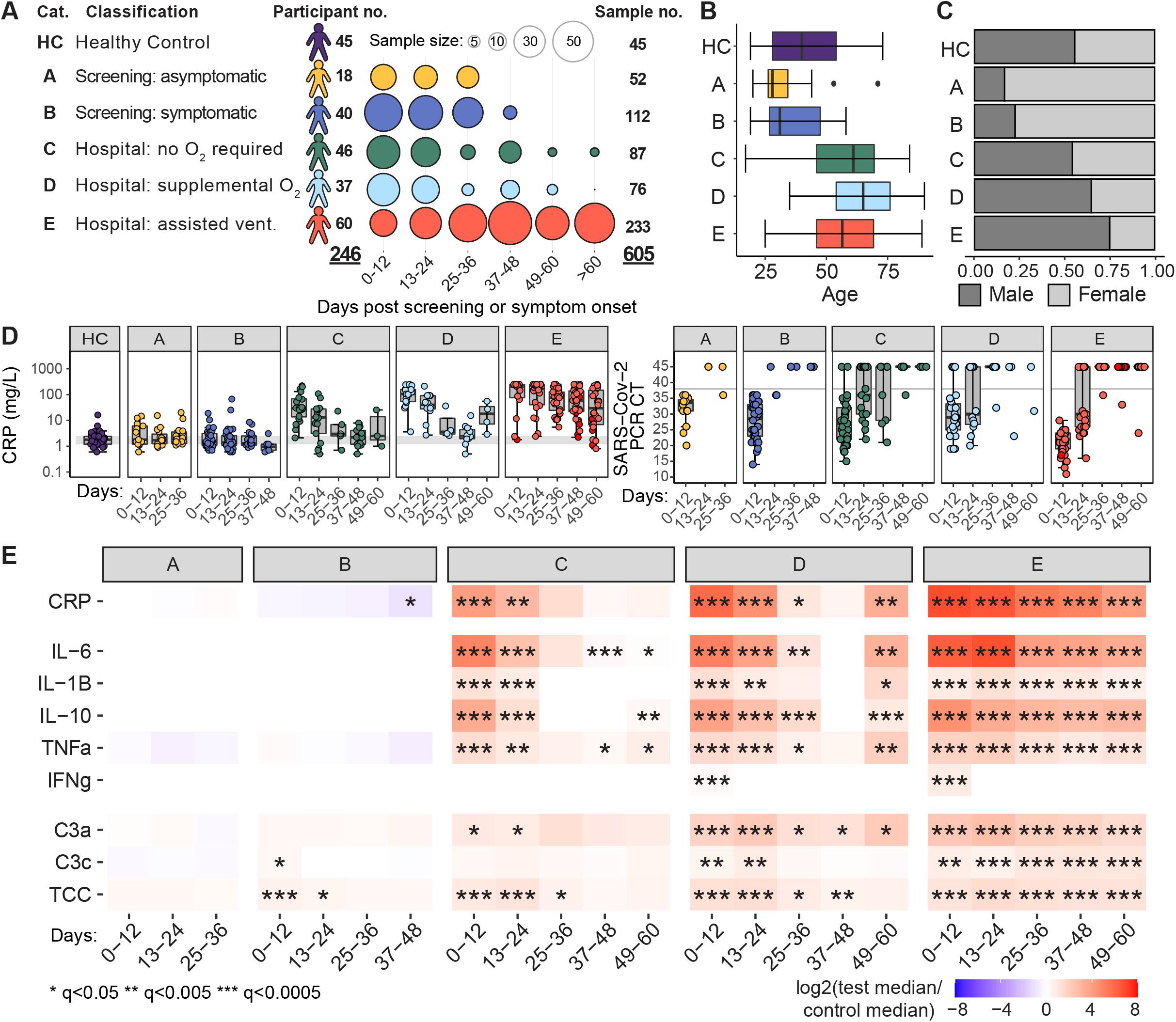
Cohort characteristics. **A)** Study participant and sample numbers split by severity categories and 12-day time bins post screening (group A) or symptom onset (group B-E). Distribution of participant age **B)** and gender **C)** across severity categories. **D)** Boxplots showing measured CRP (mg/L) and SARS-CoV-2 PCR cycle threshold for samples collected within 12-day time bins. Grey band indicates the interquartile range of the corresponding measure in HCs, or the SARS-CoV-2 negative swab cycle threshold (CT > 38). **E)** Heatmap showing log_2_ fold change in median CRP and serum cytokine and complement measures between COVID-19 cases and HC, within severity categories and across 12-day time bins. Wilcoxon test FDR adjusted p-value: *<0.05, **<0.005, ***<0.0005. CT, cycle threshold; CRP, C-reactive protein; IL, interleukin; TNF, tumour necrosis factor; IFN, interferon; C3, complement 3; TCC, terminal complement complex.

- A) asymptomatic HCWs.
- B) HCWs who either were still working with mild symptoms insufficient to meet the criteria for self-isolation (Rivett et al., 2020), or who were symptomatic and self-isolating.
- C) patients who presented to hospital but never required oxygen supplementation.
- D) patients who were admitted to hospital and whose maximal respiratory support was supplemental oxygen.
- E) patients who at some point required assisted ventilation. Three patients who died without admission to intensive care were also included in this severe group.

In addition, 45 healthy controls were recruited, distributed across a range of age and gender.

This study analysed the immune phenotype of 605 blood samples from 246 participants out to 90 days from the onset of symptoms (**Figure 1A**). Patients were included irrespective of co-morbidity to reflect “real world” disease (**Table S1**), apart from the exclusion of 6 patients whose severe non-COVID-19 disease determined their clinical outcome, which made investigation results uninterpretable, and in whom COVID-19 was essentially a side-issue (details in **Table S2**). It is important to note that as the clinical severity category increased, patients were more likely to be older and to be male (**Figures 1B and 1C**), as expected (Wang et al., 2020). A high-sensitivity C-reactive protein (CRP) assay was performed on all samples, and is shown for each cohort - demonstrating that classifying disease severity by subdivision on the basis of maximal respiratory support is reflected in the CRP (**Figure 1D**). Patient courses are measured in time since symptom onset for groups B through to E. As they are asymptomatic, those in group A are measured from the date of their first positive swab. This means that they are likely to have been sampled, on average, later post-infection than patients in the other groups, and are therefore not directly comparable to groups B-E in terms of time course. This needs to be kept in mind when comparing group A to the other groups throughout this study. CRP (and, later, other variables such as cell number) is compared to the interquartile range of 45 healthy controls. Nasopharyngeal swabs were assessed for SARS-CoV-2, allowing diagnosis and inclusion in this study, and were repeated in some patients at various times. Initial viral titres (higher in those with low PCR cycle threshold or CT values) were higher in group E. With only occasional exceptions, patients in all severity groups had cleared virus by 24 days after the onset of symptoms (**Figures 1D and S1A**). Of the 6 patients with positive swabs after 30 days, four were overtly immunosuppressed (3 solid organ transplants with recent induction/rejection treatment, 1 myeloma on B-cell depletion therapy) and one was a peritoneal dialysis patient admitted with peritonitis.

We will first outline the major datasets collected in this study, before integrating them to study early and recovering disease.

### Cytokines and complement components

We assessed cytokine and complement components in plasma at each time point (**Figures 1E and S1B**). Asymptomatic HCWs in group A had no evidence of cytokine or complement dysregulation, while those with mild symptoms (group B) showed an early, transient increase in C3c and the terminal complement complex (TCC), but not in C-reactive protein (CRP) or cytokine levels. Once patients developed symptoms severe enough to warrant attendance at hospital (group C or above), a different picture was apparent. Both IL-6 and TNF-α were significantly raised, along with other cytokines, as were all of the complement components measured. These abnormalities were maximal at the first bleed, and largely persisted in group E, where many patients remained in intensive care throughout their course. Abnormalities in both IL-6 and TNF-α persisted in groups C and D despite clinical improvement (all had been discharged by the 49-60 time window). Interferon-gamma (IFN-γ) was raised in only a subset of patients, and in all severity groups this increase was short-lived. Increased C3c was prominent in those with mild symptoms (group B), while C3a became the dominant complement component elevated in more severe disease (groups C –E). Cytokine levels vary with disease severity and over time, with IL-6, TNF-α, IL-10 and IL-1β rising in those with more severe disease (groups C-E). In contrast, there is no evidence of increased inflammatory cytokines in patients from groups A and B, pointing to marked differences in very early immune responses between resolving and progressive disease. In addition, the persistence of cytokine abnormalities even beyond 60 days from symptom onset could have implications for resolution of both immune abnormalities and clinical disease.

### Both onset and recovery of immune cellular abnormalities vary with disease severity

Using standardised flow cytometry panels, we explored the size of 64 cell populations over time across the five clinical strata. Trucount analysis enabled calculation of absolute cell numbers. Cellular changes were assessed across time “bins” of 12 days (using the earliest measure per patient per bin in instances of repeat sampling), with two cell subsets shown in **Figure 2A** as examples. The outcomes for 30 cell types are summarised in a heat map, showing changes in cell population size relative to the median for healthy controls (**Figure 2B**). CyTOF, which uses whole blood rather than peripheral blood mononuclear cells (PBMCs), was also used in a subset of patients, as this allows quantification of granulocytes (largely absent in PBMCs) and non-classical and intermediate monocytes (both variably lost in PBMC separation: **Figure 2B** and Methods). Apart from these exceptions, cell numbers generated by CyTOF correlated well with data from flow cytometry (**Figure S2**). Data for all cell types, also including time as a continuous variable, is shown in **Item S2**. Also shown in **Item S2** are samples taken beyond 48 days which, with the exception of group E, were not numerous enough for statistically useful inclusion in the “binned” data.

**Figure 2:**
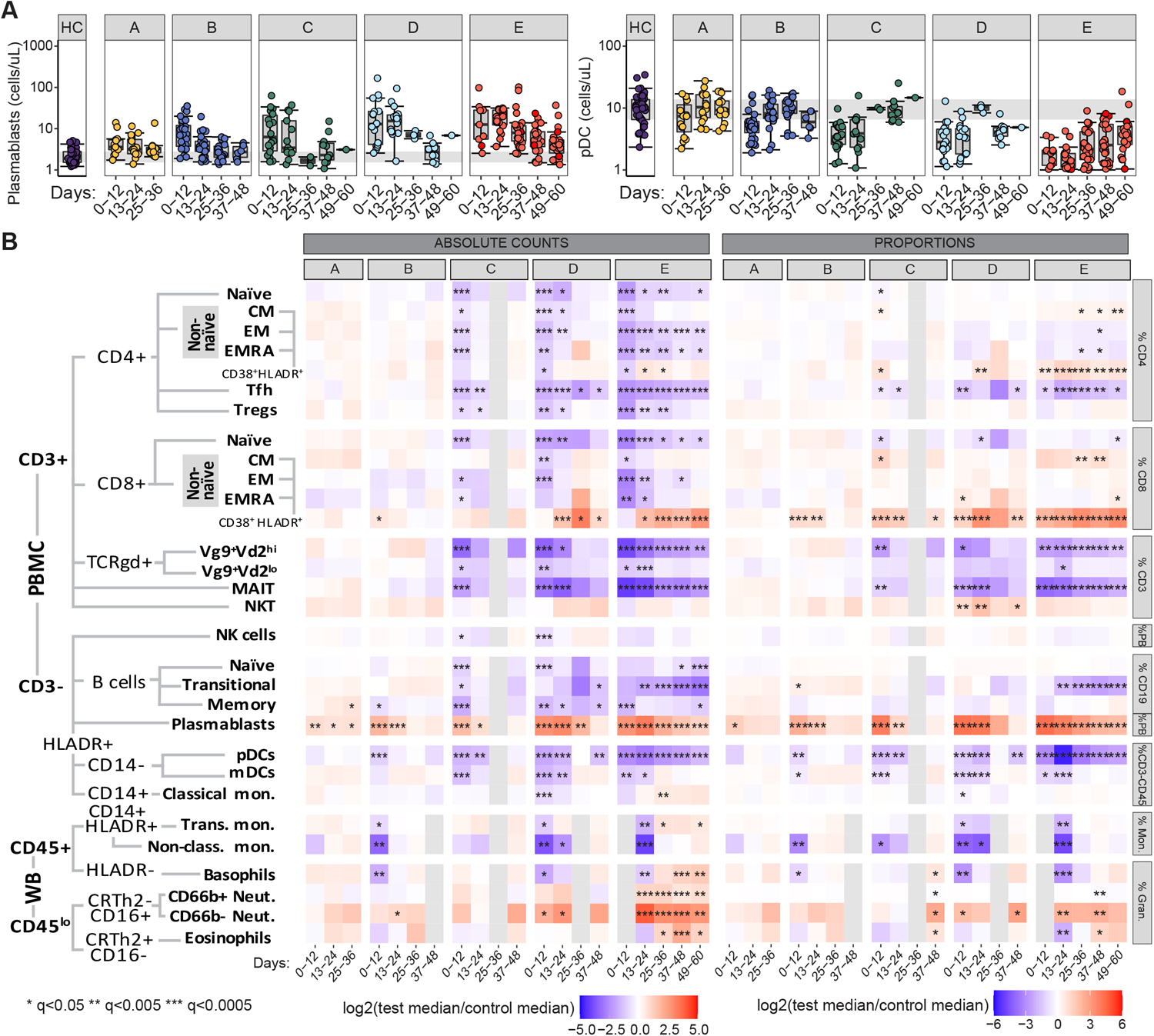
Cellular changes over time. **A)** Boxplots showing absolute counts (cells/uL) for two representative cell populations, split by severity categories and 12-day time bins post screening (group A) or symptom onset (group B-E). Grey band indicates the interquartile range of the corresponding population in HC. **B)** Heatmap showing the log_2_ fold change in median absolute cell count (left), or proportion of major subset (right), between COVID-19 cases and HCs, within severity categories and across 12-day time bins. Wilcoxon test FDR adjusted p-value: *<0.05, **<0.005, ***<0.0005. Population hierarchy is shown to the left. Population proportions are calculated within parent populations listed to the right. PB or PBMC, peripheral blood mononuclear cells (analysed by flow cytometry); WB, whole blood (analysed by CyTOF).

Few changes in the immune phenotypes were seen in patients with asymptomatic (A) and very mild (B) disease, but once symptoms had become sufficient to warrant presentation to hospital, the picture changed markedly (see below). There were widespread immune abnormalities in patients with moderate through to severe disease (C-E), most marked at the time of first blood sampling (**Figure 2B**), even when this coincided with or was within a day or two of symptom onset (**Item S2**). Almost all CD4 T cells subsets were reduced, as were many CD8 T cell subsets and both naive and memory B cells – in contrast plasmablast numbers in the blood rose in all groups (see below). A number of innate lymphoid subsets were also reduced, including MAIT cells, various γδ T cell subsets, and NK cells. The myeloid compartment was also affected, with a reduction in myeloid dendritic cells, and both non-classical and intermediate monocytes. These changes were correlated with, and were predictive of, severity, as discussed below.

We have also calculated leucocyte number as a proportion of “parent” populations, either of total PBMC (**Figure S3A**) or of major lymphoid compartments (**Figure 2B and S3A**), as this allows comparison with most published literature (including all single cell studies). Considering proportion allows the relative impact on subsets to be compared, but underestimates the immune pathology associated with COVID-19, for example missing the severity-correlated reduction seen in many lymphocyte subsets early after symptom onset, and the persistent low numbers of most T cell subsets seen in more severe disease. In agreement with this, analysis of CyTOF and CITE-seq data, which also examine differences in cell proportions, did not identify any alterations in immune cell populations between disease severity groups that were not been observed in the flow cytometry data (**Figure S3B and C**).

### Blood transcriptomic inflammation-related signatures vary with severity and time

RNA was prepared from whole blood collected in PAXGene tubes at each bleed. RNA was isolated, and whole blood transcriptomes were generated by RNA-Sequencing (see Methods) and analysed in two time “bins” – 0 to 24 days and 25 to 48 days (finer gradations were not possible due to sample size). We first analysed the transcriptome data using PLIER which performs matrix factorization to identify interpretable latent factors. The contribution to each latent factor by immune cell subsets was then calculated across the severity groups and time points (**Figure S4A and B**). These RNA-Seq-derived latent factors were broadly aligned with the pattern observed in the cell count data (**Figure 2B**). An exception to this was the pronounced neutrophil signature seen at day 0 to 24 across groups C to E, and persisting at day 25-48 in group E. This transcriptomic analysis shows more pronounced neutrophil dysregulation across severity categories than is suggested by increasing neutrophil number alone. An erythrocyte gene expression-driven latent factor was also seen, and was prominent in group E at late times. This may be associated with heme metabolism, and is discussed below.

We then used weighted gene correlation network analysis (WGCNA) to identify, in an unbiased fashion, modules of co-regulated genes in the whole blood transcriptome data, where each module can be summarised as an “eigengene”. Prominent gene expression modules were observed, that correlated with both disease severity and time (**Figures 3A and B, S4C, S4D and Table S3**). It can be seen that the module enriched for TNF-α /IL-6 genes correlates well with the cytokine levels determined in Figure 1 – rising early in groups C-E and then largely resolving by 25-48 days. A neutrophil activation module was also prominent early across groups C to E, as was one associated with glycolysis. Thus, there is clear transcriptional evidence of activation of broad inflammatory pathways at early time points, and these largely recover in most patient groups (with the exception of group E, in which many patients have persistent disease). In contrast, an interferon-related module is upregulated prominently in groups B-E at day 0 to 24 from symptom onset, but declines at later time points (**Figures 3A)**. As previously described (Banchereau et al., 2017), the relative contributions to this module by Type I, II and III interferons cannot be easily distinguished at the transcriptome level. A more detailed analysis of the kinetics of this interferon-stimulated gene (ISG)– associated module shows that, while expression peaks at different levels in each severity group, it then declines in all of them by around 30 days (**Figure 3C**), coincident with viral clearance and occurring irrespective of clinical and inflammatory state (**Figure 3D**).

**Figure 3:**
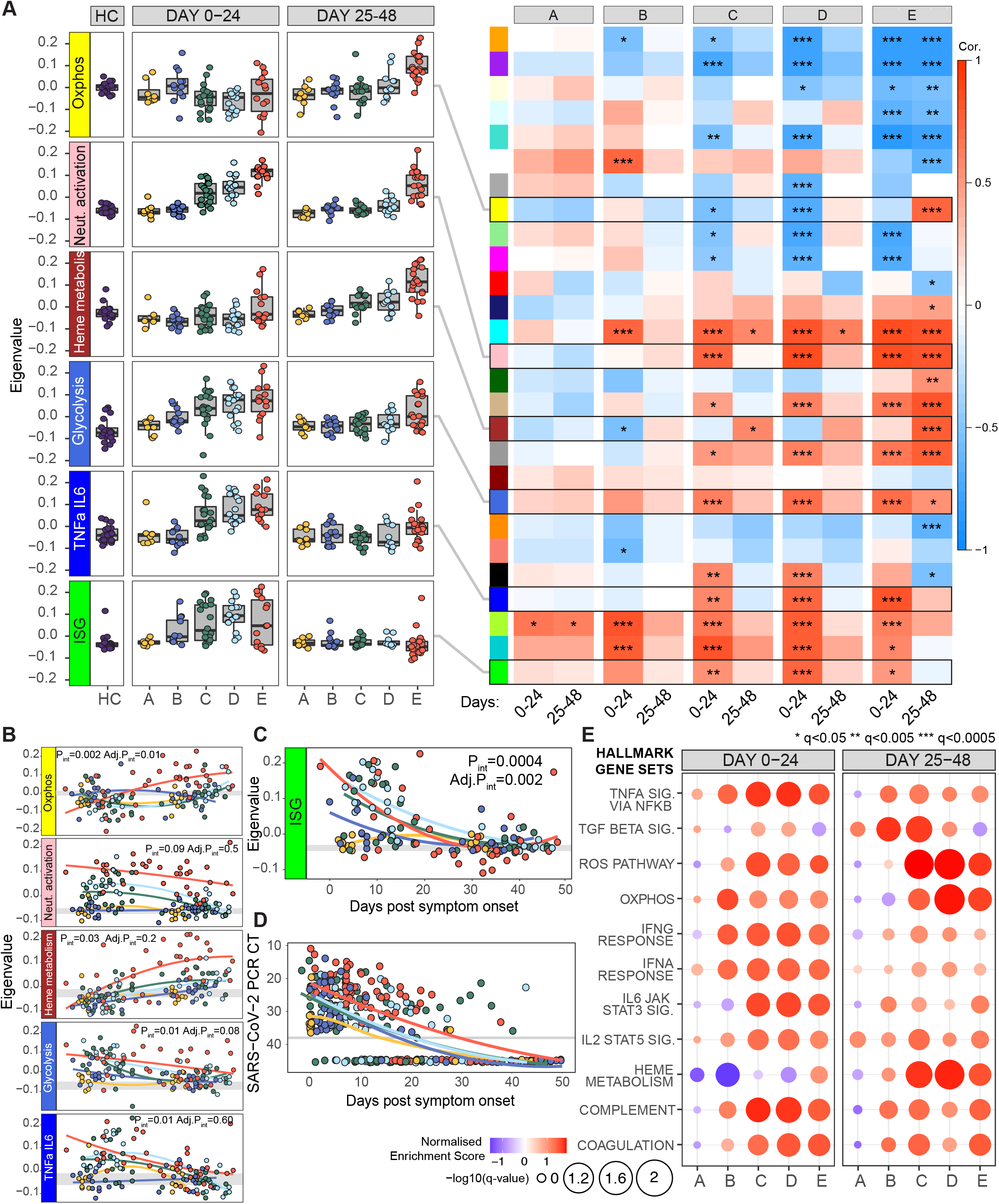
Whole blood transcriptomic signatures over time. **A)** Heatmap derived from WGCNA, illustrating the correlation of whole blood co-expression gene modules (coloured blocks, y axis) with COVID-19 severity groups (x axis) split by 24-day time bins post screening (group A) or symptom onset (group B-E). Boxes are coloured by strength of correlation. Boxplots displaying underlying sample eigenvalues within key transcriptomic modules, according to disease severity and time. **B)** Mixed-effects model with quadratic time trend showing the longitudinal expression of key eigengenes over time, grouped by severity. Grey band indicates the interquartile range of the corresponding eigengene in HCs. Nominal and adjusted p-values for the time x severity group interaction term are reported. **C)** Mixed-effects model showing longitudinal expression of eigengene capturing interferon stimulated genes (ISG), and **D)** equivalent mixed-model showing changes in SARS-CoV-2 PCR cycle threshold (viral load), over time and across severity groups. Y-axis inverted in D. **E)** GSEA assessing enrichment for HALLMARK genesets against HC in COVID-19 cases split by severity categories and 24-day time bins post screening (group A) or symptom onset (group B-E). FDR adjusted p-value is shown by circle diameter, with colour representing normalised enrichment score of the associated gene set. CT, cycle threshold; ISG, interferon stimulated genes; ROS, reactive oxygen species.

Finally, a supervised gene set enrichment analysis (GSEA) was performed using publicly available Hallmark gene signatures (**Figure 3E**) (Liberzon et al., 2015). These findings were largely consistent with those generated from the unbiased approaches above, and demonstrated a late upregulation of genes associated with reactive oxygen species and oxidative phosphorylation. Late upregulation of these pathways is discussed below in the context of immune recovery.

### Immune phenotype at presentation correlates with severity and may predict outcome

To determine if the immune phenotype at presentation correlated with, or indeed could predict, subsequent disease course, we first performed a Principal Component Analysis using cell numbers across 24 primary immune cell populations from blood draws taken between 0 and 10 days after the development of symptoms. Study participants in groups A and B clustered together with HC and were separate from those in groups C-E (**Figure S5A**). Hierarchical clustering of absolute cell counts from COVID-19 cases identified two clear clusters (**Figure 4A**), one almost entirely comprised of HCWs from groups A and B (cluster 2), and the other containing all patients who progressed to ventilation and/or death, and most who required supplementary oxygen support (cluster 1). Clustering patients using RNA-Seq data obtained from 1-10 days after onset largely recapitulated that seen using immune cell number, and was driven by ISG, TNF-α and IL-6 associated gene pathways (**Figure S5B**). The severe cluster 1 was associated with increased age, CRP, TNF-α and IL-6 (**Figures 4B and ItemS3A**). Early differences between cell types drive this clustering, despite different cell subsets having quite different subsequent trajectories (**Item S3B**). To determine which specific immune subsets underpinned this clustering, we used a sparse partial least squares discriminant analysis (sPLS-DA), which showed patient clusters 1 and 2 could be discriminated with a minimum classification error rate of 0.07 ± 0.02 (93% accuracy) based on 13 key cell populations selected by the model as most informative for cluster prediction (**Figures 4C, S5C-E**). The area under the receiver operator characteristic curve (AUROC) for patient cluster classification based on these 13 cell types was 0.98 (98% chance of accurate cluster prediction) (**Figure S5F)**. CRP alone was inferior to the cell types in classifying patients, and nor did it improve their performance when added (**Figure S5G**). These cell types were often the most profoundly affected by severe COVID-19, and those most associated tended to recover poorly over time (particularly MAIT, γδ T cells, T_FH_ and CD4 EMRA T cells) suggesting a persistent association with disease severity.

**Figure 4:**
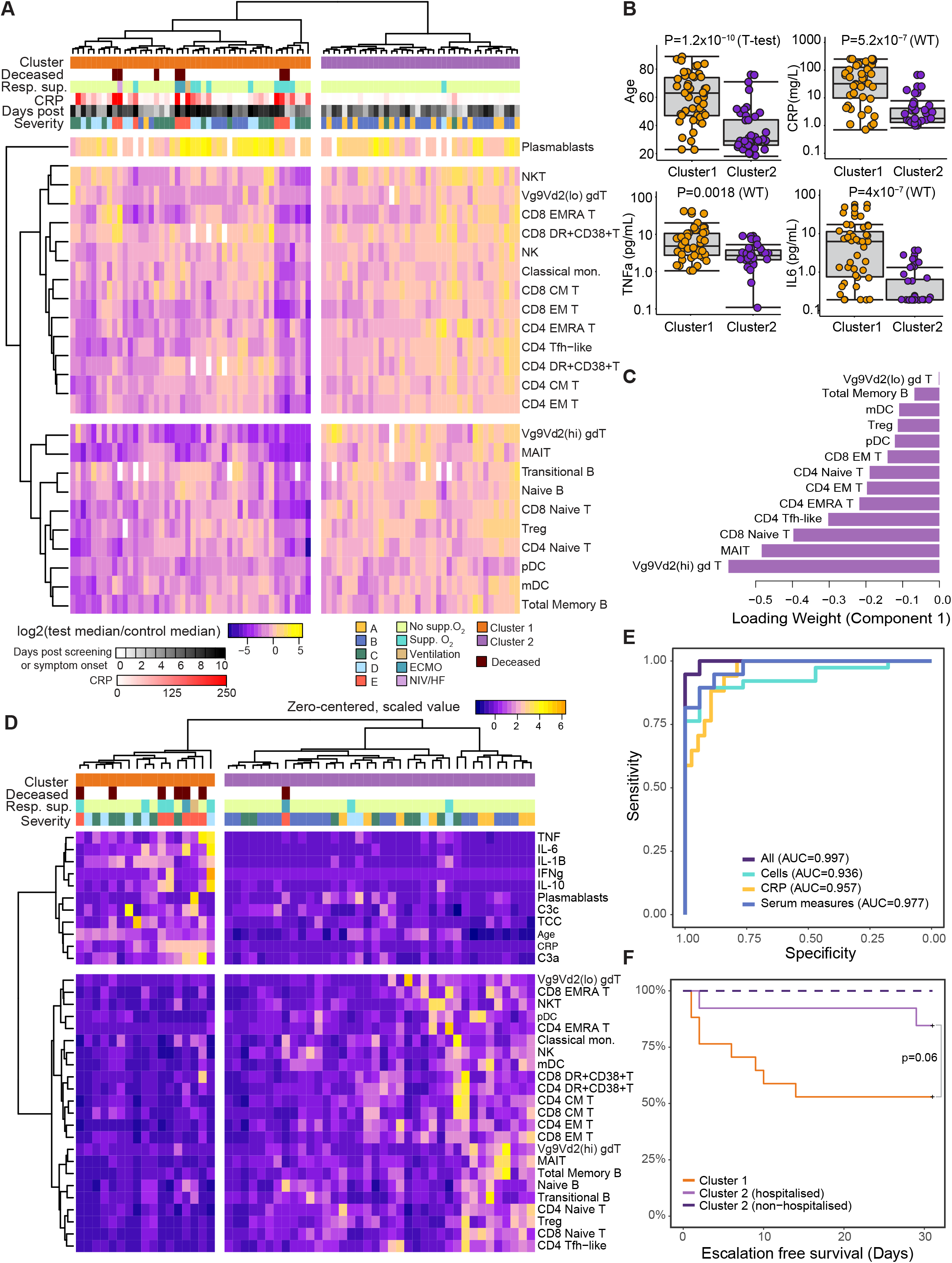
Multivariate analysis of immune-cell populations in early disease. **A)** Unsupervised clustering of absolute cell counts across 24 key cell populations (normalised to the median of healthy controls) for COVID-19 samples taken ≤10 days from screening (group A) or symptom onset (group B-E). Cases group into two clusters (cluster 1, orange; cluster 2, purple) by Euclidean distance and Ward D hierarchical clustering. **B)** Boxplots comparing age and inflammatory characteristics of individuals in clusters 1 and 2 at the time of sampling. **C)** Thirteen cell types selected by sPLS-DA as most informative in predictive models discriminating patients in clusters 1 and 2. Bars indicate loading coefficient weights of selected features (ranked from most to least informative in cluster prediction, from bottom to top). **D)** Addition of age, CRP, serum cytokine and complement measures to unsupervised clustering of cellular data in **A)** results in tighter grouping of COVID-19 patients by severity (cluster 1, orange, containing only patients in groups C-E and cluster 2, purple, predominantly patients in groups A and B). **E)** AUROC curve showing sensitivity and specificity of severity group prediction (as derived from clustering in D), based on absolute counts of 24 key cell types, CRP or serum measures alone compared to all available measures. **F)** Kaplan-Meier plot showing escalation free survival in individuals within severity clusters 1 or 2, split based on hospitalisation status. Escalation was defined as a step up in respiratory support or death. P-value for the chi-square test of the difference between cluster 1 and cluster 2 (hospitalised) survival curves is shown. CRP, C-reactive protein; IL, interleukin; TNF, tumour necrosis factor; IFN, interferon; C3, complement 3; TCC, terminal complement complex; NIV/HF, non-invasive ventilation/high-flow oxygen; ECMO, extracorporeal membrane oxygenation.

The ability to cluster patients and separate those with mild or no symptoms (AB) from those presenting to hospital (CDE) underlines the profound association of immune subset abnormalities with disease severity. It does not, however, predict outcome in a clinically useful way, as such predictions are only of practical value in those who present for medical attention. We therefore incorporated additional data related to inflammation, including cytokine levels, age, CRP and complement components; re-clustering with this combined dataset led to a smaller severity-associated group (**Figure 4D**). Input from both immune cell subsets and inflammation-related markers was required for optimal classification (**Figure 4E**). This clustering, when considering the patients recruited at initial hospital presentation (groups C, D and E), predicted disease progression, as defined by the subsequent need for increased respiratory support, or death, after blood sampling (**Figure 4F**). This analysis clearly requires validation in larger patient numbers, but is comparable to similar observations made by others (e.g. (Laing et al., 2020; Mathew et al., 2020)) and indicates that a combination of immune phenotype data and inflammatory markers could provide potentially clinically useful prediction of disease progression.

### Characteristics of a successful anti-SARS-CoV-2 immune response

HCW in categories A and B did not progress to severe COVID-19 disease, and they also cluster apart from those with more severe disease when using either early immune cell counts or RNA expression profiles (**Figure 4A and S5B**). We therefore compared the early features of these successful immune responses to those in patients with more severe COVID-19, to shed light on what differentiated effective antiviral responses from those associated with progression to severe disease - binned now in 7-day intervals to provide finer definition of the earlier immune response. For comparison, we show CRP and selected complement components (**Figure 5A**).

**Figure 5:**
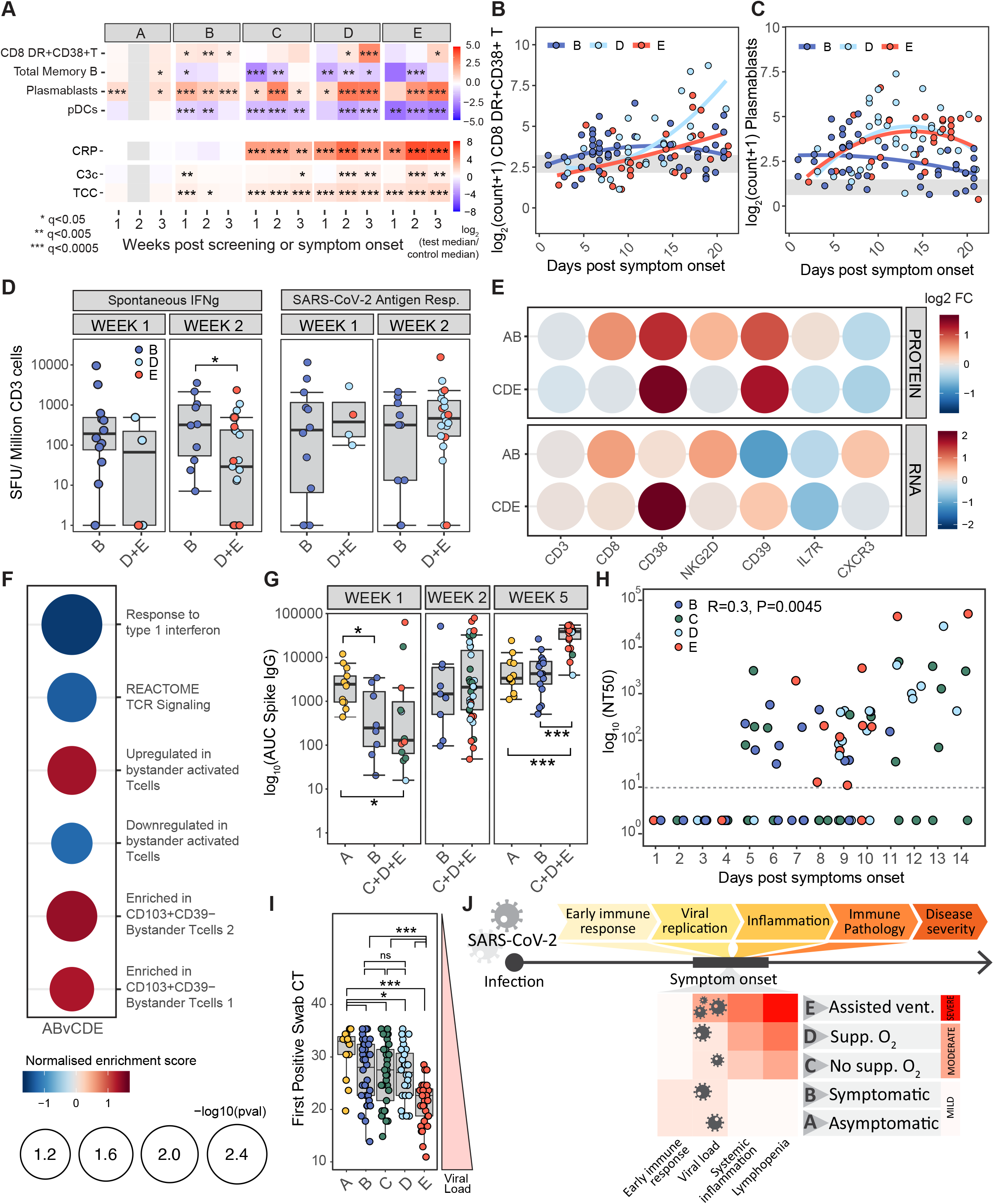
Early immune changes associated with mild disease and outcome. **A)** Heatmap showing the log_2_ fold change in median absolute cell counts, CRP or complement measures between COVID-19 cases and HCs, within severity categories and across 7-day time bins post screening (group A) or symptom onset (group B-E). Wilcoxon test FDR adjusted p-value: *<0.05, **<0.005, ***<0.0005. Mixed-effect model with quadratic time trend showing cellular trajectories over time in sample group B,D and E in non-naive HLA-DR+CD38+ CD8 T cells **B)** or plasmablasts **C)** (cells/uL) from weeks 1-3 post symptom onset. **D)** Number of CD3 T cells secreting IFN γ spontaneously or following SARS-CoV-2 antigen stimulation, in patient samples from groups B, and D and E combined, one or two weeks post symptom onset. Kruskal-Wallis test p-values: *<0.05. **E)** Log_2_ fold change (FC) in expression of CD8 T-cell transcripts capturing T-cell activation, and corresponding surface protein levels (as detected by antibody staining), in CITE-Seq data from non-naïve CD8 T-cells from patients in groups A and B, and C,D and E combined, relative to healthy controls **F)** Normalised gene set enrichment score for gene sets associated with TCR-dependent and bystander T-cell activation in single cell transcriptomic data derived from non-naïve CD8 T-cells from patients in groups A and B versus C,D and E. FDR adjusted P-value shown by circle diameter. **G)** Area under the curve for SARS-CoV-2 spike-specific IgG titres at 1, 2 and 5 weeks post screening (group A) or symptom onset (group B-E). Groups C,D and E are combined for increase statistical power, Kruskal-Wallis test p-values annotated as in **A**). **H)** SARS-CoV-2 neutralising antibody titres receiving 50% neutralisation (NT50) in patients from groups B-E in the first two weeks post symptom onset. Samples with no detectable neutralising activity at the lowest dilution (dotted line) are plotted at an arbitrary NT50 of 1. p-value and Pearson’s correlation shown. **I)** Boxplots showing SARS-Cov-2 viral load, taken as first positive swab PCR cycle threshold, in severity groups. Wilcoxon test p-values annotated as in **A**). **J)** Schematic summarising variation in immune features of SARS-CoV-2 infection across patients of varying disease severity. pDC, plasmacytoid dendritic cells, CRP, C-reactive protein; C3, complement 3; TCC, terminal complement complex SFU, spot forming units; FC, fold change, AUC; area under the curve; CT, cycle threshold.

A number of key features emerge. First, there is no evidence of systemic inflammation in groups A and B. CRP is normal (**Figure 5A**), cytokines are not raised (**Figure 1E**), and there is no RNA evidence of systemic inflammatory gene-related signatures (**Figures 3A and 3B**). The exception is the significant but transient increase in C3c and TCC. While plasmablasts rise, most cell types which are profoundly reduced in groups C to E are normal in A and B (**Figure 2 and S3**), but some show mild reductions in group B in particular, of which we show pDCs and memory B cells as examples (**Figure 5A-C, S6A and B**). pDCs fall to a lesser extent than in more severe disease, and for a shorter time period; a fall that is perhaps consistent with tissue localisation to allow local interferon production as part of a successful antiviral response (Cella et al., 1999).

There is an early increase in cytotoxic CD8 T cells seen in group B compared to the rise seen in groups C to E, with the increase seen by day 7 and peaking up to 2 weeks after symptom onset. This contrasts with the later and more sustained rise seen in the more severe COVID-19 patients (**Figures 5A, 5B and S6A**). There is enrichment of a CD8 cytotoxic RNA signature in group B by GSEA, which is significantly raised compared to group C to E between 0 and 24 days after symptom onset (**Figure S6C**). Consistent with these findings, spontaneous generation of IFNγ by T cells, as detected by ELISpot analyses, is more pronounced in group B at samples taken two weeks post symptom onset (**Figure 5D**).

Antigen-specific T cell responses were determined after simulation with anti-SARS-CoV-2 peptides and subtraction of background spontaneous IFNγ-producing T cells (see methods). The anti-SARS-CoV2-2 specific CD8 T cell response in group B was similar to more severe disease (**Figure 5D and S6D**), making it unlikely SARS-CoV-2 specific cells are solely responsible for the increased effector CD8 T cells seen in mild disease, suggesting a role for “bystander” activation. Such bystander responses are known to be important in early anti-viral defence (Maurice et al., 2021), and bystander-activated cells characteristically express higher cell surface NKG2D, involved in the killing of infected cells, and both IL-7 receptor and CD8, which are down-regulated in a TCR-dependent fashion (Kaech et al., 2003; Slifka and Whitton, 2000). Analysis by CITE-Seq of cell surface protein expression on activated CD8 T cells demonstrates a relative increase in NKG2D, IL7R and CD8 in group B compared to more severe groups (**Figure 5E**). These observations were confirmed at the RNA level (**Figure 5E**). Bystander CD8 T cells expressing CXCR3 rapidly home to sites of inflammation (Maurice et al., 2021), consistent with enrichment for CXCR3 RNA but not surface protein (**Figure 5E**). Enrichment of transcriptional signatures derived from TCR- and bystander-activated CD8 T cells was then assessed. This demonstrated enrichment of bystander-associated signatures in patients with mild disease and enrichment signatures of TCR activation in patients with severe COVID-19 (**Figure 5F**), again consistent with widespread early bystander activation in the CD8 T cell population in patients destined to have good disease outcomes.

There is also an early increase in plasmablasts seen in both groups A and B, again occurring up to a week before a larger rise is seen in more severe disease (**Figures 5C and S6B**). We therefore measured total immunoglobulin levels, anti-spike IgG, and anti-SARS-CoV-2 neutralising antibodies in group B, for comparison with patients progressing to more severe disease (**Figure 5G, H and S6E, F**). These patients maintained their serum IgM levels, which fell rapidly in those with more severe disease (**Figure S6G**). They had broadly equivalent titres of anti-spike IgG, and evidence of early neutralisation responses which were comparable to patients progressing to more severe COVID-19 (**Figure 5G and H**). This suggests that the B cell difference between patients with mild and more severe disease might lie in more robust non-antigen-specific B cell activation, perhaps impacting via “natural” antibody and/or non-antibody-dependent mechanisms.

Virus at first swab, as assessed by PCR CT value, is comparable in groups B, C and D, and is low in group A (again perhaps in part because these samples may be taken later after infection than symptomatic groups, see above). Initial viral titre was therefore not associated with an increased risk of hospital admission (being similar in groups B, C and D), but was significantly higher in group E than in other groups (**Figure 5I**). These viral titres are reflected in interferon-related transcription signatures, which are prominent in groups B-E (**Figures 3A, C and S6H**). Despite the fact that high IFN signatures correlate with severe disease, they are not necessarily driving that severity. The fact that the subgroup group with the highest IFN *within* group E do best (**Figure S6I**), suggests that the inflammatory pathways that create severe disease and push patients in to group E are distinct from IFN (which is also consistent with comparable kinetics of reduction of IFN signature regardless of severity – **Figure 3C**), and indeed that a robust IFN response may be beneficial in this context. That those in group E with low interferon signatures in early disease are more likely to have persistently high CRP appears consistent with this notion, though this needs confirmation in a larger dataset.

Taken together, these data suggest that an early adaptive immune response is prominent in individuals who are asymptomatic or have mild disease, characterised by a rapid production of activated bystander CD8 T cells, plasmablasts and likely pDC tissue localisation before antigen-specific responses become apparent. This appears a more important correlate of severity than viral titre, which only becomes relevant in those progressing to ventilation or death (**Figure 5J**).

In those with more severe disease (groups C-E), evidence of systemic inflammation is present from the first blood test, evident in samples taken from 0 and 7 days from the onset of symptoms (**Figure 5**) but evident early in that window. If we focus on the 16 patients in groups C-E sampled between 2 days before and 4 days after symptom onset, 15 had a CRP > 10 and/or neutrophil activation eigengene > 0. All 5 patients sampled between 2 days before and 2 days after symptom onset met these criteria. In contrast, this is not seen in groups A or B despite the fact that they mount a more prominent early immune response. It is not clear whether inflammation seen in C-E is a consequence, a cause or is indeed is only circumstantially related to the relatively poor immune response seen in patients who progress to severe disease. Nonetheless, it clearly does not develop later from the progression of a non-inflammatory immune response or as a result of failure to clear virus, and suggests that the inflammatory die is cast very early, and that strategies to prevent it, and to therefore reduce any impact it may have on immunity, would need to be established very early (**Figure 5J**).

### Distinct patterns of immune recovery in COVID-19

In contrast to groups A and B, cellular changes in groups C - E were profound and usually most prominent at the first bleed (**Figure 2**). Determination of the rate and direction of change among immune cell subsets over time was likely to be most informative in these groups, and we therefore explored immune cell kinetics in groups C, D and E, assigning patients to two categories based on whether their CRP levels remained elevated above 10mg/L (“Persisting CRP”) or fell below 10mg/L (“Resolving CRP”) by their final bleed within 3 months post symptom onset (**Figure 6A**). The latter group included both individuals with high CRP levels early, and those for which CRP remained low (10mg/L) at all measured time points over the course of study. Changes in CRP over time differed significantly between these two groups when assessed using a mixed-effects model, with time modelled as a continuous variable (**Figure 6B**).

**Figure 6:**
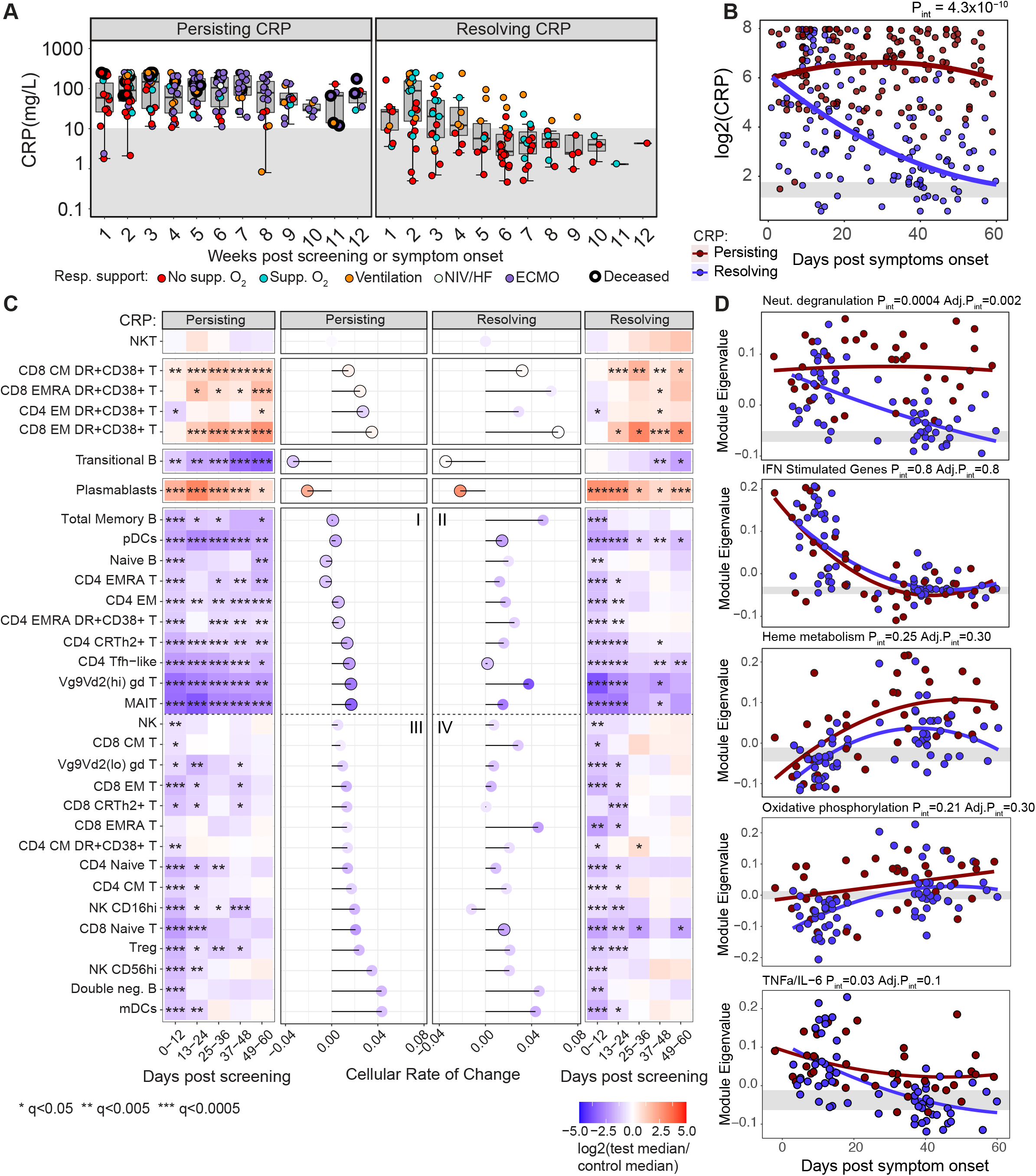
Cellular and transcriptional trajectories in persisting and resolving disease. **A)** CRP (mg/L) over 7-day time bins (weeks) in patients from groups C, D and E grouped by persisting and resolving CRP. **B)** Mixed-effect model with quadratic time trend showing log_2_(CRP) trajectories in both patient groups, the likelihood-ratio test p-value for the time x group interaction term is reported. Grey band indicates the interquartile range of the corresponding measure in HCs. **C)** Heatmap showing the log_2_ fold change in median absolute cell count between COVID-19 cases in groups C, D and E, split according to persisting or resolving CRP, and HCs across 12-day time bins. Wilcoxon test FDR adjusted p-value: *<0.05, **<0.005, ***<0.0005. Rate of cellular change is shown for each cell by lollipop plot; a faster rate of recovery, or deviation from normal, is indicated by increasing stem length. Points are coloured by log_2_ fold change in median absolute cell counts from HC at 0-12 days, black outline indicates failure to recover to HC levels within 60 days (as defined in methods). **D)** Mixed-effect models showing longitudinal trajectories of gene module eigenvalues capturing neutrophil degranulation, interferon stimulated genes, heme metabolism and oxidative phosphorylation in CRP groups, p-values reported as **B)**. CRP, C-reactive protein; NIV/HF, non-invasive ventilation/high-flow oxygen; ECMO, extracorporeal membrane oxygenation.

In order to compare the nature of cellular changes over time, both across cell subsets and between persisting and resolving CRP patient groups, a “rate of change” for each cell population was calculated over 60 days post symptom onset. In brief, this rate captured both the initial deviation in cell counts from normal within a window of 0-12 days, and the time taken for cells counts to stabilise within a normal range if cellular recovery did occur (see Methods). Five predominant trajectories were observed; populations that did not deviate from heathy levels over the duration of study (e.g. NKT cells), those which increased progressively from normal over time (e.g. effector CD8 T cells), those which fell progressively from normal over time (e.g. transitional B cells), those which trended toward recovery after an initial rise in numbers (plasmablasts) and those which tended toward recovery after an initial drop in numbers (e.g. naïve CD4 T cells) (**Figure 6C**).

The absolute number of most cell populations fell precipitously early, and then showed variable degrees of recovery. For descriptive purposes these are arranged into a group of cell subtypes that failed to recover, or recovered, in the persisting CRP group (**Figure 6C** quadrants I and III respectively), and their equivalents in the resolving CRP group (**Figure 6C** quadrants II and IV respectively). Notably, a few populations remained consistently abnormal in both persisting and resolving CRP groups out to 60 days post symptom onset, including pDCs, Tfh, MAIT cells and Vg9Vd2 (hi) γδ T cells. All other populations showing an early drop in counts (with the exception of naïve CD8 T cells) recovered to normal levels in the patients with resolving CRP (II and IV), and at rates more rapid than seen in those with persisting high CRP values. In the persisting CRP group, a number of cell types remained markedly abnormal (including memory B cells and various CD4 T cell subsets: quadrant I), whereas a second group of cell types recovered despite persisting inflammation (including NK cells and a number of CD8 T-cell subsets: quadrant III).

We then explored the relationship between cell recovery and the nature and kinetics of the inflammatory response. It is not surprising that where the CRP remains persistently elevated, immune defects might persist, on the assumption that these defects are secondary to the inflammatory state. Consistent with this, the cohort with persistently raised CRP also has raised TNF-α and IL-6 at the protein level over time (**Figure 1D**). Likewise, transcriptional signatures of TNF-α, IL-6 and neutrophil activation were increased in severe disease (**Figure 3**), particularly in the persistent CRP group (**Figure 6D**). This ongoing inflammation may contribute to the sustained reduction in cell numbers at late times seen in quadrant I, together with persistently raised HLA-DR+/CD38+ effector T cells and plasmablasts (**Figure 6C**). Consistent with this, it is also perhaps not unexpected that most cell types reduced in acute disease recover over a few weeks as the CRP falls, as is seen for most cells in quadrants II and IV (**Figure 6C**).

More intriguing are the cells that recover rapidly in the face of ongoing inflammation (quadrant III). While the reasons for this are likely to differ between cell types, and to be multifactorial, there is the possibility that some of these cell reductions are driven by the viral infection *per se*, virus-induced interferon, or a combination of the two. It is notable that, after an initial rise, IFN-γ returns to normal in patients irrespective of disease severity (**Figures 1E and S1B**). Interferon-stimulated gene (ISG) signatures fall to normal levels over about 3 weeks independent of disease severity group (**Figure 3C**) and CRP (**Figure 6D**), but correlating with declining virus titre (**Figures 3D and S4E**). Thus, cell types known to leave the circulation due to interferon stimulation (Kamphuis et al., 2006), such as T and NK cells (Hirsch and Johnson, 1986; Zafranskaya et al., 2007), may recover as interferon-dependent inflammation falls, presumably as a result of control of viral infection and independently of ongoing CRP-associated inflammation.

Finally, a small number of cell types remain statistically abnormal after 60 days, even in the resolving CRP group. These include effector CD4 and CD8 T cells (HLA-DR+/CD38+), and plasmablasts, all of which remain elevated, and pDCs, Tfh, Vg9Vd2 expressing γδ T cells and MAIT cells, which remain reduced (**Figure 6C**) and are among those cells most predictive of poor prognosis (**Figure 4C**). These abnormalities persist despite resolution of CRP-reflected inflammation, with evidence for neutrophil degranulation, TNF-α /IL-6 and glycolysis all falling alongside CRP (e.g. **Figures 1D and 6D**). They also persist despite early resolution of interferon-stimulated gene signatures in all severity groups. Possible mechanisms behind these sustained abnormalities are discussed below.

### The late appearance of OXPHOS and ROS pathways correlates with differential immune recovery

At late time points, whole blood transcriptome analysis shows an increase in inflammation-related signatures distinct from those that are prominent early in the disease course, particularly in severity groups C-E. These signatures are characteristic of oxidative phosphorylation (OXPHOS), reactive oxygen species generation (ROS) and heme metabolism. These are demonstrated in an un-biased fashion using WGCNA, where modules characterised by OXPHOS and heme metabolism signatures are prominent in samples analysed at day 25 to 48 post symptoms, with OXPHOS most prominent in group E, and heme metabolism in C, D and E (**Figures 3B and 3C**). Enrichment of Hallmark signatures in RNA-seq datasets confirmed the association of OXPHOS and heme metabolism in groups C, D and E, and also found association of a ROS signature (**Figures 3E and 7A**). Consistent with this, the expression of the genes driving the enrichment of each signature was upregulated in the three most severe clinical groups (**Figure 7B and Table S3**). The late rise in these three correlated signatures occurs irrespective of persisting or resolving CRP-associated inflammation (**Figure 6D**), and appears independent of specific cell population recovery (**Figure S7A**).

**Figure 7:**
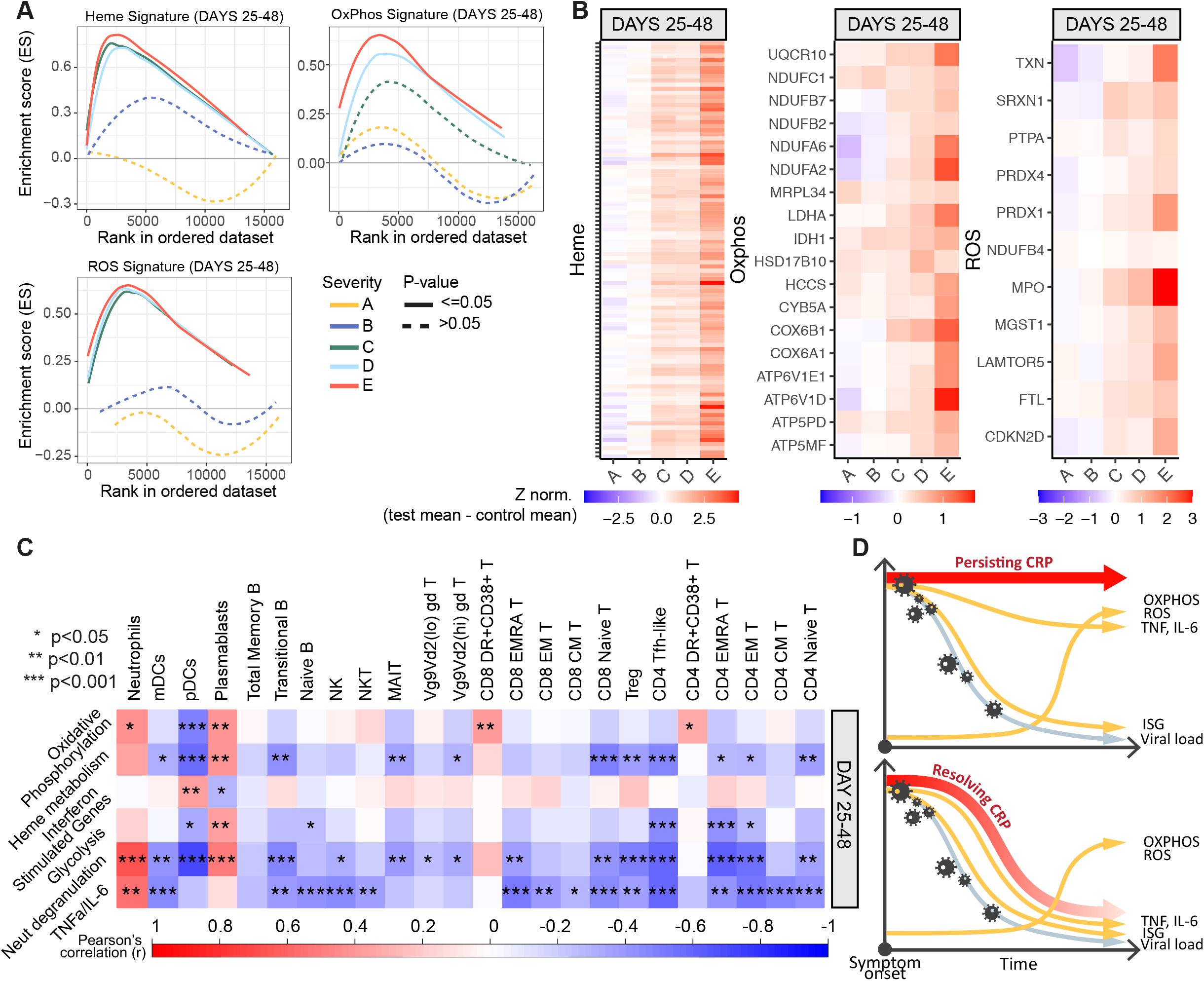
Transcriptional changes in prolonged disease. **A)** Enrichment score for HALLMARK genesets capturing heme metabolism, oxidative phosphorylation and ROS related genes (as determined by GSEA) in group A-E samples taken 25-48 days post screening (group A) or symptom onset (groups B-E). **B**) Heatmap showing relative expression of the intersection of GSEA leading edge genes from groups C, D and E, across all severity groups in samples taken 25-48 days post screening (group A) or symptom onset (groups B-E). **C)** Heatmap showing correlation between transcriptional eigengenes and absolute cell counts, at 25-48 days post symptom onset. Boxes are coloured by strength of correlation, Pearson correlation p-values: *<0.05, **<0.01, ***<0.001. **D)** Schematic representation of the trajectory of immunological changes in SARS-CoV2 infection over time in patients with persisting or resolving systemic inflammation.

We then sought correlation between cellular and transcript signature changes in COVID-19. In the first 24 days after symptom onset, there is a strong association between TNF-α /IL-6, neutrophil degranulation and interferon signatures with most of the lymphoid cell types whose numbers fall in severe disease (**Figure S7B**). Later, between 24 and 48 days after symptom onset, these associations change (**Figure 7C and S7C)**. While TNF-α /IL-6 and neutrophil degranulation signatures are still associated with many cell subsets that continue to be reduced, the interferon signature is no longer a significant player. Strikingly, the persistent increase seen in effector lymphocytes, both CD4 and CD8 activated T cells (HLA-DR+, CD38+) and plasmablasts, is now particularly associated with the OXPHOS signature which, having become more prominent later in disease (**Figure 3B**), has a much more restricted and specific association with immune dysfunction than other inflammatory signatures.

It is thus clear that, for some cell types, the association with the inflammatory milieu changes over time, but for others it is more consistent. It is interestingly the inflammatory signatures which appear late in disease, in particular OXPHOS, are specifically associated with persistent derangement of cell types of potential pathological importance, such as increased HLA-DR+CD38+ T cells and plasmablasts, and reduced pDCs (**Figure 7D)**.

## Discussion

In asymptomatic or mildly symptomatic, non-progressive SARS-CoV-2 infection (groups A and B) there is evidence of an early robust adaptive immune response. Circulating plasmablasts and CD8^+^HLA-DR+CD38+ activated T cells expand earlier and in higher numbers than in more severe COVID-19 groups, most notably in the first week after symptom onset. Both of these cell populations then contract in A and B, as they continue to rise in groups C-E. Despite this, and a prominent early interferon transcription signature, both the antibody and T cell SARS-CoV-2-specific immune responses were comparable between early mild and severe disease. This suggested the increased plasmablasts and effector CD8 T cells could reflect an enhanced bystander response in mild disease, something then supported by single cell CITE-Seq analysis.

Bystander CD8 T cell activation was first described in the context of LCMV infection in mice (Tough et al., 1996). It involves activation of memory CD8 T cells independent of TCR stimulation (whether by the initiating or cross-reactive antigen) (Maurice et al., 2021). It is driven by type 1 IFN in many inflammatory contexts, in particular viral infection. Bystander CD8 T cells are activated long before the antigen-specific response is seen, being detectable within a day of viral infection, and often peaking within a week (Berg et al., 2003). Bystander cells can migrate to infected organs using CXCR3, and kill virally infected cells through NKG2D and granzyme B-dependent mechanisms (Maurice et al., 2019). They have both been shown to be critically important in controlling the early defence against viral infections, in part by direct anti-viral effect, and in part by producing IFN-gamma and other cytokines which provide vital activation signals for antigen presenting cells (Soudja et al., 2014) and control the memory/effector balance of subsequent antigen-specific responses (Krummel et al., 2018). Similar, though less well defined, bystander phenomena occur in CD4, MAIT and gamma delta T cell responses (Holzapfel et al., 2014; Ribeiro et al., 2015; Ussher et al., 2018). The early and pronounced production of bystander-activated CD8 T cells in patients groups A and B could be directly related to disease resolution and prevention of progression to more severe disease, a process likely to begin well before symptom onset. Understanding the factors which impede this early bystander response could be important for developing strategies to prevent severe disease.

In contrast, persistent bystander CD8 T cell activation has been associated with inflammatory pathology in the context of both chronic infection and autoimmunity. NKG2D-dependent killing of non-virally infected hepatocytes in hepatitis A exacerbates liver damage (Kim et al., 2018) and may also play a role in inflammatory lung disease (Borchers et al., 2009), and there is evidence that NKG2D ligands are expressed in the lungs in COVID-19 (Wauters et al., 2021), so it could be that a late and persistent expansion of bystander-activated effector T cells in severe COVID-19 could be a driver of lung pathology. The well-documented links between bystander CD8 T cell activation and autoimmunity (Groh et al., 2003; Meresse et al., 2004) also raises the prospect they may play a role in the autoimmune manifestations that have been observed in COVID-19.

The early increase in circulating plasmablasts observed in mild COVID-19 are likely to be comprised of SARS-CoV-2 specific cells, perhaps to an extent not yet fully reflected in serum antibody, together with non-SARS-CoV-2-specific bystander activated cells. Bystander B cell activation is known to involve memory B cells (Horns et al., 2020) as well as innate-like B cells (including B1 cells in the mouse, and marginal zone cells in both mouse and human). Their function is not fully defined, but includes an increase in production of “natural antibodies” (Antin et al., 1986) alongside non-antibody-associated functions, which may include antigen transport to secondary lymphoid organs, antigen presentation to T cells, cytokine production and a broader role in immune regulation. These “bystander” B cell functions are known to play a role in early defence against bacterial infection (Boes et al., 1998; Haas et al., 2005; Ochsenbein et al., 1999), and may be involved in autoimmunity (Espeli et al., 2019; Sanderson et al., 2017). Their role in viral infection is less clear, but it may be that early bystander B cell activation helps determine the outcome of COVID-19, in a manner analogous to the role proposed for bystander CD8 T cells. A closer investigation of the early B cell response in COVID-19 will be required to confirm this.

At the same time as these signs of an early adaptive immune response are seen in groups A and B, there is no evidence of systemic inflammation, apart from some early, transient complement activation. CRP, circulating TNF-α and IL-6, and transcriptional signatures of a number of inflammatory pathways (including those associated with neutrophil activation and TNF-α /IL6 signalling) are not raised in groups A and B, at times when they are already prominent in groups C-E. The severe and widespread leucocyte subset depletion seen at the initial bleed in the more severe COVID-19 patient groups C-E, and observed by many others (Carissimo et al., 2020; Laing et al., 2020; Mathew et al., 2020; Su et al., 2020), is not apparent in those with asymptomatic or mildly symptomatic disease, suggesting this too is a unique feature of a pathological immune response. Immune cell subset numbers correlate strongly with severe and progressive disease (**Figure 4A**). Coupled with evidence of early systemic inflammation in severity groups C-E, our findings suggest that the immune pathology associated with severe COVID-19 is either established immediately post-infection or, if there is a transition point from an effective to a pathological response, this is likely to occur before the time of symptom onset (**Figure 5J**). This finding may have major implications as to how disease needs to be managed, since intervention to prevent immune pathology would need to be targeted very early in the disease course, and perhaps pre-emptively in high risk groups screened and diagnosed before symptoms develop.

The reason for the failure to mount a robust early B and T cell response in the context of severe COVID-19 is likely to be multifactorial. There is no evidence for a relationship with viral load and progression to inflammatory disease, as initial viral titres were comparable between group B and groups C and D. Once inflammatory disease is established, however, viral titre may be associated with subsequent outcome, as early increased viral titre is seen in group E, consistent with reports that high initial viral titre might be associated with mortality (Pujadas et al., 2020). Genetic association studies in severe COVID-19 point to genes that are implicated in driving antiviral responses. The most prominent are associations with genes involved in the type 1 interferon pathway (Pairo-Castineira et al., 2020; Zhang et al., 2020), known to be the key driver of bystander T cell activation (Maurice et al., 2021). Increasing age and comorbidity such as diabetes and chronic inflammatory disease are known to suppress early CD8 T and B cell responses (Shen-Orr et al., 2016; Weiskopf et al., 2009). An in-depth understanding of these risk factors may instruct screening strategies to assess risk of progression before inflammatory responses become self-sustained.

While clear distinctions in immune responsiveness were apparent between groups A and B versus groups C, D and E, differences between the more severe groups themselves were less obvious. Those with symptomatic disease warranting admission to hospital clustered together using the size of just 13 key cell populations: these clusters correlated strongly with clinical severity, and immune cell subset numbers together with inflammatory mediator levels provided prediction of subsequent progression, as well as COVID-19 associated death. Similar observations have been made by others (Chen and Wherry, 2020) though whether the predictive ability of clustering by immune parameters adds to that provided by clinical and other predictors remains to be determined. While determining prognosis after presentation to hospital could be of clinical use, it would be of more benefit to predict progression to severe disease in cases with milder COVID-19 – but it is not clear whether this will be possible in practice, given our observation that inflammatory immunopathology is likely to be present at symptom onset. A study to address this issue would need to be conducted in particularly high-risk patient groups to ensure an adequate event rate, and require diagnosis through asymptomatic screening to detect changes before symptoms develop.

We can find no equivalent studies in other severe infections which explore the kinetics of the cellular immune response across this range of time and clinical severity. Nonetheless, many of the changes seen in response to SARS-CoV-2 have been reported in the context of other viral infections. Both an early reduction in blood pDC and T cells has been shown in RSV (Russell et al., 2017) and influenza A (Fox et al., 2012; Lichtner et al., 2011), for example. NK cells in the blood also fall in influenza, probably as a result of migration to the lungs. Finally, plasmablast expansion has also been seen, particularly in dengue fever (Wrammert et al., 2012). The nature of these studies makes a direct comparison with the situation in SARS-CoV-2 difficult, but it would seem likely that many immunological changes seen in COVID-19 mirror those seen in responses to other infections, though in general the cellular changes we observe appear to be more profound, peak earlier in disease course, and are more persistent in COVID-19 than in studies performed in other infectious contexts.

The recovery of the profound immune dysregulation seen in those with severe COVID-19 is potentially of major clinical relevance, as such recovery may be important in the resolution of inflammatory disease or be required to prevent secondary infection or SARS-CoV-2 reinfection. and persistent immunopathology could be associated with clinical manifestations of “long COVID”. We find immune cell abnormalities often persist for weeks to months after SARS-CoV-2 infection, and different cell populations exhibit strikingly different patterns of resolution. Some recover as systemic inflammation (as measured by CRP) resolves, others remain persistently abnormal despite normalisation of CRP, and a third group resolve even in the face of persistent systemic inflammation.

Understanding the different inflammatory drivers or associations of this differential recovery could provide insight into the immune pathology of COVID-19, and potentially of other infections. Here, to begin to explore this, we correlated immune changes with measurements of systemic inflammation throughout the disease course. Patients with severe COVID are characterised by high CRP, and this correlates with evidence of TNF-α and IL-6 driven processes both at the protein and transcriptome level, as well as with both neutrophil activation and glycolytic metabolism. The fact that many cellular abnormalities persist while these biologic processes are apparent – while others appear to resolve alongside them – suggests that the nature of systemic inflammation is important in driving different aspects of immune pathology.

Finally, some cell populations remain markedly abnormal, or show a limited recovery, even once CRP-associated inflammation has resolved, and indeed after patients have been discharged from hospital. These persistent changes may reflect a slow intrinsic regenerative capacity of the cell type concerned, but in other situations, such as the continued elevation of effector T and B cells, it is tempting to speculate that there is ongoing abnormal signalling driving such changes. For that reason, we explored late changes that are seen in the inflammatory response in COVID-19. Interestingly, three transcriptional signatures arise late in those with severe COVID-19 that are not present in early severe, nor mild, disease. These include activation of OXPHOS-, ROS- and heme-related metabolic pathways (**Figure 7D**). Activation of immune cells results in metabolic reprogramming that supports cell growth, proliferation and differentiation. Disruption of metabolic pathways can result in bioenergetic, anabolic, epigenetic or redox cellular crises – culminating in immune dysfunction (Bantug et al., 2018). It is unlikely that the metabolic signatures observed here simply reflect heightened bioenergetic requirements of activated immune cells, as one would expect similar requirements to be also present early in disease. OXPHOS can drive inflammation (Mills et al., 2017), and it is intriguing to note COVID-19 patients treated with metformin, which inhibits Complex I of the respiratory chain, had lower amounts of circulating inflammatory cytokines (Cheng et al., 2020). The ROS transcriptional signature may reflect more abundant production of ROS-species inevitably accompanying increased OXPHOS. Alternatively, it may reflect specific mitochondrial pathology, and thus *per se* contribute to immune cell dysfunction (Nathan and Cunningham-Bussel, 2013) Mitochondria are also critically involved in heme biosynthesis. Heme serves as a prosthetic group for haemoglobin as well as many other proteins – including several that constitute the respiratory chain of mitochondria. While free heme can act as damage-associated molecular pattern and promote ROS formation, the role of heme biosynthesis *vs*. catabolism in balancing cellular sensitivity to oxidants is complex and context dependent (Prestes et al., 2020). Here, given correlated regulation of heme and OXPHOS pathways in the clinical categories C - E, activity of these modules may be interrelated and possibly together reflect dysfunctional mitochondria. How heme and OXPHOS transcriptional programmes are linked on a molecular level cannot be inferred from our data. Erythroid cell activation has recently been detected in severe COVID-19 (Bernardes et al., 2020) and could also contribute to a heme transcriptional signature. However, the increase in heme metabolism in our cohort correlates strongly with falling haemoglobin, and reticulocytes in patients in groups C, D and E are low (**Figure S7C and D**) – suggesting suppression rather than activation of erythropoiesis. Understanding the mechanism linking metabolic dysregulation to persistent immune pathology in COVID-19, and also to “long COVID”, will require further study over longer disease courses.

## Supporting information

Supplementary Item 1

Supplemental Item 2

Supplementary Item 3

Supplementary Item 4

## Data Availability

In progress and we will update when available

## Acknowledgements

We thank all the patients and Health Care Workers who consented to take part in this study. We are grateful for the generous support of CVC Capital Partners, the Evelyn Trust (20/75), UKRI COVID Immunology Consortium, Addenbrooke’s Charitable Trust (12/20A), the NIHR Cambridge Biomedical Research Centre and the UKRI/NIHR through the UK Coronavirus Immunology Consortium (UK-CIC) for their financial support for their financial support. K.G.C.S. is the recipient of a Wellcome Investigator Award (200871/Z/16/Z); M.P.W. is the recipient of Wellcome Senior Clinical Research Fellowship (108070/Z/15/Z); C.H. was funded by a Wellcome COVID-19 Rapid Response DCF and the Fondation Botnar; N.M. was funded by the MRC (CSF MR/P008801/1), NHSBT (WPA15-02) and Addenbrooke’s Charitable Trust (grant ref. to 900239 NJM); I.G.G. is a Wellcome Senior Fellow and was supported by funding from the Wellcome (Ref: 207498/Z/17/Z). P.J.L. is the recipient of a Wellcome Trust Principal Research Fellowship (084957/Z/08/Z) and MRC research grant (MR/V011561/1). We would like to thank: the NIHR Cambridge Clinic Research Facility outreach team for enrolment of patients; the NIHR Cambridge Biomedical Research Centre Cell Phenotyping Hub and the CRUK Cambridge Institute flow cytometry core facility for their support with flow and mass cytometry; and the Cambridge NIHR BRC Stratified Medicine Core Laboratory NGS Hub (supported by an MRC Clinical Infrastructure Award) for their support with whole blood RNA-Sequencing.

## Author Contributions

Conceptualization: C.H., J.R.B., P.A.L., and K.G.C.S.; Methodology: L.B., L.T., M.R.W., I.G.G., and R.D.; Software: K.H.; Validation: L.T.; Formal Analysis: L.B., F.M., L.T., A.H., P.K., and H.R.; Data Curation: F.M., A.H., P.K., L.T.; Investigation: L.B., F.M., L.T., P.K., A.D.S., O.H., M.D.M., P.P.G., M.R.W., I.G.G., M.H., F.J.C., R.D., J.K.N., and E.J.M.T.; Visualization: A.H., and P.K; Project Administration: L.B., F.M., L.T., B.J.D, C.S. and A.E.; Funding Acquisition: P.J.L., N.K., J.R.B., C.H., G.D., M.P.W., P.A.L, and K.G.C.S.; Supervision: J.R.B, P.A.L and K.G.C.S.; Resources: A.M.P., M.T., M.P.W., N.J.M.; Writing – Original Draft: P.A.L, P.J.L., C.H. and K.G.C.S; Writing – Review & Editing: L.B., F.M., A.H., L.T., P.K., H.R., B.J.D., A.D.S., O.H., M.D.M., P.P.G., F.J.C., R.D., K.H., S.B., R.K.G., J.K.N., C.S., A.E., A.M.P., E.J.M.T., N.K., P.J.L., N.J.M., S.R., J.E.D.T., M.P.W., B.G., M.T., C.H., J.R.B, G.D., M.R.W., M.H., I.G.G., P.A.L., and K.G.C.S.

## Methods

### Participant recruitment

Study participants were recruited between 31/3/2020 and 20/7/2020 from patients attending Addenbrooke’s Hospital with a suspected or nucleic acid amplification test (NAAT) confirmed diagnosis of COVID-19 (including point of care testing (Collier et al., 2020; Mlcochova et al., 2020)), patients admitted to Royal Papworth Hospital NHS Foundation Trust or Cambridge and Peterborough Foundation Trust with a confirmed diagnosis of COVID-19, together with Health Care Workers identified through staff screening as PCR positive for SARS-CoV-2 (Rivett et al., 2020). Controls were recruited among hospital staff attending Addenbrooke’s serology screening programme, and selected to cover the whole age spectrum of COVID-19 positive study participants, across both genders. Only controls with negative serology results (45 out of 47) were subsequently included in the study. Recruitment of inpatients at Addenbrooke’s Hospital and Health Care Workers was undertaken by the NIHR Cambridge Clinical Research Facility outreach team and the NIHR BioResource research nurse team. Ethical approval was obtained from the East of England – Cambridge Central Research Ethics Committee (“NIHR BioResource” REC ref 17/EE/0025, and “Genetic variation AND Altered Leucocyte Function in health and disease - GANDALF” REC ref 08/H0308/176). All participants provided informed consent.

Inpatients were sampled at study entry, and then at regular intervals as long as they remained admitted to hospital (approximately weekly up to 4 weeks, and then every 2 weeks up to 12 weeks). Discharged patients were invited to provide a follow-up sample 4-8 weeks after study enrolment. Health care workers were sampled at study entry, and subsequently after approximately 2 and 4 weeks. At each time-point, blood samples were drawn in EDTA, sodium citrate, serum and PAXgene Blood RNA tubes (BD Biosciences).

### Clinical data collection

Clinical data were retrospectively collected by review of medical charts and entered into spreadsheets or Castor EDC, a cloud-based clinical data management system. Available laboratory test results and administered in-patient medications were extracted from Epic electronic health records (Addenbrooke’s Hospital) and from MetaVision ICU (RPH ITU). Data were merged from the various data sources using R version 3.6 and the R packages readr (1.3.1), openxlsx (4.1.4), dplyr (0.8.3), tidyr (1.0.2) and lubridate (1.7.4).

Health care workers were classified in 2 groups (A and B) according to whether they were asymptomatic (group A) or had possible COVID-19 symptoms (group B) at the time of PCR testing. For this purpose, new-onset fever (>37.8 C), cough, loss of sense of smell, hoarseness, nasal discharge or congestion, shortness of breath, wheeze, headache, muscle aches, nausea, vomiting and diarrhoea were considered as possible COVID-19 symptoms.

Participants in group A were further sub-grouped according to whether they were completely asymptomatic (n= 8), or had had any of the above COVID-19 symptoms before PCR testing (n=10, median time from symptoms to COVID-19 PCR test 26 days, range 9-42 days).

Group B participants included both staff who were self-isolating because of COVID-19 symptoms (n=30), and staff members who were reporting fit for duty, but had some symptoms that did not reach the threshold for self-isolation at that time (n=10).

Hospital patients were assigned to 3 severity groups, mainly reflecting the maximum level of respiratory support for COVID-19 received during their hospital stay:

- group C: did not receive any supplemental oxygen. Five patients were discharged soon after initial diagnosis and assessment but followed up as part of the study.
- group D: received supplemental oxygen using low flow nasal prongs, simple face mask, Venturi mask or non-rebreather face mask.
- group E: received any of non-invasive ventilation (NIV), mechanical ventilation or ECMO. Patients who received supplemental oxygen (but no ventilation) and deceased in hospital were also assigned to group E.

In patients who were already established on home NIV for chronic respiratory failure, NIV delivered as per the home prescription (e.g. nocturnal) was not considered for the purpose of classification. Moreover, oxygen requirements that were clearly not related to COVID-19 were also not considered for classification purposes. In particular, 2 cases who received low flow supplemental oxygen for non-COVID-19 indications (ascitic splinting in decompensated cirrhosis in one case, and recovery from anesthesia after orthopedic surgery in the other) were assigned to group C. Cases in group C were further sub-classified according to chest radiology results (X-ray and, if available, CT scan), as:

- abnormal radiology: chest X-ray/ CT scan showed changes compatible with COVID-19
- normal radiology: chest X-ray/ CT scan did not show any abnormality compatible with COVID-19 (reported as normal or showing lung changes diagnostic of conditions other than COVID-19).

Immunological parameters were analyzed according to time since onset of symptoms, or otherwise time since positive SARS-CoV-2 NAAT (in group A and in 4 asymptomatic patients in group C). Seven cases admitted to hospital for COVID-19 had no date of onset of symptoms documented in the medical records. In these cases, the date of onset of symptoms was estimated as follows: hospital admission date - median time from symptoms to hospital admission in patients admitted for COVID-19.

Following clinician review, 6 cases were considered not classifiable, due to complex concomitant pathologies that coexisted with COVID-19 and dominated the clinical picture, confounding the interpretation of clinical outcome. These cases were not included in any analyses; more details are reported in **Table S2. Item S1** summarizes the timing of research samples and clinical trajectories for volunteers in severity groups C, D and E included in the analysis.

### Peripheral blood mononuclear cell preparation and flow immunophenotyping

Each participant provided 27 mL of peripheral venous blood collected into 9 mL sodium citrate tube. Peripheral blood mononuclear cells (PBMCs) were isolated using Leucosep tubes (Greiner Bio-One) with Histopaque 1077 (Sigma) by centrifugation at 800x g for 15 minutes at room temperature. PBMCs at the interface were collected, rinsed twice with autoMACS running buffer (Miltenyi Biotech) and cryopreserved in FBS with 10% DMSO. All samples were processed within 4 hours of collection.

Five distinct antibody cocktails (**Table S4**) were used to label approximately 10^6^ PBMCs using standard methods. T regulatory cells were fixed and permeabilised following surface staining prior to the addition of intracellular antibodies. Samples were stored at 4°C and acquired within 4 hours using a 5-laser BD Symphony X-50 flow cytometer. Single colour compensation tubes (BD CompBeads) or cells were prepared for each of the fluorophores used and acquired at the start of each flow cytometer run.

For direct enumeration of T, B and NK cells, an aliquot of whole blood (50 μl) was added to BD TruCount™ tubes with 20μl BD Multitest™ 6-colour TBNK reagent (BD Biosciences) and processed as per the manufacturer’s instructions.

Samples were gated in FlowJo v10.2 according to the schema set out in **Item S4**. The number of cells falling within each gate was recorded. For analysis, these were expressed as an absolute concentration of cells per μl, calculated using the proportions of daughter populations present within the parent population determined using the BD TruCount™system.

### CyTOF

The protocol used to isolate PBMCs led to an impaired recovery of the different monocytes population, specifically intermediate and non-classical monocytes. To extend our analysis to these and other granolucyte populations we performed mass cytometric analysis on a subgroup of patients and healthy controls (249 samples). Briefly, whole blood samples (270μl) were stained using the Fluidigm Maxpar® Direct™ Immune Profiling Assay™ according to the manufacturer’s instructions. Samples were cryopreserved at −80°C following staining and thawed for analysis within 4 weeks. Samples were acquired using a Fluidigm Helios mass cytometer and normalised using the CyTOF Software v6.7.1016. FCS files generated were analysed using the Maxpar® Pathsetter™ software v2.0.45 (Verity Software House, Topsham, ME). Standard settings were used to generate immune cell frequencies for 37 immune cell populations. Absolute cell numbers were calculated using the proportions of these immune cell populations within the parent populations determined by BD TruCount™.

### Reticulocyte counts

Reticulocyte numbers were measured using a Sysmex XN-1000 haematology analyser as previously described (Akbari et al., 2020).

### Complement

Complement activation was assessed by measuring C3 activation products (C3a and C3c) together with the terminal complement complex (TCC) as an end product of the complement cascade. Concentrations of these complement components were measured in EDTA plasma from patients using commercially available enzyme-linked immunosorbent assays (ELISA) kits (HK354 (C3a), HK368 (C3c), HK328 (TCC), Hycult Biotech, Uden, the Netherlands) according to the manufacturer’s protocols.

### CRP

High sensitivity CRP was measured using the standard assay by the Core Biochemical Assay Laboratory (CBAL) at Cambridge University Hospitals NHS Foundation Trust.

### Cytokines

IL-6, IL-10, IL-1β, TNFα and IFNγ were measured using standard clinical assays performed by the Clinical Immunology Laboratory at the Department of Biochemistry and Immunology, Addenbrooke’s Hospital Cambridge.

### IFNγ FLUOROSPOT assays

Frozen PBMCs were rapidly thawed, and the freezing medium was diluted into 10ml of TexMACS media (Miltenyi Biotech), centrifuged and resuspended in 10ml of fresh media with 10U /ml DNase (Benzonase, Merck-Millipore via Sigma-Aldrich), PBMCs were incubated at 37°C for 1h, followed by centrifugation and resuspension in fresh media supplemented with 5% Human AB serum (Sigma Aldrich) before being counted. PBMCs were stained with 2ul of each antibody: anti-CD3-fluorescein isothiocyanate (FITC), clone UCHT1; anti-CD4-phycoerythrin (PE), clone RPA-T4; anti-CD8a-peridinin-chlorophyll protein - cyanine 5.5 (PerCP Cy5.5), clone RPA-8a (all BioLegend, London, UK), LIVE/DEAD Fixable Far Red Dead Cell Stain Kit (Thermo Fisher Scientific). PBMC phenotyping was performed on the BD Accuri C6 flow cytometer. Data were analysed with FlowJo v10 (Becton Dickinson, Wokingham, UK). 1.5 to 2.5 x 10^5^ PBMCs were incubated in pre-coated Fluorospot plates (Human IFNγ FLUOROSPOT (Mabtech AB, Nacka Strand, Sweden)) in triplicate with peptide mixes specific for Spike, Nucleocapsid and Membrane proteins of SARS-CoV-2 (final peptide concentration 1µg/ml/peptide, Miltenyi Biotech) and an unstimulated and positive control mix (containing anti-CD3 (Mabtech AB), Staphylococcus Enterotoxin B (SEB), Phytohaemagglutinin (PHA) (all Sigma Aldrich)) at 37°C in a humidified CO_2_ atmosphere for 48 hours. The cells and medium were decanted from the plate and the assay developed following the manufacturer’s instructions. Developed plates were read using an AID iSpot reader (Oxford Biosystems, Oxford, UK) and counted using AID EliSpot v7 software (Autoimmun Diagnostika GmbH, Strasberg, Germany). All data were then corrected for background cytokine production and expressed as SFU/Million CD3 T cells.

### SARS-CoV-2 serology

Quantification of Spike SARS-CoV-2 specific antibodies was performed by ELISA as described by Xiong X et al (Xiong et al., 2020). Briefly, serum samples collected at time of enrolment in the study and at the 4-8 week follow-up visit were first screened for positivity and then antibody titres were determined by an end-point analysis. AUC values were calculated in R (3.6.3) using the flux (0.3-0) package. Kruskal–Wallis test was used to calculate p-values among the different disease severities.

### SARS-CoV-2 neutralisation assay

The virus used in this study was the clinical isolate SARS-CoV-2/human/Liverpool/REMRQ0001/2020, a kind gift from Ian Goodfellow (University of Cambridge), isolated by Lance Turtle (University of Liverpool) and David Matthews and Andrew Davidson (University of Bristol) (Daly et al., 2020; Patterson et al., 2020).

Sera were heat-inactivated at 56°C for 30 mins, then frozen in aliquots at −80°C. Neutralising antibody titres at 50% inhibition (NT50s) were measured as previously described (Pereyra Gerber, P. et al., under review).

In brief, HEK293T reporter cells expressing Renilla luciferase (Rluc) and SARS-CoV-2 Papain-like protease-activatable circularly permuted firefly luciferase (FFluc) were seeded in flat-bottomed 96-well plates. The next day, SARS-CoV-2 viral stock (MOI=1) was pre-incubated with a 3-fold dilution series of each serum for 2 h at 37°C, then added to the cells. After 24 h, cells were lysed in Dual-Glo Luciferase Buffer (Promega) diluted 1:1 with PBS and 1% NP-40. Lysates were transferred to white half-area 96-well plates, and infectious virus quantitated as the ratio of FFluc/Rluc activity measured using the Dual-Glo kit (Promega) according to the manufacturer’s instructions.

Experiments were conducted in duplicate. To obtain NT50s, FFluc/Rluc ratios were analysed using the Sigmoidal, 4PL, X is log(concentration) function in GraphPad Prism.

### Whole blood bulk RNA-Seq

Whole blood RNA was extracted from PAXgene Blood RNA tubes (BD Biosciences) of 188 COVID-19 patients at up to 2 time points and 42 healthy volunteers. RNA-Sequencing libraries were generated using the SMARTer® Stranded Total RNA-Seq v2 - Pico Input Mammalian kit (Takara) using 10ng RNA as input following the manufacturer’s protocol. Libraries were pooled together (n = 96) and sequenced using 75bp paired-end chemistry across 4 lanes of a Hiseq4000 instrument (Illumina) to achieve 10 million reads per sample. Read quality was assessed using FastQC v.0.11.8 (Babraham Bioinformatics, UK), and SMARTer adaptors trimmed and residual rRNA reads depleted in silico using Trim_galore v.0.6.4 (Babraham Bioinformatics, UK) and BBSplit (BBMap v.38.67(BBMap - Bushnell B. - sourceforge.net/projects/bbmap/)), respectively. Alignment was performed using HISAT2 v.2.1.0 (Kim et al., 2019) against the GRCh38 genome achieving a greater than 95% alignment rate. Count matrices were generated using featureCounts (Rsubreads package - (Liao et al., 2020) and stored as a DGEList object (EdgeR package (Robinson et al., 2010) for further analysis.

All downstream data handling was performed in R (R Core Team, 2015). Counts were filtered using filterByExpr (EdgeR) with a gene count threshold of 10 CPM and the minimum number of samples set at the size of the smallest disease group. Library counts were normalised using calcNormFactors (EdgeR) using the method ‘weighted trimmed mean of M-values’. The function ‘voom’ (Law et al., 2014) was applied to the data to estimate the mean-variance relationship, allowing adjustment for heteroscedasticity.

### Single cell RNA-seq

CITE-seq data were generated from frozen PBMCs as described by Stephenson et al. (Stephenson et al., 2021). Briefly, after thawing, pool of 4 samples were generated by combined 500,000 viable cells per individual (total of 2 million cells per pool). TotalSeq-C™ antibody cocktail (BioLegend 99813) was used to perform cell surface marker staining on 500,000 cells per pool. 50,000 live cells (up to a maximum of 60,000 total cells) for each pool were processed using Single Cell V(D)J 5’ version 1.1 (1000020) together with Single Cell 5’ Feature Barcode library kit (1000080), Single Cell V(D)J Enrichment Kit, Human B Cells (1000016) and Single Cell V(D)J Enrichment Kit, Human T Cells (1000005) (10xGenomics) according to the manufacturer’s protocols. Samples were sequenced on NovaSeq 6000 (Illumina) using S1 flowcells. Droplet libraries were processed using Cellranger v4.0. Reads were aligned to the GRCh38 human genome concatenated to the SARS-Cov-2 genome (NCBI SARS-CoV-2 isolate Wuhan-Hu-1) using STAR (Dobin et al., 2013) and unique molecular identifiers (UMIs) deduplicated. CITE-seq UMIs were counted for GEX and ADT libraries simultaneously to generate feature X droplet UMI count matrices.

### Statistics

All statistical analyses were conducted using custom scripts in R (R Core Team, 2015). Absolute cell counts (cells/uL) were offset by +1 to allow subsequent log2 transformation of zero counts. Where shown, time measures represent time from symptom onset (for severity groups B, C, D and E) or first positive COVID-19 swab (group A). Unless otherwise specified, longitudinally collected data was grouped by bins of 7 or 12 days. Pairwise statistical comparison of absolute cell counts, CRP or serum measures between individuals in a given severity group at a given time bin and HCs, or between severity groups, was conducted by Wilcoxon test unless otherwise specified. For analyses involving repeated measures, false discovery rate corrected (Benjamini & Hochberg) p value were reported. For individuals sampled more than once within a given time bin, data from the earliest blood collection was used.

Cell subset deconvolution of the whole blood RNA-Seq dataset was performed using pathway-level information extractor (PLIER) (http://gobie.csb.pitt.edu/PLIER). Latent factors were generated by leveraging off pre-existing knowledge of cell specific pathways. To better understand the relationship between gene expression and clinical severity, weighted gene co-expression network analysis was carried out using the WGCNA package (Langfelder and Horvath, 2008) in R. Briefly, a signed adjacency matrix was generated and a soft thresholding power was chosen to impose approximate scale-free topology. Modules were identified from the resulting topological overlap matrix with a specified minimum module size of n = 30. Modules were summarized using singular value decomposition, and the resulting module eigengene correlated with clinical traits. Significance of the correlation between a given clinical trait and a modular eigengene was assessed using linear regression with Bonferroni adjustment to correct for multiple testing. Modules were annotated using Enrichr (Chen et al., 2013). Longitudinal mixed modelling of gene module changes over time (*y*_*ij*_) was conducted using the *nlme* package in R (Pinheiro J et al., 2020), including time (*t*_*ij*_) with a quadratic trend and disease severity category or unsupervised cluster ids (*X*_*j*_) as fixed effects, and sampled individuals as random effects (*u*_*j*_):

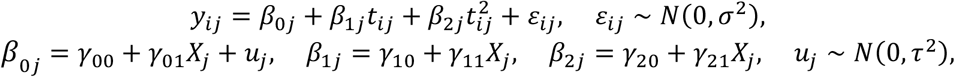

i.e., using the *lme* formula:

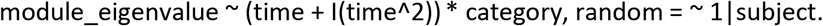

Gene set enrichment analysis (GSEA) (Subramanian et al., 2005) was used to identify biological pathways enriched in COVID-19 severity groups relative to healthy controls. Briefly, a list of ranked genes, determined by Signal-To-Noise ratio was generated. An enrichment score was calculated, determined by how often genes from the geneset of interest appeared at the top or the bottom of the pre-ranked set of genes with the enrichment score representing the maximum deviation from zero. To assess statistical significance, an empirical phenotype-based permutation test was run, where a collection of enrichment scores was generated from the random assignment of phenotype to samples and used to generate a null distribution. To account for multiple testing, an FDR rate q < 0.20 was deemed significant. A leading edge analysis was performed to determine the genes contributing the most to the enrichment of a given pathway and was subsequently illustrated in a heatmap. HALLMARK gene sets from the Molecular Signatures Database (http://www.broadinstitute.org/gsea/msigdb) were used in analysis.

Principal component analysis (PCA) of centred and scaled absolute counts for 24 major cell types was conducted using the *pca()*function from the package *mixOmics* (Rohart et al., 2017). Unsupervised clustering of log2 transformed absolute cell counts, normalised to the median of the corresponding control population, was conducted using the *heatmap*.*2()* function from the package *gplots* (Gregory R. Warnes et al., 2020), with a Euclidean distance function applied to both rows and columns of the data matrix and hierarchical clustering computed using the *ward*.*D* method. Partial least squares discriminant analysis (PLS-DA) was conducted using the *plsda()* function from the package *mixOmics* (Rohart et al., 2017), a supervised method of sample discrimination whereby sample clustering is informed by group membership (here patient clusters 1 and 2). The classification performance of the PLS-DA model was determined using the *perf()* function via 10 iterations of 5-fold cross-validation, with two components deemed sufficient to minimise the balanced error rate of prediction. Variable selection on components 1 and 2 was conducted using the *tune()* function, with 13 cell types selected as those most strongly contributing to discrimination of patient clusters. An AUROC curve showing the performance of a predictive model based on these 13 cell types was generated using the *auroc()* function. To assess whether clinical severity was reflected on a transcriptional level in an unsupervised fashion, K-means clustering was utilised on normalised whole blood RNASeq gene expression counts. Heat maps were created using the ComplexHeatmap package (Gu et al., 2016), with data scaled and centred prior to visualisation.

Cellular recovery rates over 60 days were calculated for each cell type in patients from groups C, D and E, split into those with persistently elevated (>10mg/L) or resolving CRP (falling below 10mg/L by final bleed), over 60 and 40 days respectively. Using a 12 day sliding window with single day increments, the ‘window of recovery’ for each given cell population was defined as the window in which absolute cell counts for COVID-19 samples no longer differed significantly from controls when assessed by Wilcoxon test, and remained as such for the subsequent 7 windows, and 80% of all windows remaining. Recovery rate was taken as the log2 normalised ratio of test and control absolute counts for patient samples collected within the first time window (0-12 days), subtracted from the equivalent value calculated within the window of recovery, divided by the upper day boundary of the recovery window.

The relationships between immunophenotyping and transcriptional data in the form of gene expression modules were assessed using Pearson’s correlation (Hmisc package) and visualized with corrplot.

## Supplementary Figure Legends

**Figure S1: Inflammation and viral load changes over time in COVID-19 patients – extended figure 1 A)** SARS-CoV-2 PCR CT values over time across patient severity groups. CT values ≤ 38 (dashed horizontal line) were reported as a positive result. A discretional CT number of 45 was assigned to samples with no detectable SARS-CoV-2 RNA. Each colour corresponds to a different subject. Repeat measures for a single participant are linked by dashed line. Y axis is inverted. **B)** Boxplots showing complement components and cytokine levels for samples collected within 12-day time bins. Grey band indicates the interquartile range of the corresponding measure in HCs. Points are coloured based on asymptomatic or symptomatic classification for categories A and B respectively, normal or abnormal chest radiology (group C), and level of respiratory support at sampling (group D and E). NIV/HF, non-invasive ventilation/high-flow oxygen; vent, mechanical ventilation; ECMO, extracorporeal membrane oxygenation.

**Figure S2: Comparison of absolute cell counts derived by flow and mass cytometry**

Scatter plots showing the correlation between cell populations quantified by both flow and mass cytometry (cells/uL). Pearson correlation R value and p-values of correlation test are reported for each comparison.

**Figure S3: Cellular changes over time in COVID-19 patients - extended figure 2 including full cell dataset**.

**A)** Heatmap showing the log2 fold change in median absolute cell count between COVID-19 patients and healthy controls, within severity categories and across 12-day time bins post screening (group A) or symptom onset (groups B-E). Missing data are shown in grey. Wilcoxon test FDR adjusted p-values: *<0.05, **<0.005, *** <0.0005. **B and C)** tSNE plots comparing groups A and B to groups C, D and E within 14 or later (>14) days post screening (group A) or symptom onset (groups B-E) for **B)** CyTOF and **C)** CITE-Seq dataset. Cell clusters are coloured as key provide.

**Figure S4: Whole blood transcriptomic signatures over time – extended figure 3**.

**A)** Annotation of Latent Factors used to perform the cell subset deconvolution shown in **B). B)** Cell subset deconvolution performed using PLIER, leveraging off prior knowledge of cell specific pathways. COVID-19 cases split by severity categories and 24-day time bins. Latent factor expression compared with HC, FDR adjusted p-value: *<0.05, **<0.005, ***<0.0005. **C)** Heatmap illustrating the correlation among whole blood co-expression gene modules derived from WGCNA (coloured blocks, y axis) and age, gender, steroid treatment, CRP levels, and the comparison between healthy controls and COVID-19 cases split by severity in 24 days bins (x axis). Pearson correlation and FDR corrected p-values are shown for each comparison. **D)** Annotation of modules that correlate with disease and/or severity. **E)** Correlation between SARS-CoV-2 PCR CT values and Interferon Signature Genes (IGS) module eigenvalues.

**Figure S5: Multivariate analysis of immune-cell populations in early disease - extended figure 4**

**A)** Principal component analysis of peripheral blood absolute cell counts for 24 key cell subsets from HC and COVID-19 cases, for samples taken <10 days from screening (group A) or symptom onset (groups B-E). Points are coloured according to severity category. **B)** K-means clustering of 18357 whole blood transcripts from COVID-19 samples taken <10 days from screening or symptom onset. Gene clusters are annotated for enriched signatures, samples are annotated according to corresponding cluster membership in Figure 4A where possible. **C)** Variable selection by sPLS-DA showing discrimination of patient clusters 1 and 2 derived in Figure 4A. **D)** Associated classification error rate of the predictive model across 10 iterations of 5-fold cross validation for components 1-10. E) Feature selection on components 1 and 2, determining 13 cell subsets as key contributors to cluster discrimination with minimal error. Unsupervised clustering of 13 selected cell types (normalised to the median of healthy controls), with original sample clusters and patient characteristics indicated. **F) and G)** AUROC curves showing sensitivity of cluster group prediction at varying specificity thresholds, based on **F)** absolute counts of 13 selected cell types, or **G)** CRP alone and combine with the absolute counts of 13 selected cell types.

**Figure S6: Early immune changes associated with mild disease and outcome – extended figure 5**

**A and B):** Boxplots showing **A)** non-naive HLA-DR+CD38+ CD8 T and **B)** plasmablast absolute cell counts (cells/uL) across severity categories at weeks 1-3 post screening (group A) or symptom onset (groups B-E). Wilcoxon test FDR adjusted p-value: *<0.05, **<0.005, ***<0.0005. Grey bar represents the interquartile range of the same cell populations in HCs. **C)** Enrichment score for CD8 T-cell activation signature as determined by GSEA in group B-E for samples taken <24 days from symptom onset. **D)** Spot forming unit (SFU) numbers of CD3 T cells secreting IFN g in response to membrane, nucleocapsid and spike SARS-CoV-2 antigen stimulation, in patient samples from groups B, and D and E combined, one or two weeks post symptom onset. Kruskal-Wallis test p-values. **E)** Area under the curve for SARS-CoV-2 spike-specific IgM and IgA titres at 1, 2 and 5 weeks post screening (group A) or symptom onset (group B-E). Groups C, D and E are combined for increased statistical power. Wilcoxon test p-values. **F)** Area under the curve for SARS-CoV-2 nucleocapsid-specific IgG, IgM and IgA titres. Groups, timepoints and p-values as in **E). G)** Total IgG and IgM levels across severity groups, within 3 weeks post screening (group A) or symptom onset (group B-E). Grey band corresponds to 5-95th centile ranges based on UK Caucasian population, as published in the Protein Reference Unit Handbook (9th Edition). P-values from comparisons with the reference range using Pearson’s chi-square test, annotated as in **A). H)** Boxplots capturing expression of interferon stimulated genes (ISG) and neutrophil activation-related transcriptomic eigenvalues across disease severity and time. **I)** Stratification of group E samples taken <24 days post symptom onset into high and low expression of ISG, with persisting and resolving CRP status and final respiratory status reported within 12 weeks shown by bar charts.

**Figure S7: Transcriptional changes in prolonged disease – extended figures 7A and C**

**A)** GSEA assessing HALLMARK oxidative phosphorylation geneset in different cell type identify by CITE-Seq against HC in COVID-19 patients collected within 14 days post screening (group A) or symptom onset (groups B-E). FDR adjusted p-value is shown by circle diameter, with colour representing normalised enrichment score. **B) and C):** Heatmap showing the correlation between gene expression eigenvalues derived from whole blood RNA-Seq, absolute cell counts and inflammatory characteristics in COVID-19 patients collected within **B)** the first 24 days, or **D)** between 25-48 days post screening or symptom onset. Pearson correlation p-values: *p<0.05, **p<0.01 and ***p<0.001.

### Item S2: Cell data

**A)** Boxplots showing absolute counts (cells/uL) for all the cell populations, split by severity categories and 12-day time bins post screening (group A) or symptom onset (group B-E). Points are coloured based on asymptomatic or symptomatic classification for categories A and B respectively, normal or abnormal chest radiology (group C), and level of respiratory support at time of sampling (groups D and E), as per the key colour provided. **B)** Mixed-effects model with quadratic time trend showing the longitudinal trajectories of all the cell populations over time, grouped by severity. Nominal and adjusted p-values for the time x severity group interaction term are reported. Grey band in **A)** and **B)** indicates interquartile range of the corresponding population in healthy controls. NIV/HF, non-invasive ventilation/high-flow oxygen; vent, mechanical ventilation; ECMO, extracorporeal membrane oxygenation.

### Item S3: Differences between patients in cluster 1 and cluster 2 as defined in figure 4A

**A)** Mixed-effect model with quadratic time trend showing the longitudinal trajectories of inflammatory markers, and boxplot at time of sampling (<=10 days post symptom onset), for individuals in clusters 1 and 2. **B)** Mixed-effect model with quadratic time trend showing the longitudinal trajectories for all cell populations over time, for COVID-19 cases in clusters 1 and 2. Grey bands in A) and B) indicate the interquartile range of the corresponding measurements in HCs. Nominal and adjusted p-values for the time x cluster interaction term are reported.

### Item S4: Flow cytometry gating strategy

**A)** B cell, **B)** non-conventional T cell, **C)** DCs and monocyte, **D)** T regulatory cells, and **E)** conventional T cell panels.

