## Supplementary Figures for "Delayed bystander CD8 T cell activation, early immune pathology and persistent dysregulation characterise severe COVID-19": Supplementary_figures_15032021_complete_reduce_v3.pdf

**Figure S1:** Inflammation and viral load changes over time in COVID-19 patients – extended Figure 1

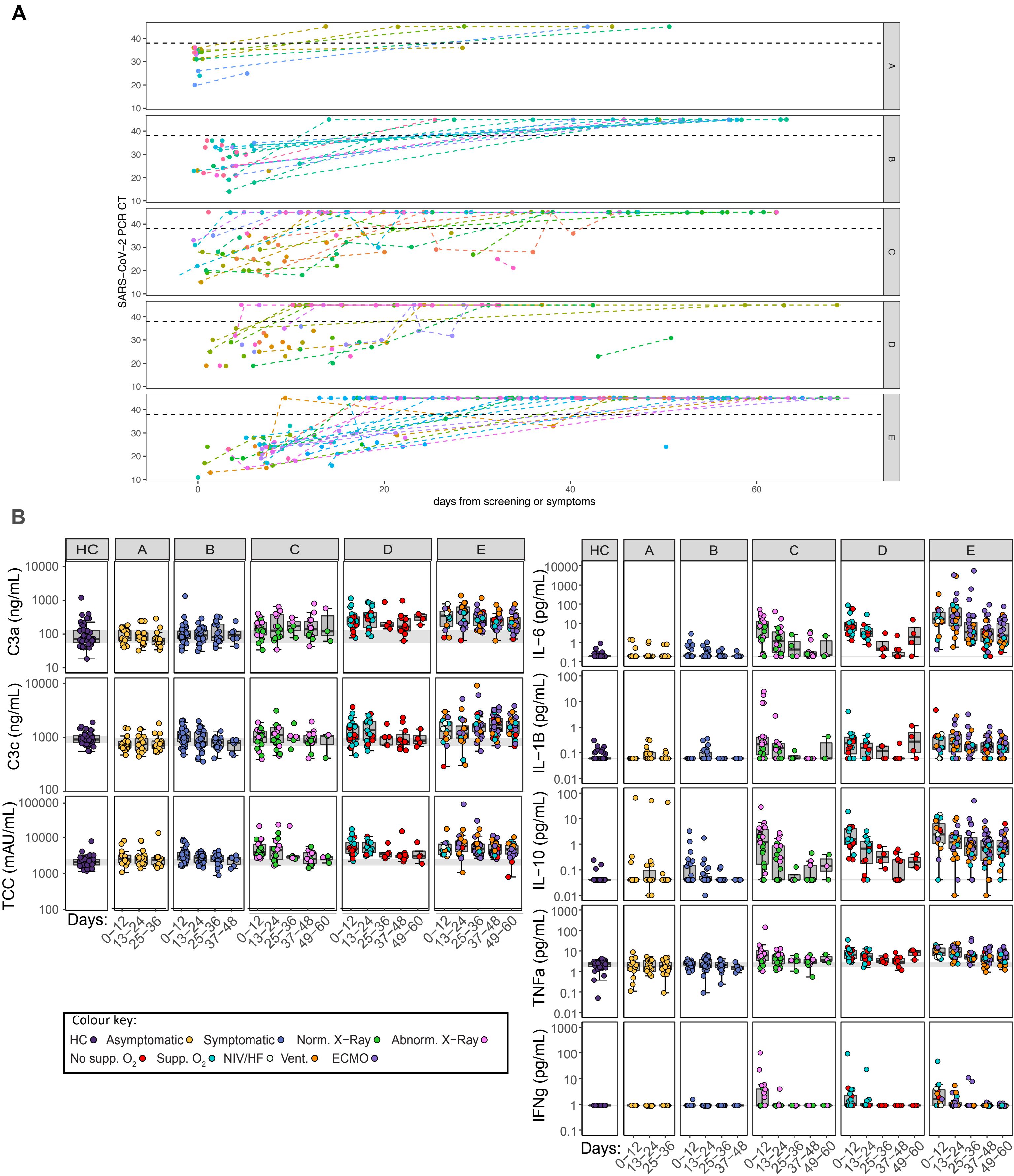

### **Figure S2:** Comparison of absolute cell counts derived by flow and mass cytometry

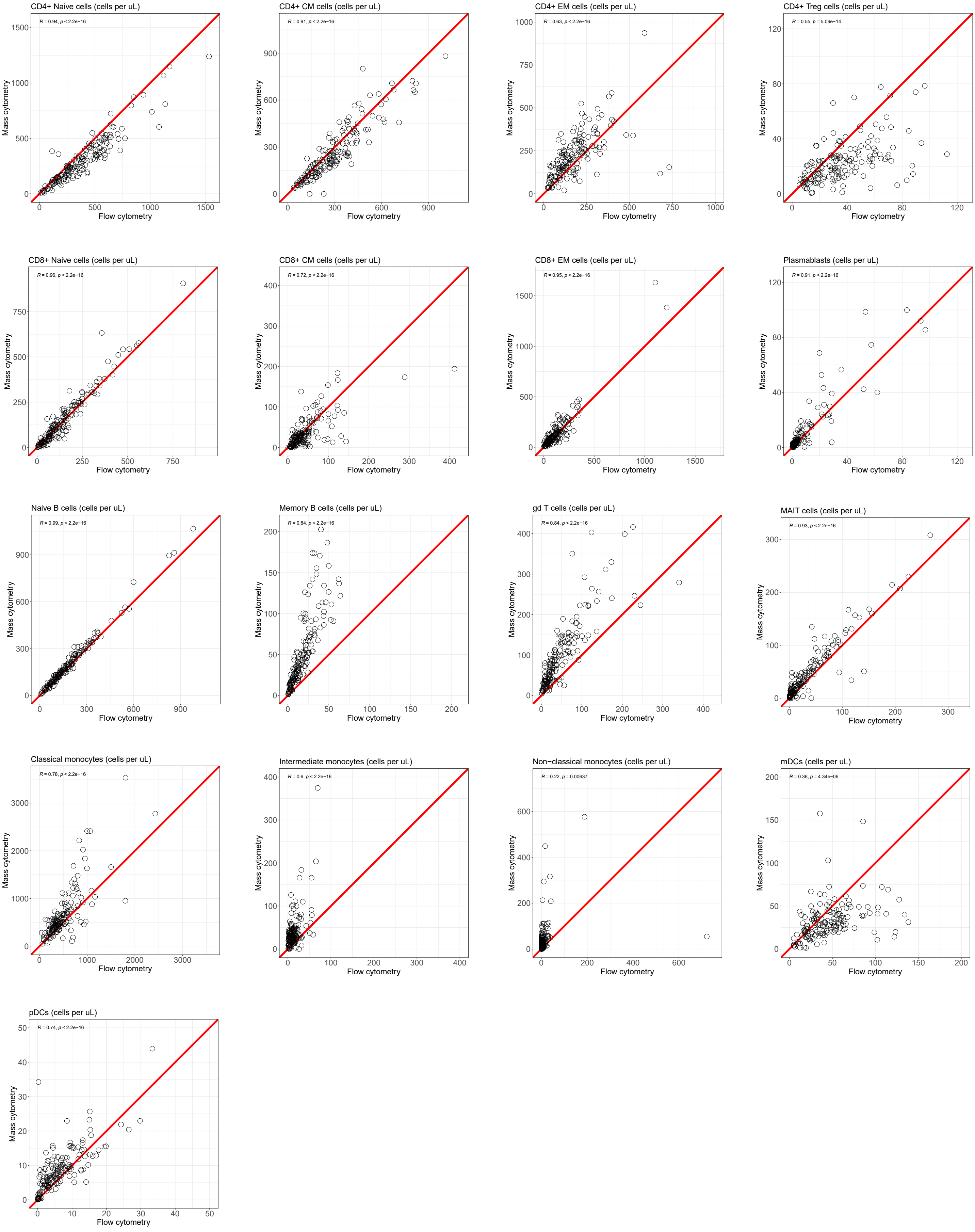

**Figure S3:** Cellular changes over time in COVID-19 patients – extended figure 2 including full cell dataset

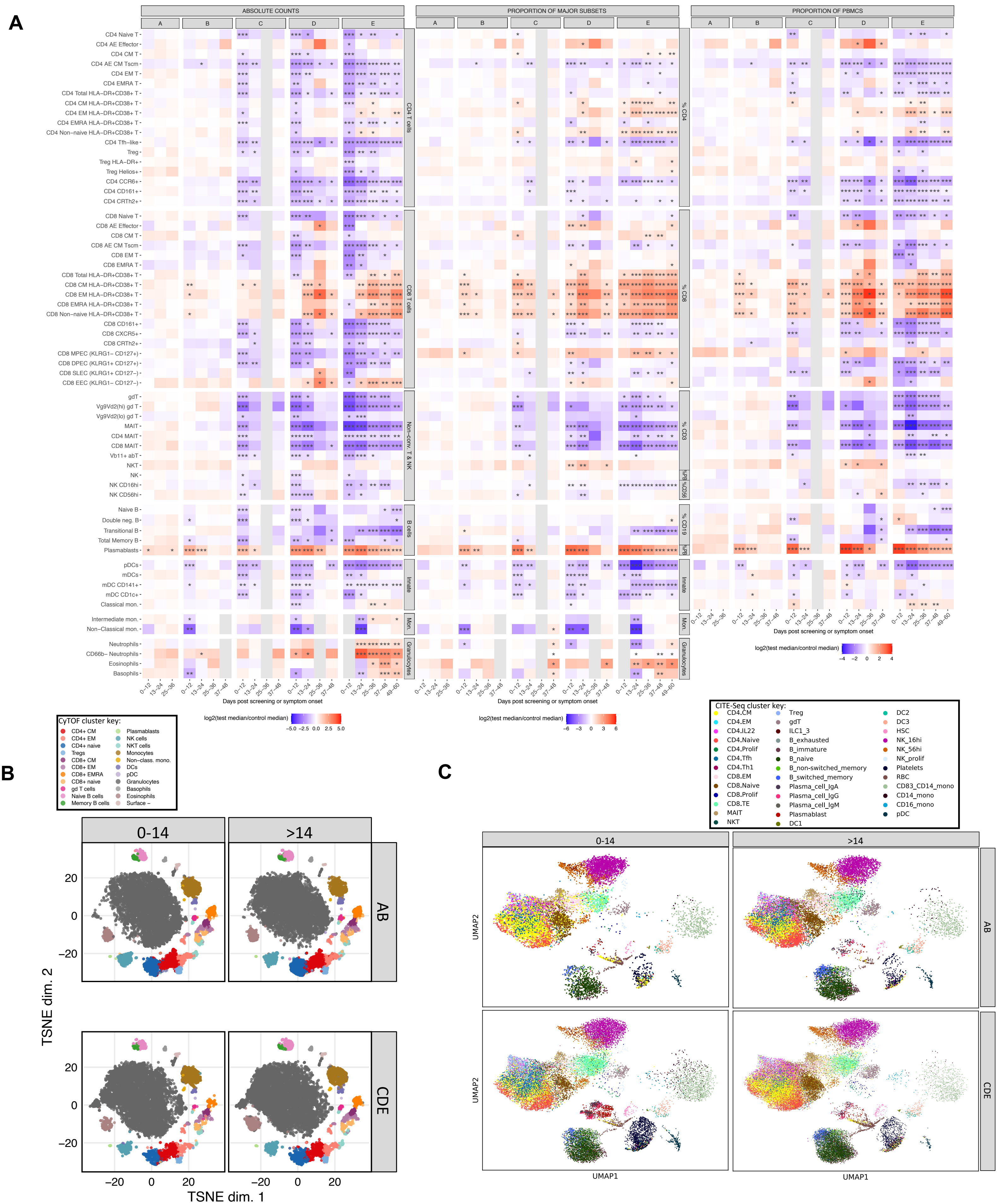

### Figure S4: Whole blood transcriptomic signatures over time – extended figure 3.

A

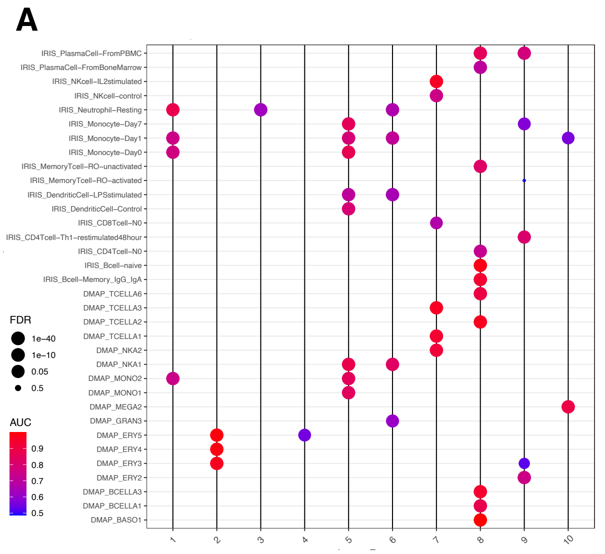


B

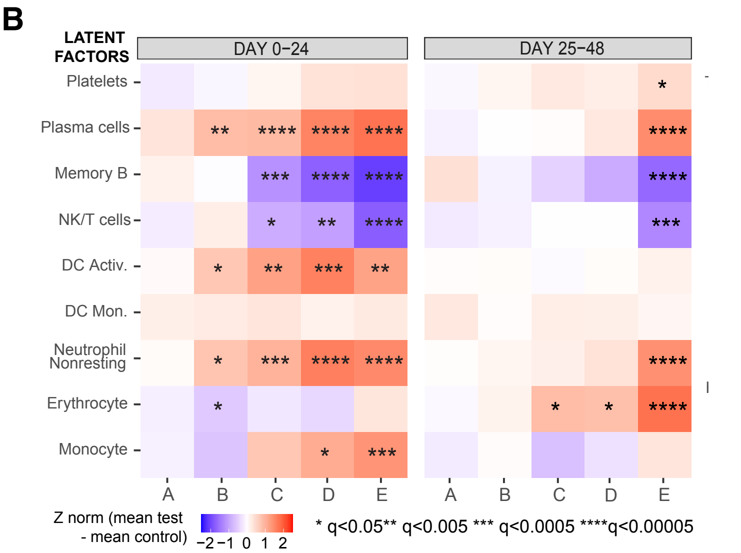


C

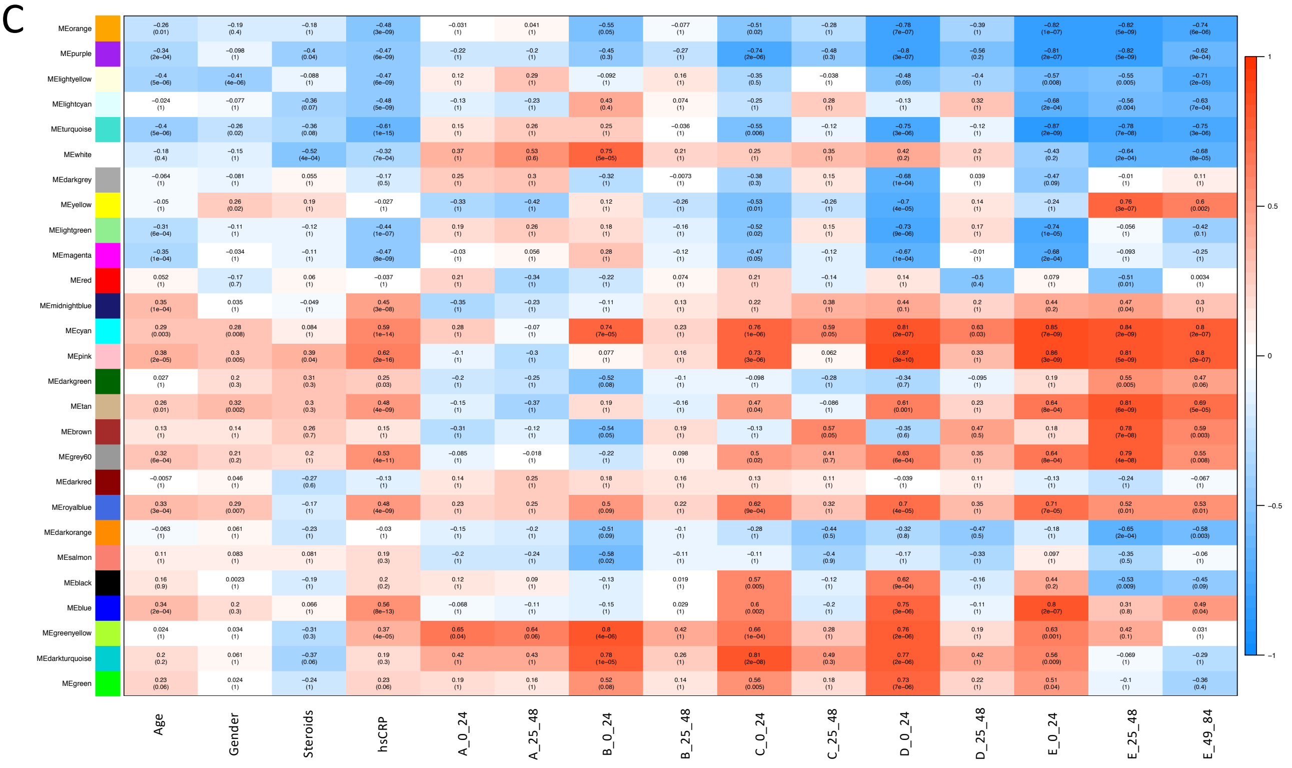


D

| Module | Annotation |
|---|---|
| **Positively correlated in all severity groups during active disease** | |
| MEcyan | Histones |
| MEblue | TNFa/IL-6 |
| MEpink | Complement/Coagulation/Neut degranulation |
| MEmidnightblue | Platelet activation |
| MEgrey60 | Ferroptosis |
| MEroyalblue | Glycolysis |
| **Positively correlated at all time points except late severe** | |
| MEgreenyellow | Immunoglobulins |
| MEgreen | Interferon Stimulated Genes |
| **Positively correlated in late mod/severe groups** | |
| MEbrown | Heme metabolism |
| **Positively correlated in late severe group** | |
| MEyellow | Oxidative Phosphorylation |
| **Negatively correlated with disease especially in early mod/severe disease** | |
| MEdarkgrey | GPCR |
| MElightgreen | Ribosomal proteins |
| MEmagenta | MYC targets |
| **Negatively correlated with disease especially in late severe group** | |
| MEturquoise | Gene transcription |
| MEpurple | Splicesome |
| MElightyellow | BCR signalling |
| MElightcyan | IL-2/NK |

E

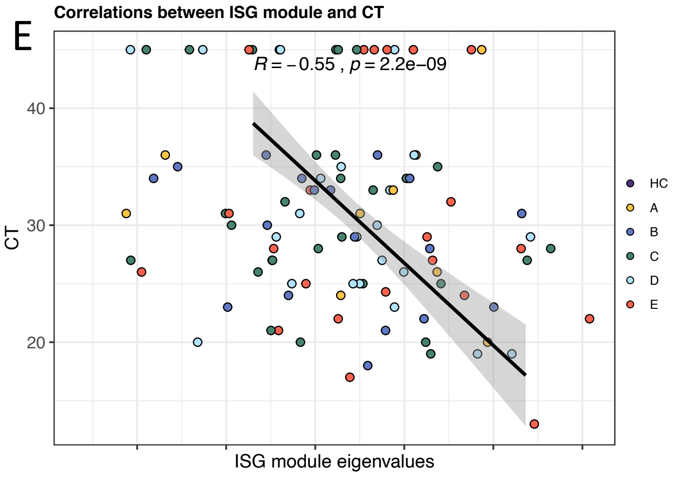

**Figure S5:** Multivariate analysis of immune-cell population in early disease – Extended figure 4.

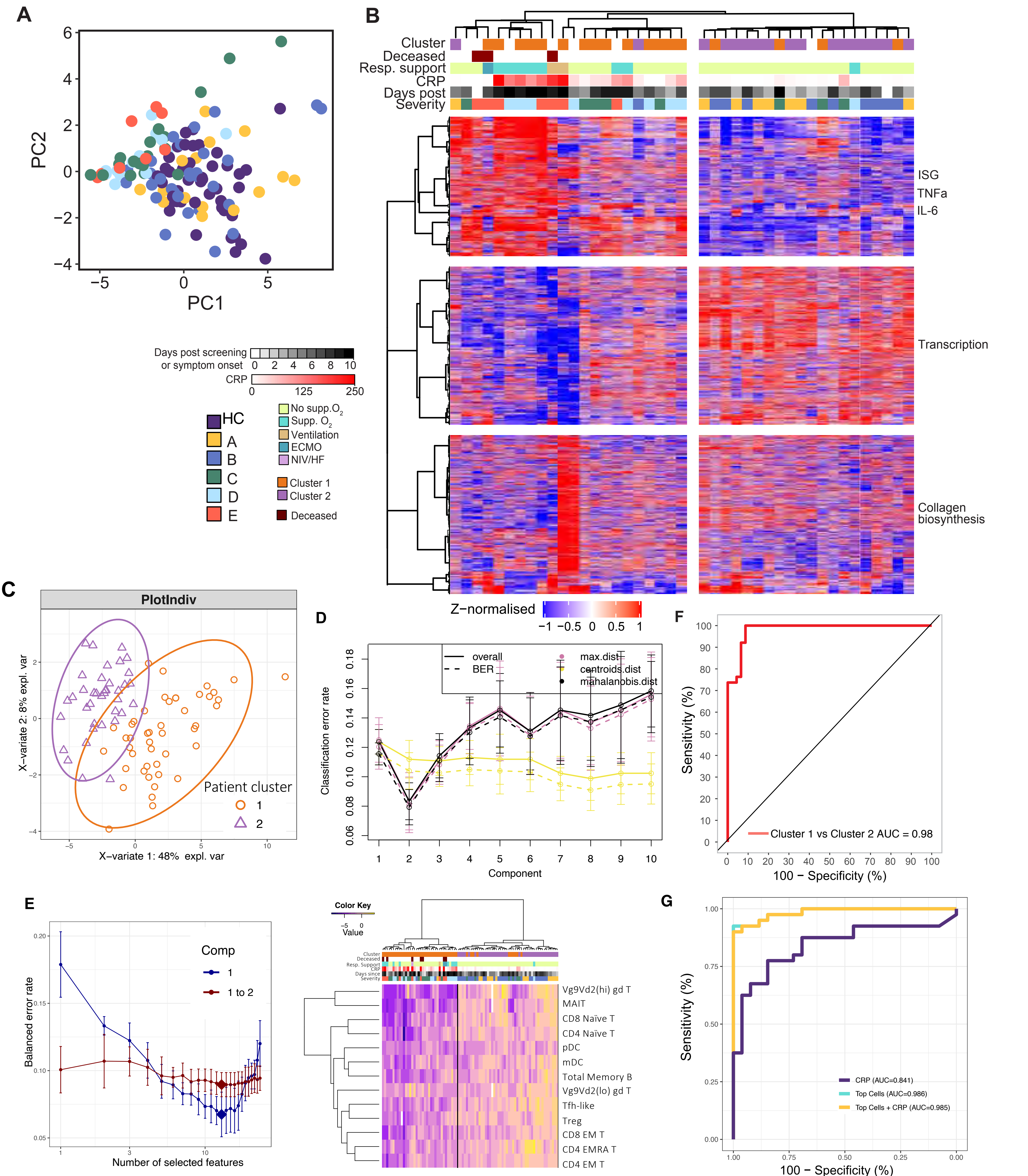

**Figure S6:** Early immune changes associated with mild disease and outcome – extended figure 5

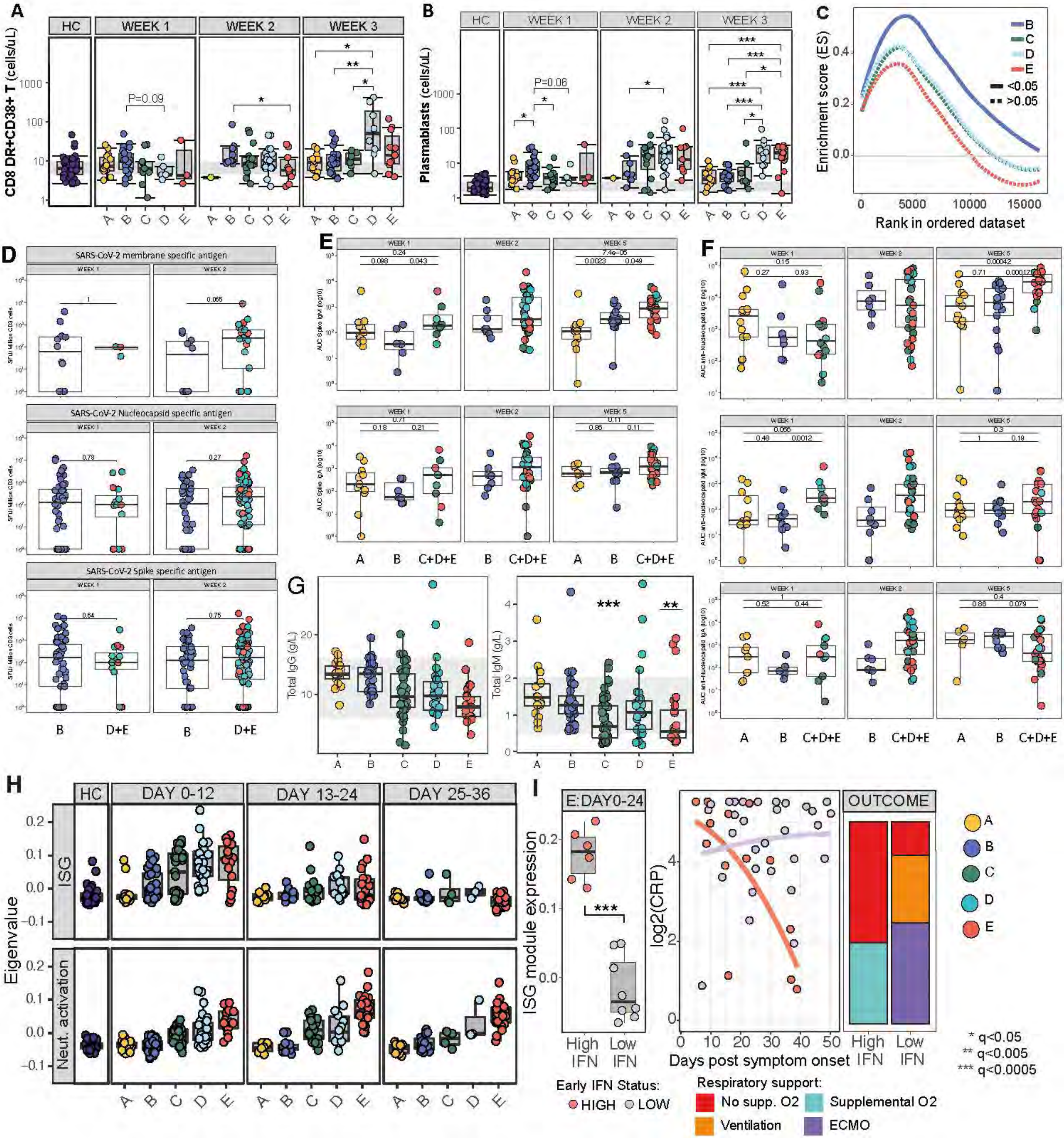

**Figure S7:** Transcriptional changes in prolonged disease – extended figures 7

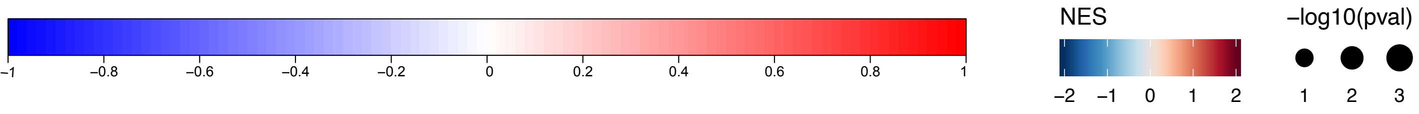


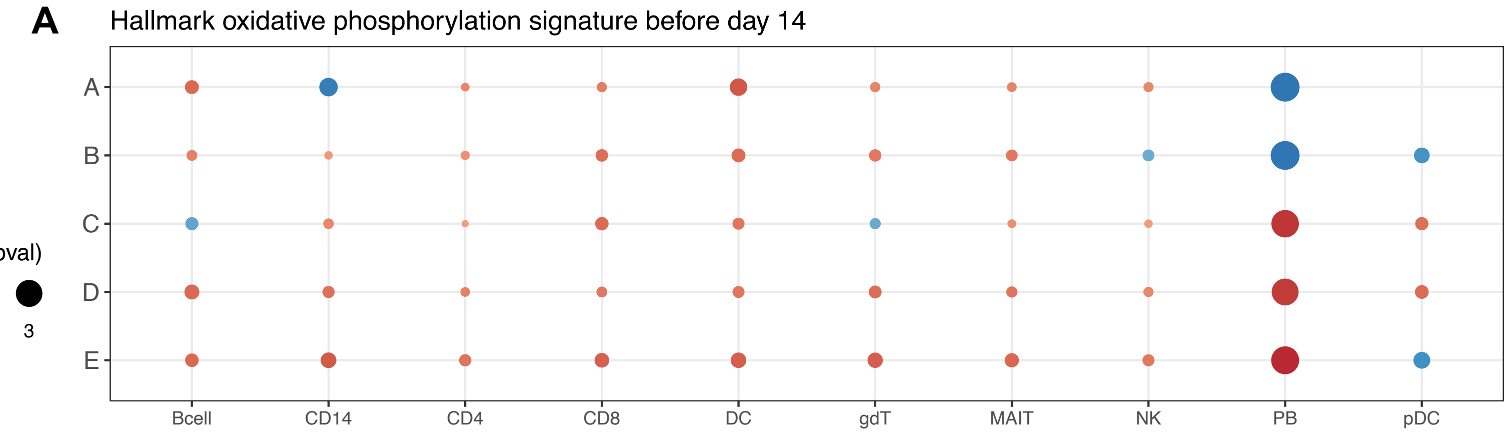


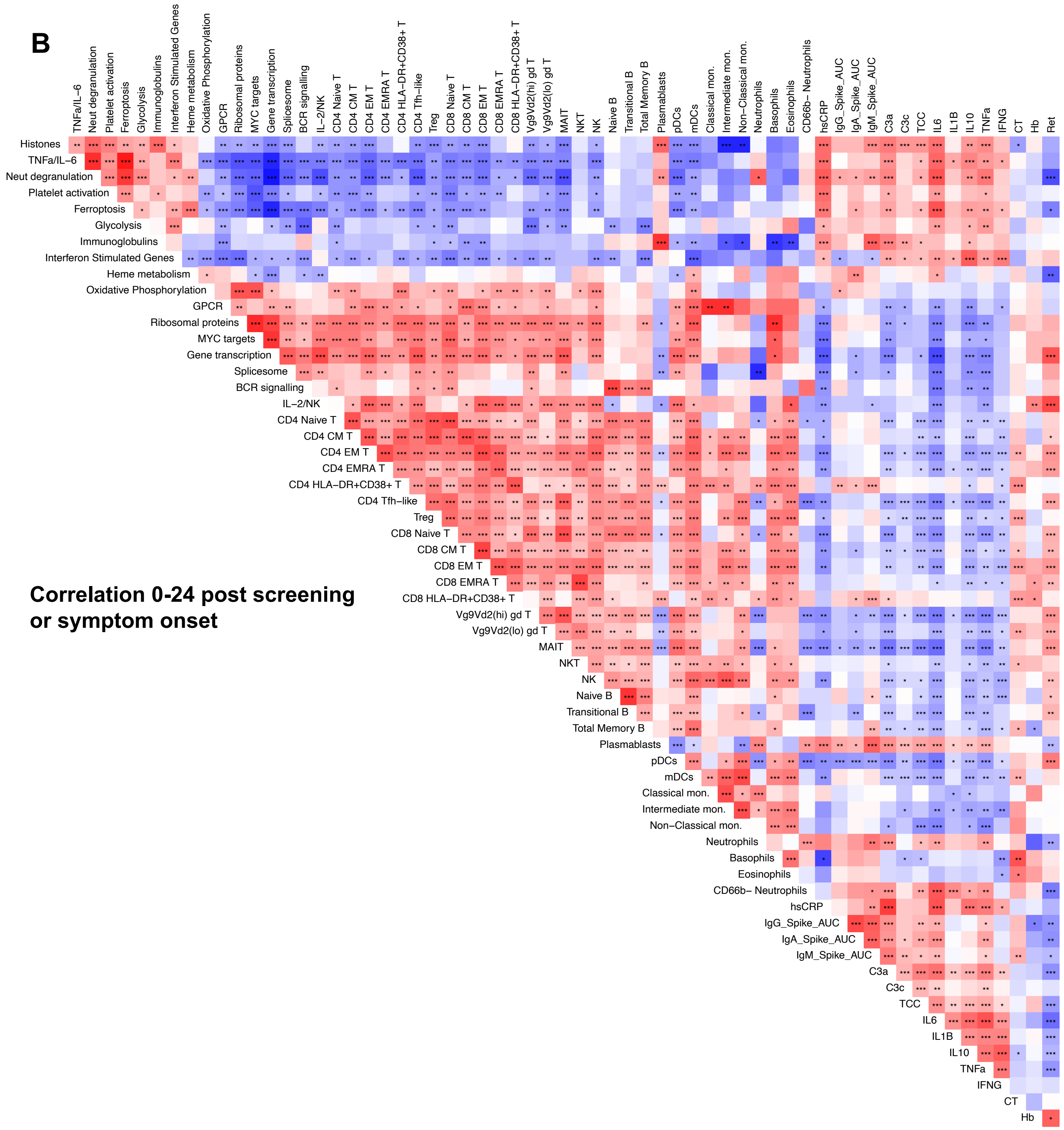


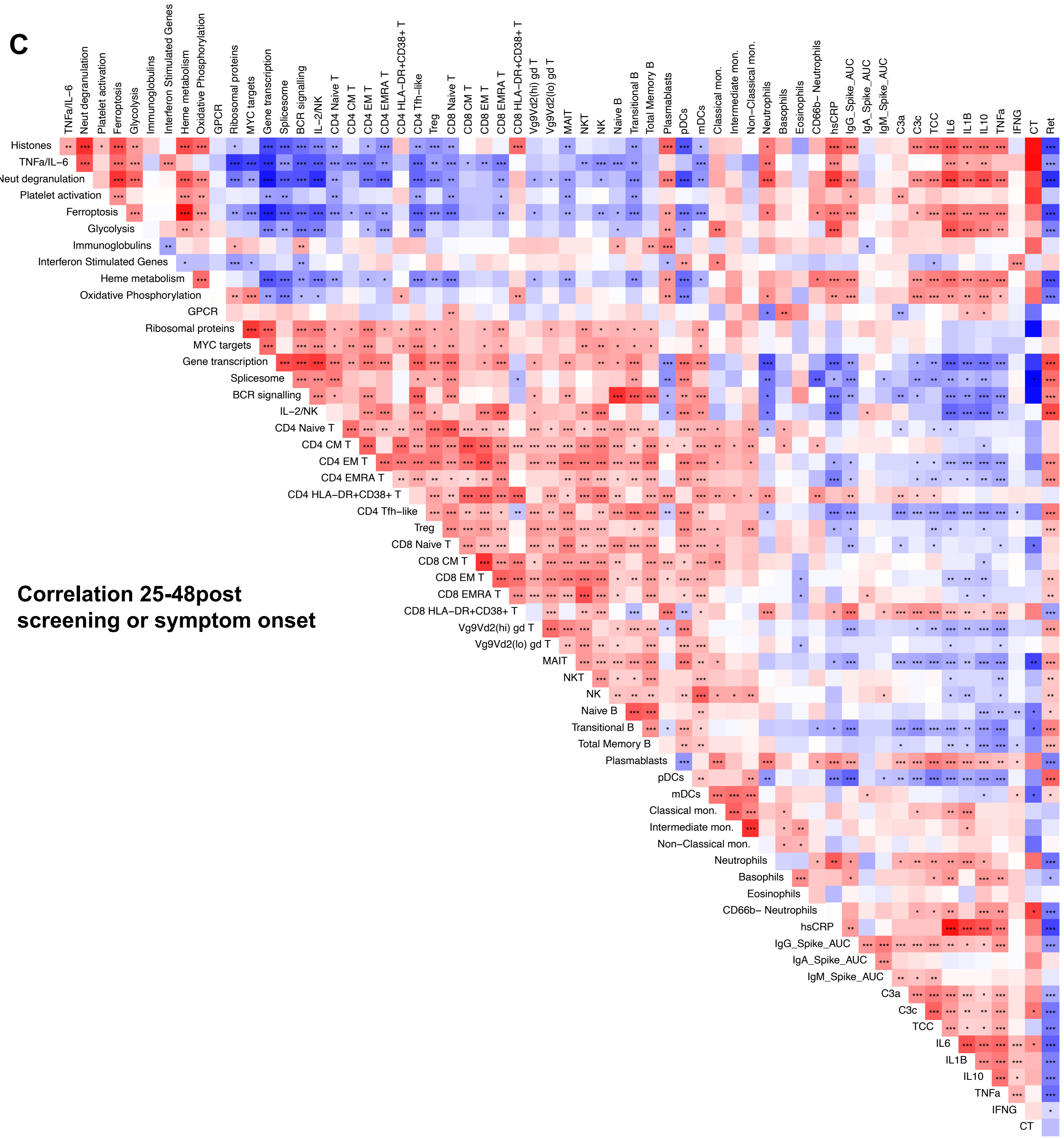
