## Supplementary Tables for "Delayed bystander CD8 T cell activation, early immune pathology and persistent dysregulation characterise severe COVID-19": COVID Pheno Paper Supplementary tables S1 S2 and S4 CURRENT.docx

Table S1: Clinical features of study participants, stratified by group A-E

|  | A | B | C | D | E |
| --- | --- | --- | --- | --- | --- |
| n | 18 | 40 | 46 | 37 | 60 |
| Gender (% male) | 22.2% | 22.5% | 54.3% | 64.9% | 75.0% |
| Age  (years, mean (SD)) | 32.9 (12.7) | 36.0  (11.8) | 58.0 (16.9) | 64.4  (15.1) | 57.0  (14.9) |
| Days from COVID-19 symptoms to enrollment (days, mean (SD)) | NA | 6.5  (2.9) | 11.4  (6.7) | 10.6  (8.1) | 24.6  (14.3) |
| COVID-19 chest radiology | NA | NA | 50.0% | 89.2% | 100% |
| Non-COVID19 admissions | NA | NA | 30.2% | 8.1% | 6.7% |
| Haemoglobin  (g/L, mean (SD)) | NA | NA | 124.8 (16.0) | 121.6 (18.0) | 95.2  (16.8) |
| Serum creatinine  (μmol/L, mean (SD)) | NA | NA | 82.9 (40.1) | 117.5 (154.7) | 103.5  (129.3) |
| Serum albumin  (g/L, mean (SD)) | NA | NA | 32.4  (7.1) | 28.0  (6.3) | 24.4  (7.2) |
| LOS  (days, median (IQR)) | NA | NA | 4  (1.25-10) | 10  (6-16) | 44  (33.7-63.2) |
| Admitted to ITU | NA | NA | 0% | 13.5% | 90.0% |
| Deceased in hospital | NA | NA | 2.2% | 0.0% | 30.0% |
| Hypertension | NA | NA | 47.8% | 43.2% | 48.3% |
| CAD | NA | NA | 8.7% | 24.3% | 16.7% |
| Other heart condition | NA | NA | 10.9% | 18.9% | 13.3% |
| Diabetes mellitus | NA | NA | 26.1% | 29.7% | 43.3% |
| CKD | NA | NA | 8.7% | 16.2% | 8.3% |
| PVD | NA | NA | 6.5% | 8.1% | 8.3% |
| CVA/TIA | NA | NA | 10.9% | 2.7% | 6.7% |
| COPD | NA | NA | 6.5% | 18.9% | 5.0% |
| Asthma | NA | NA | 21.7% | 10.8% | 10.0% |
| Other lung disease | NA | NA | 10.9% | 16.2% | 10.0% |
| Cancer | NA | NA | 4.4% | 5.4% | 1.7% |
| Haematological cancer | NA | NA | 2.2% | 5.4% | 0.0% |
| Corticosteroids | NA | NA | 19.6% | 10.8% | 10.0% |
| Immunosuppressive treatment | NA | NA | 17.4% | 16.2% | 5.0% |

SD is standard deviation, and IQR is interquartile range.

*COVID-19 chest radiology*: chest X-ray/ CT scan showed changes compatible with COVID-19, as opposed to normal findings or lung changes diagnostic of other conditions.

*Non-COVID19 admissions*: cases where COVID-19 was diagnosed during the hospital stay in patients initially admitted to hospital for reasons unrelated to COVID-19

*Haemoglobin, serum albumin and serum creatinine*: results from routine lab tests on the day of study enrollment, or closest result up to 2 days before. The included test results are available for at least for 75% of each severity group.

*LOS*: length of hospital stay (days from hospital admission to discharge, transfer or death in hospital)

*Hypertension*: history of hypertension, defined as blood pressure ≥140/80 on multiple occasions, or on treatment with any medication explicitly employed to reduce blood pressure

*CAD*: history of coronary artery disease, defined as myocardial infarction, angina, coronary artery stenting or coronary artery bypass grafting

*Other heart condition*: history of any other chronic cardiac disease (not CAD/hypertension), e.g. heart failure, congenital heart disease, cardiomyopathy, rheumatic heart disease

*CKD*: history of chronic kidney disease, defined as any of estimated glomerular filtration rate < 60 mL/min/1.73m^2^, dialysis or kidney transplant

*PVD*: history of peripheral vascular disease, defined as intermittent claudication or past bypass for chronic arterial insufficiency, history of gangrene or acute arterial insufficiency, or thoracic/abdominal aneurysm (≥6 cm)

*CVA/TIA*: history of a cerebrovascular accident or transient ischemic attacks

*COPD*: history of chronic obstructive pulmonary disease

*Other lung disease*: history of other chronic pulmonary disease (non asthma/COPD), e.g. cystic fibrosis, bronchiectasis, interstitial lung disease

*Cancer*: current solid organ malignancy (active or in the last 5 years), except non-melanoma skin cancers

*Corticosteroids*: history of treatment with systemic corticosteroids in the 14 days prior to hospital admission/presentation

*Immunosuppressive treatment*: history of treatment with immunosuppressants (excluding corticosteroids) in the 14 days prior to hospital admission/presentation, or chemotherapy/biologic drugs in the previous 6 months

Table S2: patients excluded because of extreme confounding comorbidities

| Study ID | Patient profile |
| --- | --- |
| CV0266 | Metastatic lung adenocarcinoma on immunotherapy and chemotherapy, presentation with new onset heart failure and pulmonary oedema, borderline positive COVID-19 PCR on nasopharyngeal swab, but no other clinical features of COVID-19 |
| CV0258 | Prolonged hospital admission for new diagnosis of acute myeloid leukemia with suspected leukemic lung infiltration and fungal chest infection, commenced on chemotherapy. COVID-19 PCR negative at hospital admission and subsequently positive. |
| CV0143 | Mild COVID-19 symptoms, admitted for extensive necrotizing fasciitis/mediastinitis, treated with surgical debridement and complicated by massive haemorrhage. Enrolled in the study in the ITU after surgery. |
| CV0192 | Emergency splenectomy following trauma. Fever and pneumonia in the post-op, with radiology in keeping with aspiration. Borderline positive COVID-19 PCR on nasopharyngeal swab. |
| CV0033 | Lymphoma on palliative chemotherapy, initially admitted for interstitial pneumonia and neutropenia and treated as Pneumocystis pneumonia. COVID-19 PCR initially negative 4x and subsequently positive after 2 weeks. |
| CV0313 | End-stage alcoholic cirrhosis with variceal bleeding and hepatic encephalopathy, admitted for transjugular intrahepatic portosystemic shunt procedure. Positive COVID-19 PCR on nasopharyngeal swab, initially with normal chest X-ray. Subsequently developed severe chest infection with bronchoalveolar lavage positive for Gram negative organisms and negative for COVID-19 PCR. |

Table S3: WGCNA module gene list and heme GSEA intersect genes (shared between C/D/E in the leading edge)

Csv file

Table S4: **Antibody staining panel used for flow cytometry (FACS) analysis.** Aliquots of PBMCs were stained with one of two antibody combinations prior to FACS analysis. The laser and band pass filter combination used to detect each antibody is also shown.

| **Panel** | **Laser** | **BPF** | **Antigen** | **Fluorophore** | **Clone** | **Vendor** | **Cat** |
| --- | --- | --- | --- | --- | --- | --- | --- |
| **DCs and monocytes** | 355 | 379/28 | CD40 | BUV395 | 5C3 | BD | 565202 |
|  |  | 515/30 | CD16 | BUV496 | 3G8 | BD | 612944 |
|  |  | 560/40 |  |  |  |  |  |
|  |  | 670/30 | CD3 | BUV661 | UCHT1 | BD | 612964 |
|  |  | 740/35 | CD86 | BUV737 | FUN-1 | BD | 612784 |
|  |  | 820/60 | CD45 |  BUV805 | HI30 | BD | 612891 |
|  | 405 | 450/50 |  |  |  |  |  |
|  |  | 515/20 | Zombie Aqua | |  | BioLegend | 423101 |
|  |  | 585/15 | CD45RA | BV570 | HI100 | BioLegend | 304132 |
|  |  | 605/40 | CD141 | BV605 | 1A4 | BD | 740421 |
|  |  | 670/30 | CCR7 | BV650 | G043H7 | BioLegend | 353234 |
|  |  | 710/40 | CD14 | BV711 | MφP9 | BD | 563372 |
|  |  | 741/40 |  |  |  |  |  |
|  |  | 780/60 | CD123 | BV786 | 7G3 | BD | 564196 |
|  | 488 | 525/50 | CD56 | FITC | MEM188 | Thermo | MHCD5601 |
|  |  | 610/20 |  |  |  |  |  |
|  |  | 685/35 |  |  |  |  |  |
|  |  | 715/30 | CD32 | BB700 | FLI8.26 | BD | 742216 |
|  |  | 780/60 |  |  |  |  |  |
|  | 561 | 585/15 | CD304 | PE | U21-1283 | BD | 565951 |
|  |  | 610/20 | CD163 | PE-CF594 | GHI/61 | BD | 562670 |
|  |  | 670/30 | CD80 | PECy5 | L307.4 | BD | 559370 |
|  |  | 710/40 |  |  |  |  |  |
|  |  | 780/60 | CD303 | PE-Vio770 | REA693 | Miltenyi | 130-110-321 |
|  | 640 | 670/30 | CD1c | AF647 | F10/21A3 | BD | 565048 |
|  |  | 730/45 | CD11c | AF700 | B-ly6 | BD | 561352 |
|  |  | 780/60 | HLA-DR | APC-H7 | G46-6 | BD | 561358 |
| **conventional T cells** | 355 | 379/28 | CD3 | BUV395 | SK7 | BD | 564001 |
|  |  | 515/30 | CD4 | BUV496 | SK3 | BD | 612936 |
|  |  | 560/40 |  |  |  |  |  |
|  |  | 670/30 | CD38 | BUV661 | HIT2 | BD | 612969 |
|  |  | 740/35 | CD95 | BUV737 | DX2 | BD | 612790 |
|  |  | 820/60 | CD45RA | BUV805 | HI100 | BD | 742020 |
|  | 405 | 450/50 | PD-1 | BV421 | EH12.2H7 | BioLegend | 329920 |
|  |  | 515/20 | CD8b | BV480 | 2ST8.5H7 | BD | 746266 |
|  |  | 585/15 | Zombie Yellow | |  | BioLegend | 423103 |
|  |  | 605/40 | HLADR | BV605 | L243 | BioLegend | 307640 |
|  |  | 670/30 | CD69 | BV650 | FN50 | BioLegend | 310934 |
|  |  | 710/40 | CD27 | BV711 | O323 | BioLegend | 302834 |
|  |  | 741/40 |  |  |  |  |  |
|  |  | 780/60 | CD28 | BV785 | CD28.2 | BioLegend | 302950 |
|  | 488 | 525/50 | KLRG1 | FITC | REA261 | Miltenyi | 130-103-640 |
|  |  | 610/20 |  |  |  |  |  |
|  |  | 685/35 |  |  |  |  |  |
|  |  | 715/30 |  |  |  |  | |
|  |  | 780/60 |  |  |  |  |  |
|  | 561 | 585/15 | CD161 | PE | HP-3G10 | BioLegend | 339904 |
|  |  | 610/20 | CRTh2 | PE-dazzle | BM16 | BioLegend | 350126 |
|  |  | 670/30 | CD127 | PECy5 | eBioRDR5 | Thermo | 15-1278-42 |
|  |  | 710/40 |  |  |  |  |  |
|  |  | 780/60 | CCR6 | PECy7 | G034E3 | BioLegend | 353418 |
|  | 640 | 670/30 |  |  |  |  |  |
|  |  | 730/45 | CXCR5 | APC-R700 | RF8B2 | BD | 565191 |
|  |  | 780/60 | CCR7 | APC-fire | G043H7 | BioLegend | 353246 |
| **non-conventional T cells.** | 355 | 379/28 | CD3 | BUV395 | SK7 | BD | 564001 |
|  |  | 515/30 | CD4 | BUV496 | SK3 | BD | 612936 |
|  |  | 560/40 |  |  |  |  |  |
|  |  | 670/30 |  |  |  |  |  |
|  |  | 740/35 | TCRgd | BUV737 | 11F2 | BD | 748533 |
|  |  | 820/60 | CD28 | BUV805 | L293 | BD | 748474 |
|  | 405 | 450/50 | PD-1 | BV421 | EH12.2H7 | BioLegend | 329920 |
|  |  | 515/20 | CD8b | BV480 | 2ST8.5H7 | BD | 746266 |
|  |  | 585/15 | Zombie Yellow | |  |  |  |
|  |  | 605/40 | HLADR | BV605 | L243 | BioLegend | 307640 |
|  |  | 670/30 | CD69 | BV650 | FN50 | BioLegend | 310934 |
|  |  | 710/40 | TCRV7.2 | BV711 | 3C10 | BioLegend | 351732 |
|  |  | 741/40 |  |  |  |  |  |
|  |  | 780/60 |  |  |  |  |  |
|  | 488 | 525/50 | KLRG1 | FITC | REA261 | Miltenyi | 130-103-640 |
|  |  | 610/20 |  |  |  |  |  |
|  |  | 685/35 |  |  |  |  |  |
|  |  | 715/30 | TCR-DV2 | PerCPCy5.5 | B6 | BioLegend | 331424 |
|  |  | 780/60 |  |  |  |  |  |
|  | 561 | 585/15 | CD161 | PE | HP-3G10 | BioLegend |  |
|  |  | 610/20 |  |  |  |  |  |
|  |  | 670/30 | CD127 | PECy5 | eBioRDR5 | Thermo | 15-1278-42 |
|  |  | 710/40 |  |  |  |  |  |
|  |  | 780/60 | TCR-DV1 | PECy7 | TS8.2 | Thermo | 25-5679-42 |
|  | 640 | 670/30 | CCR5 | AF647 | HEK/1/85a | Biolegend | 313712 |
|  |  | 730/45 | TCR Vγ9 | AF700 | B3 | Biolegend | 331318 |
|  |  | 780/60 | Vb11 | APCVio770 | REA559 | Miltenyi | 130-108-735 |
| B cells | 355 | 379/28 | IgM | BUV395 | G20-127 | BD |  |
|  |  | 515/30 | CD19 | BUV496 | SJ25C1 | BD |  |
|  |  | 560/40 |  |  |  |  |  |
|  |  | 670/30 | CD38 | BUV661 | HIT2 | BD |  |
|  |  | 740/35 | IgD | BUV737 | IA6-2 | BD |  |
|  |  | 820/60 | CD20 | BUV805 | 2H7 | BD |  |
|  | 405 | 450/50 | CD39 | BV421 | A1 | BioLegend |  |
|  |  | 515/20 | CD3 | BV510 | UCHT1 | BioLegend |  |
|  |  | 515/20 | CD14 | BV510 | 63D3 | BioLegend |  |
|  |  | 515/20 | CD15 | BV510 | W6D3 | BioLegend |  |
|  |  | 515/20 | CD193 | BV510 | 5.00E+08 | BioLegend |  |
|  |  | 585/15 | Zombie Yellow | |  |  |  |
|  |  | 605/40 | HLADR | BV605 | L243 | BioLegend |  |
|  |  | 670/30 | CD71 | BV650 | CY1G4 | BioLegend |  |
|  |  | 710/40 | CD27 | BV711 | O323 | BioLegend |  |
|  |  | 741/40 |  |  |  |  |  |
|  |  | 780/60 | CD73 | BV785 | AD2 | BioLegend |  |
|  | 488 | 525/50 | CD56 | FITC | MEM188 | Thermo |  |
|  |  | 610/20 |  |  |  |  |  |
|  |  | 685/35 |  |  |  |  |  |
|  |  | 715/30 | CD24 | BB700 | ML5 | BD |  |
|  |  | 780/60 |  |  |  |  |  |
|  | 561 | 585/15 | IgA | PE | Polyclonal goat IgG | Jackson |  |
|  |  | 610/20 | CD138 | PE-dazzle | DL-101 | BioLegend |  |
|  |  | 670/30 | CD80 | PECy5 | L307.4 | BD |  |
|  |  | 710/40 |  |  |  |  |  |
|  |  | 780/60 | CD86 | PECy7 | BU63 | BioLegend |  |
|  | 640 | 670/30 | GLUT1 | AF647 | 202915 | BD |  |
|  |  | 730/45 | CXCR5 | APC-R700 | RF8B2 | BD |  |
|  |  | 780/60 | IgG | APC-H7 | G18-145 | BD |  |
| T regulatory cells. | 355 | 379/28 | CD3 | BUV395 | SK7 | BD | 564001 |
|  |  | 515/30 | CD4 | BUV496 | SK3 | BD | 612936 |
|  |  | 560/40 |  |  |  |  |  |
|  |  | 670/30 |  |  |  |  |  |
|  |  | 740/35 |  |  |  |  |  |
|  |  | 820/60 | CD45RA | BUV805 | HI100 | BD | 742020 |
|  | 405 | 450/50 | PD-1 | BV421 | EH12.2H7 | BioLegend | 329920 |
|  |  | 515/20 | CD8b | BV480 | 2ST8.5H7 | BD | 746266 |
|  |  | 585/15 | Zombie Yellow | |  |  |  |
|  |  | 605/40 | HLADR | BV605 | L243 | BioLegend | 307640 |
|  |  | 670/30 | CCR7 | BV650 | G043H7 | BioLegend | 353234 |
|  |  | 710/40 |  |  |  |  |  |
|  |  | 741/40 |  |  |  |  |  |
|  |  | 780/60 | CD73 | Brilliant Violet 785™ | AD2 | BioLegend | 344028 |
|  | 488 | 525/50 |  |  |  |  |  |
|  |  | 610/20 |  |  |  |  |  |
|  |  | 685/35 |  |  |  |  |  |
|  |  | 715/30 | CD127 | PerCP efluor710 | eBioRDR5 | Thermo | 46-1278-42 |
|  |  | 780/60 |  |  |  |  |  |
|  | 561 | 585/15 | CD25 | PE | BC96 | Thermo | 12-0259-42 |
|  |  | 610/20 | Helios | Pedazzle | 22F6 | BioLegend | 137232 |
|  |  | 670/30 |  |  |  |  |  |
|  |  | 710/40 |  |  |  |  |  |
|  |  | 780/60 | CCR4 | PEVio770 | REA279 | Miltenyi | 130-103-814 |
|  | 640 | 670/30 | FoxP3 | APC | 236A/E7 | Thermo | 17-4777-42 |
|  |  | 730/45 | CXCR5 | APC-R700 | RF8B2 | BD | 565191 |
|  |  | 780/60 | CD39 | APC-fire | A1 | BioLegend | 328230 |
