## Supplementary Item 1 for "Delayed bystander CD8 T cell activation, early immune pathology and persistent dysregulation characterise severe COVID-19": Item_S1_IMMUNITY_11032021.pdf

**Time course plots for hospitalized study patients (groups C/D/E)**

#### Legend

no supplemental oxygen

supplemental oxygen

NIV/HF

invasive ventilation

ECMO

research bloods

asymptomatic

admitted to other hospital

transferred to other hospital

deceased

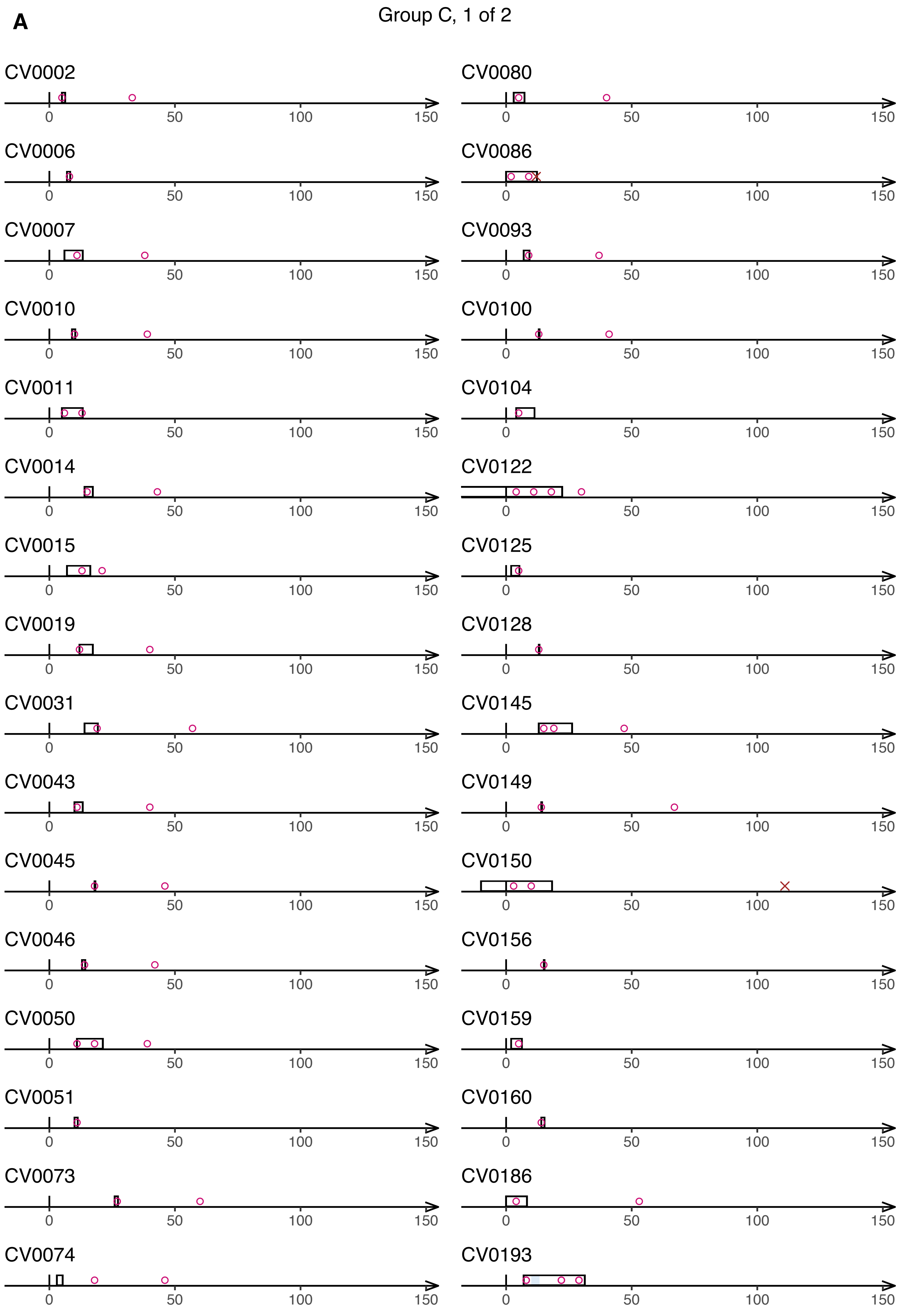
A
Group C, 1 of 2
CV0002
CV0006
CV0007
CV0010
CV0011
CV0014
CV0015
CV0019
CV0031
CV0043
CV0045
CV0046
CV0050
CV0051
CV0073
CV0074
CV0080
CV0086
CV0093
CV0100
CV0104
CV0122
CV0125
CV0128
CV0145
CV0149
CV0150
CV0156
CV0159
CV0160
CV0186
CV0193
0
50
100
150

### Group C, 2 of 2

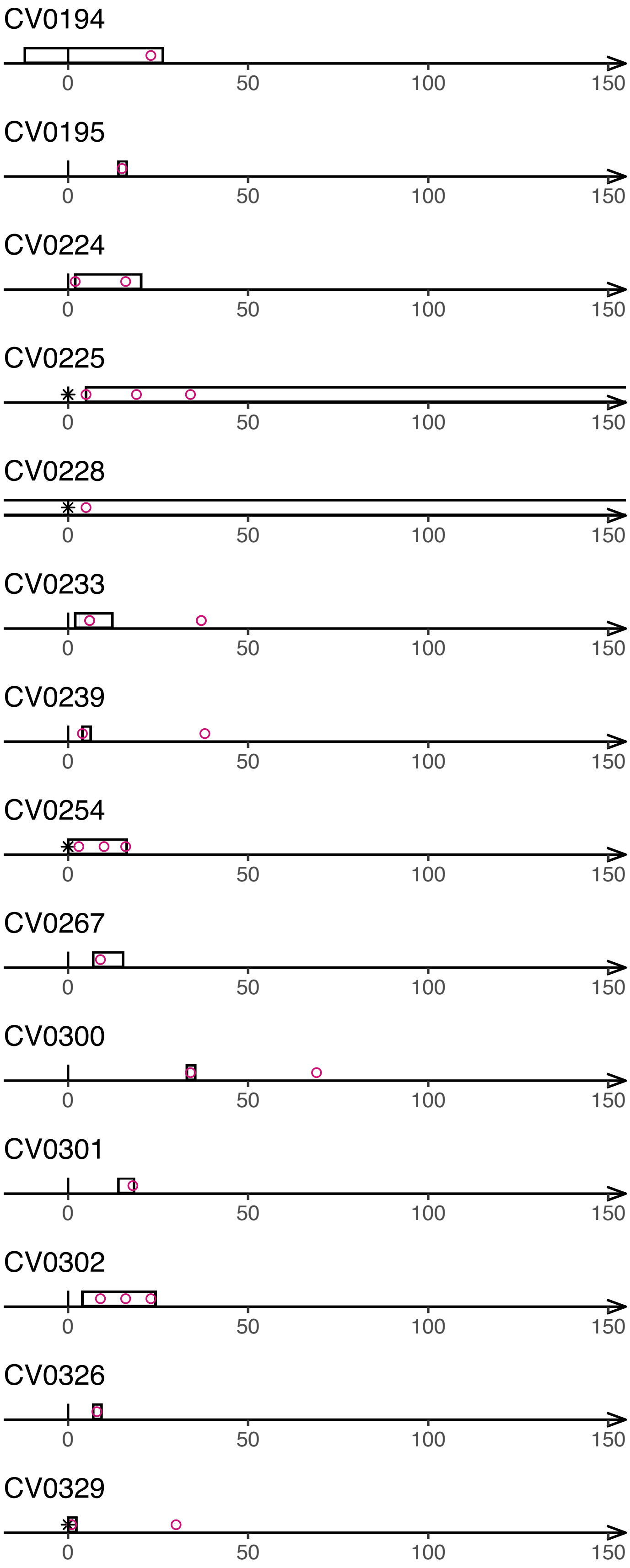
CV0194
0 50 100 150
CV0195
0 50 100 150
CV0224
0 50 100 150
CV0225
0 50 100 150
CV0228
0 50 100 150
CV0233
0 50 100 150
CV0239
0 50 100 150
CV0254
0 50 100 150
CV0267
0 50 100 150
CV0300
0 50 100 150
CV0301
0 50 100 150
CV0302
0 50 100 150
CV0326
0 50 100 150
CV0329
0 50 100 150

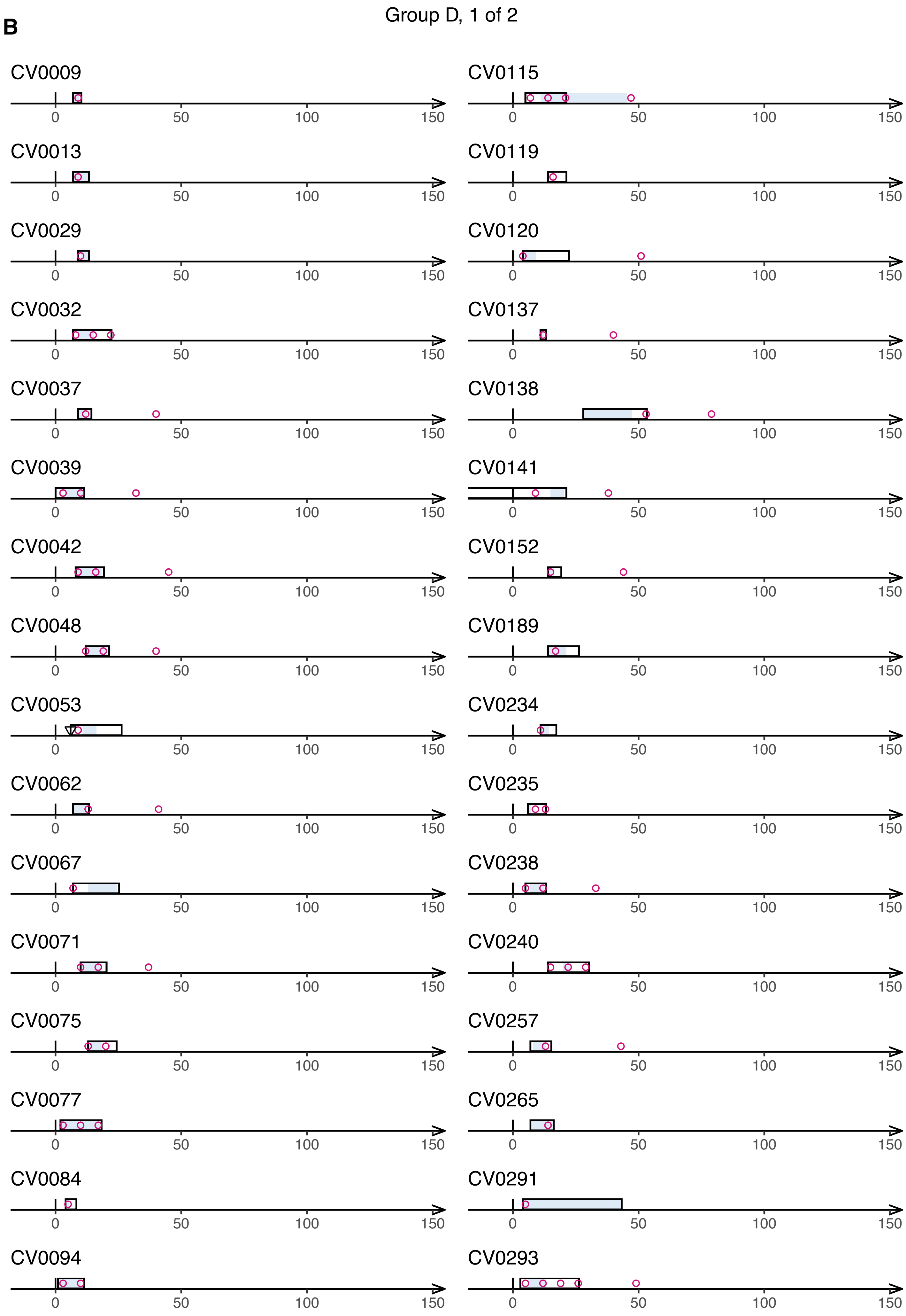

B
Group D, 1 of 2
CV0009
CV0013
CV0029
CV0032
CV0037
CV0039
CV0042
CV0048
CV0053
CV0062
CV0067
CV0071
CV0075
CV0077
CV0084
CV0094
CV0115
CV0119
CV0120
CV0137
CV0138
CV0141
CV0152
CV0189
CV0234
CV0235
CV0238
CV0240
CV0257
CV0265
CV0291
CV0293
0
50
100
150

### Group D, 2 of 2

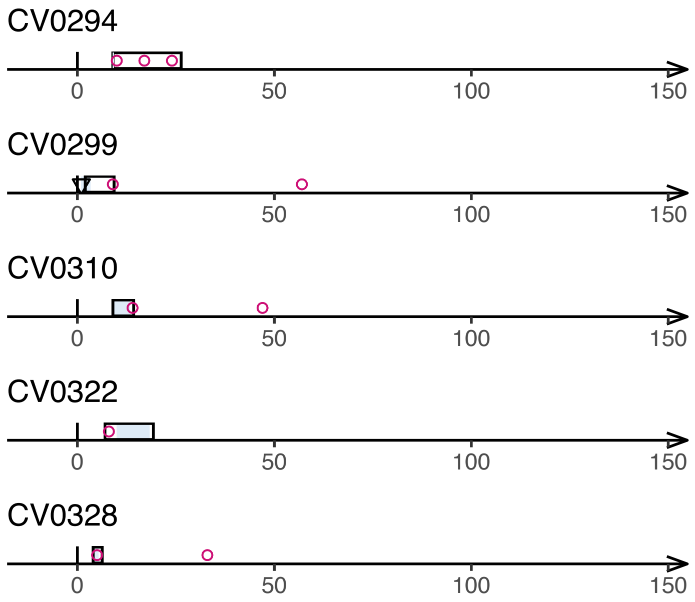
CV0294
0
50
100
150
CV0299
0
50
100
150
CV0310
0
50
100
150
CV0322
0
50
100
150
CV0328
0
50
100
150

**C** Group E, 1 of 2

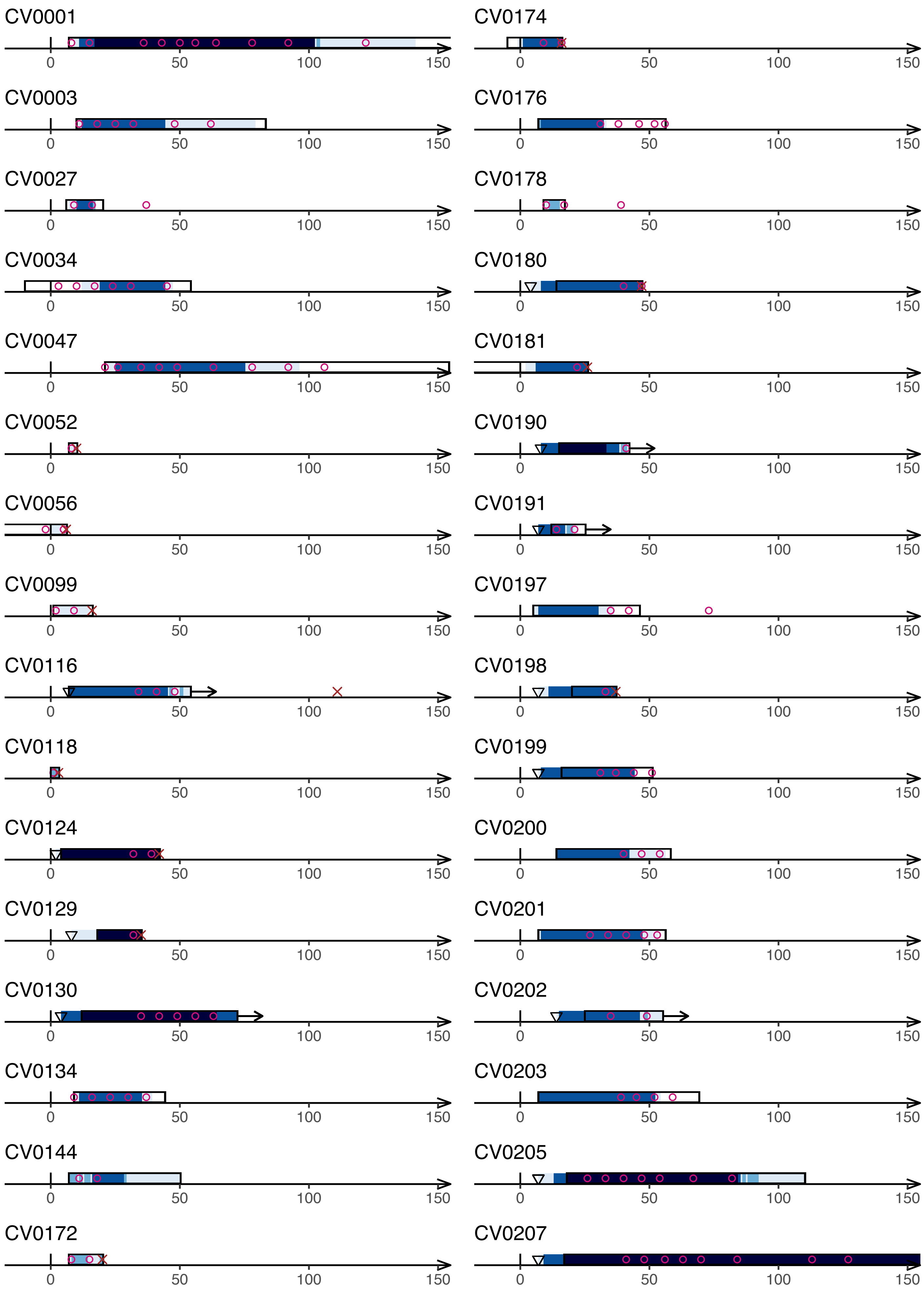

### Group E, 2 of 2

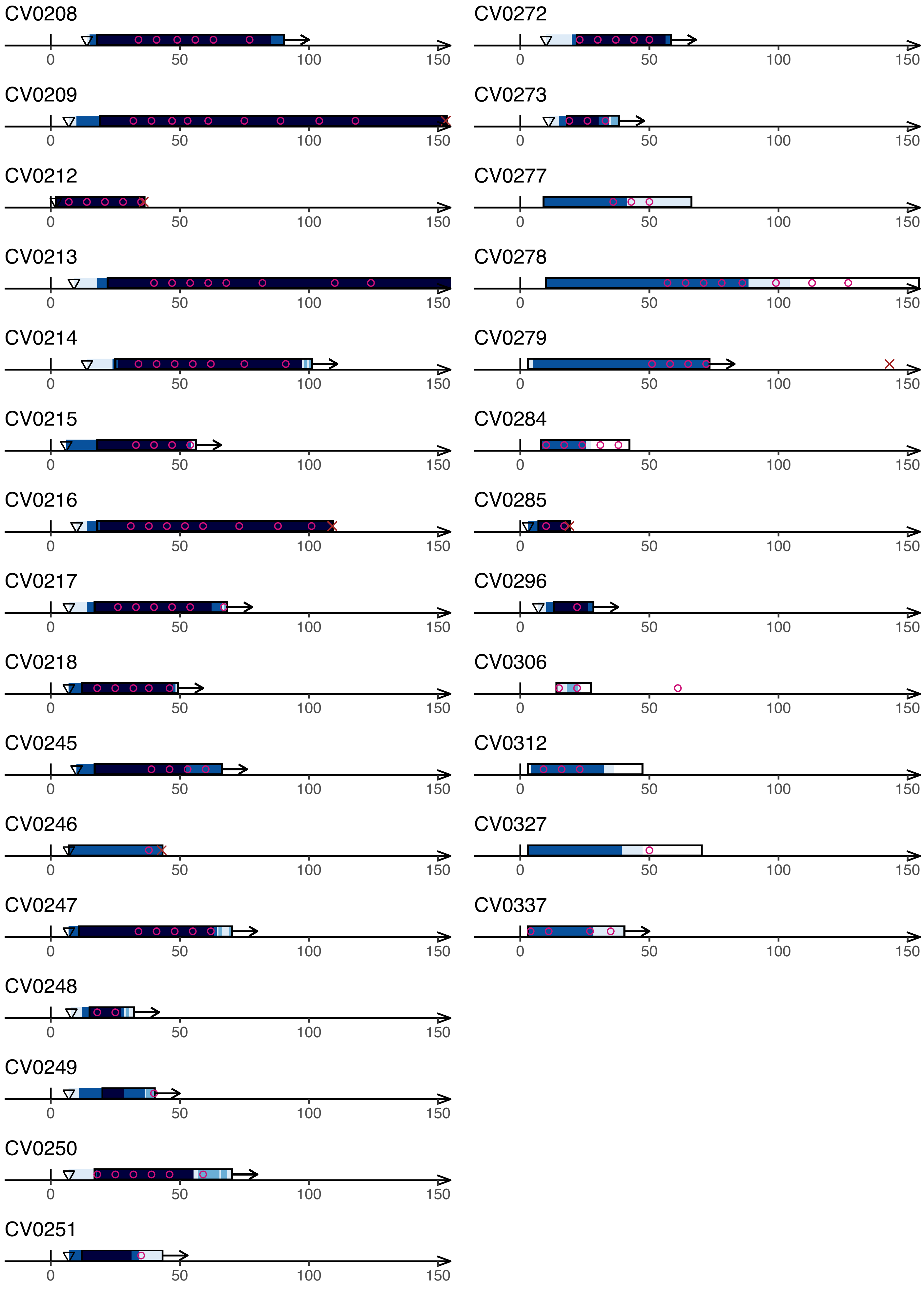
CV0208
CV0209
CV0212
CV0213
CV0214
CV0215
CV0216
CV0217
CV0218
CV0245
CV0246
CV0247
CV0248
CV0249
CV0250
CV0251
CV0272
CV0273
CV0277
CV0278
CV0279
CV0284
CV0285
CV0296
CV0306
CV0312
CV0327
CV0337
0
50
100
150
