## Supplemental Item 2 for "Delayed bystander CD8 T cell activation, early immune pathology and persistent dysregulation characterise severe COVID-19": Item_S2__IMMUNITY_15032021.pdf

### **Item S2:** Cell data – absolute counts

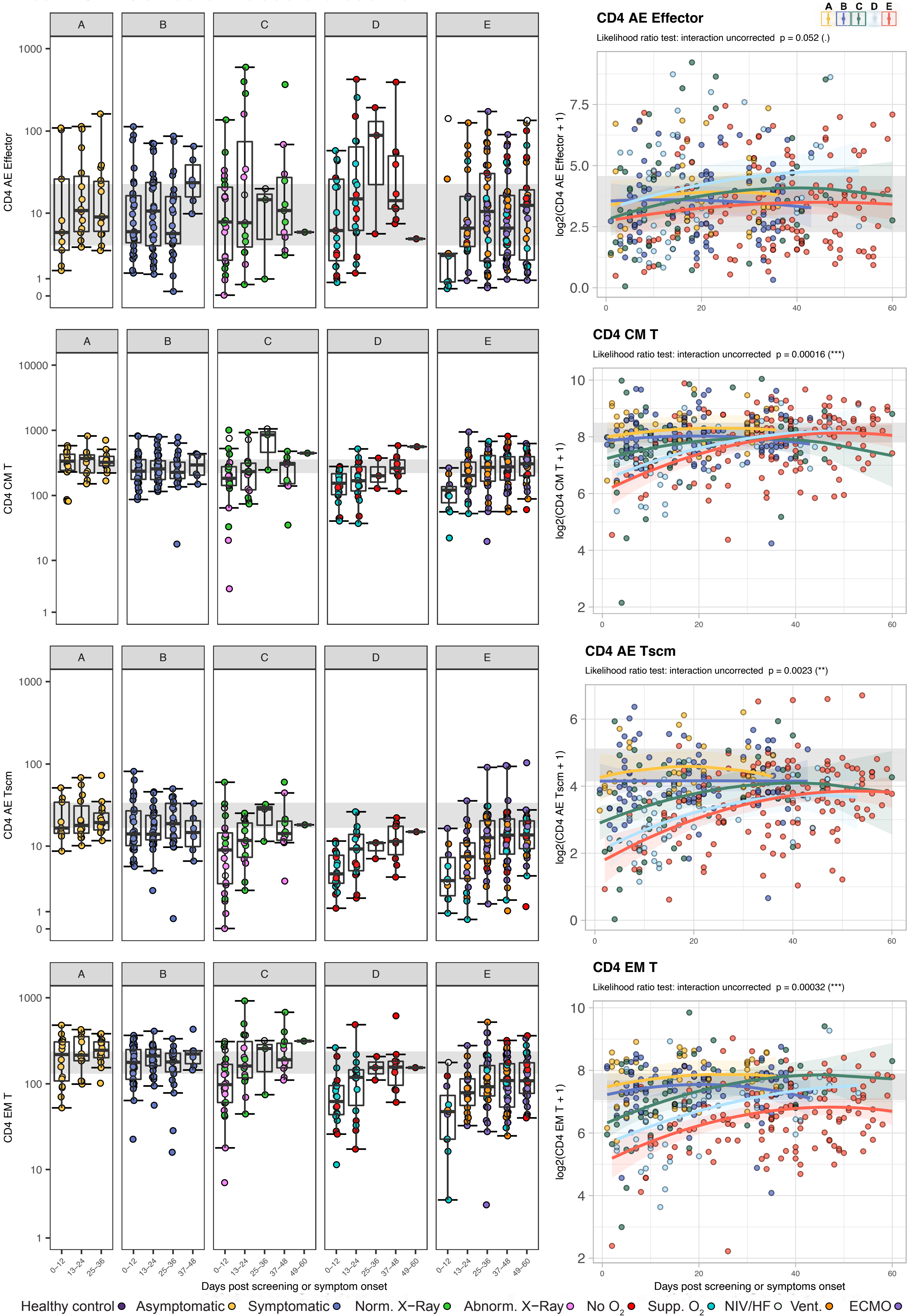

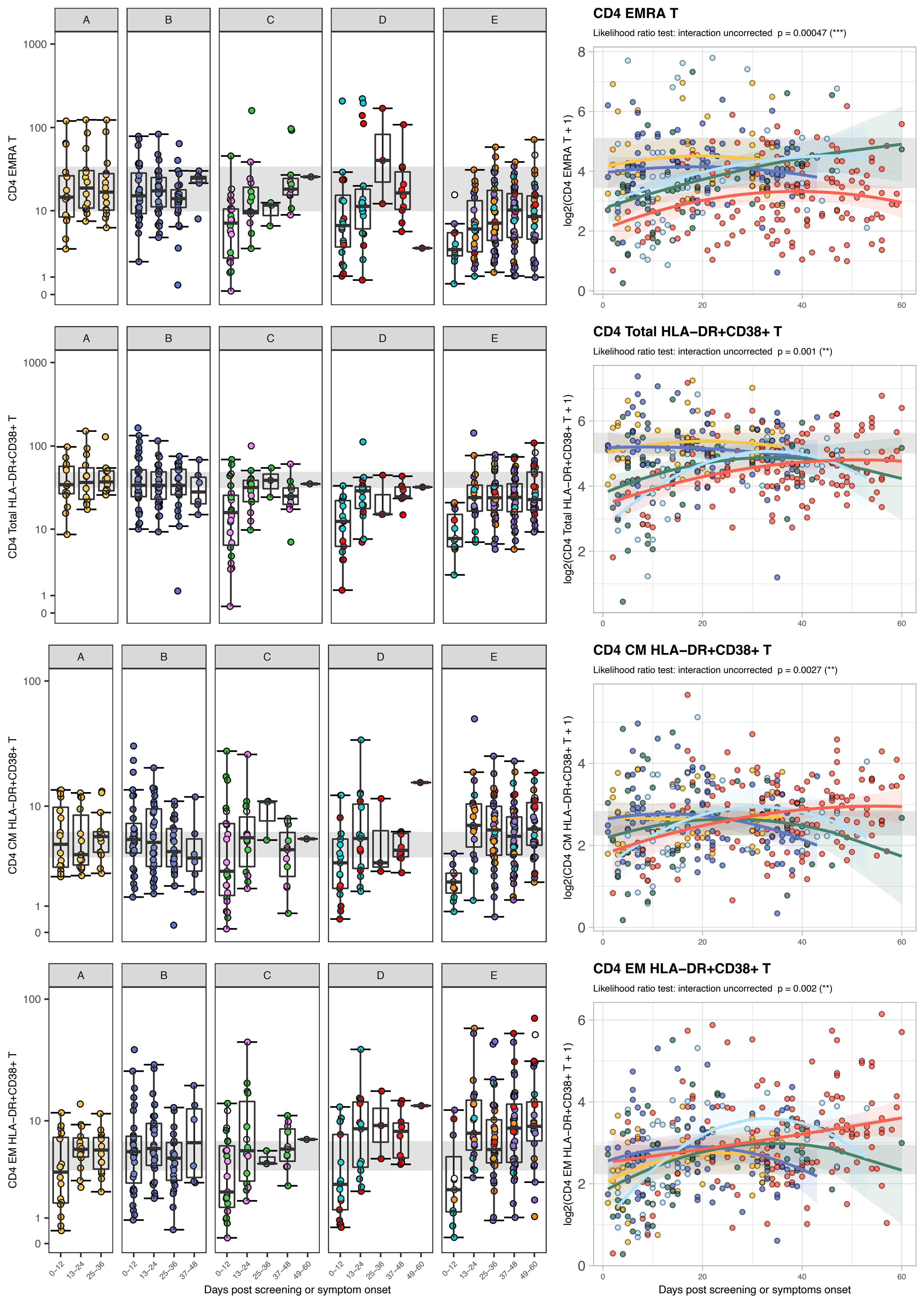

CD4 EMRA T
Likelihood ratio test: interaction uncorrected p = 0.00047 (***)
log2(CD4 EMRA T + 1)
CD4 Total HLA−DR+CD38+ T
Likelihood ratio test: interaction uncorrected p = 0.001 (**)
log2(CD4 Total HLA−DR+CD38+ T + 1)
CD4 CM HLA−DR+CD38+ T
Likelihood ratio test: interaction uncorrected p = 0.0027 (**)
log2(CD4 CM HLA−DR+CD38+ T + 1)
CD4 EM HLA−DR+CD38+ T
Likelihood ratio test: interaction uncorrected p = 0.002 (**)
log2(CD4 EM HLA−DR+CD38+ T + 1)
A
B
C
D
E
0−12
13−24
25−36
37−48
49−60
Days post screening or symptom onset
Days post screening or symptoms onset

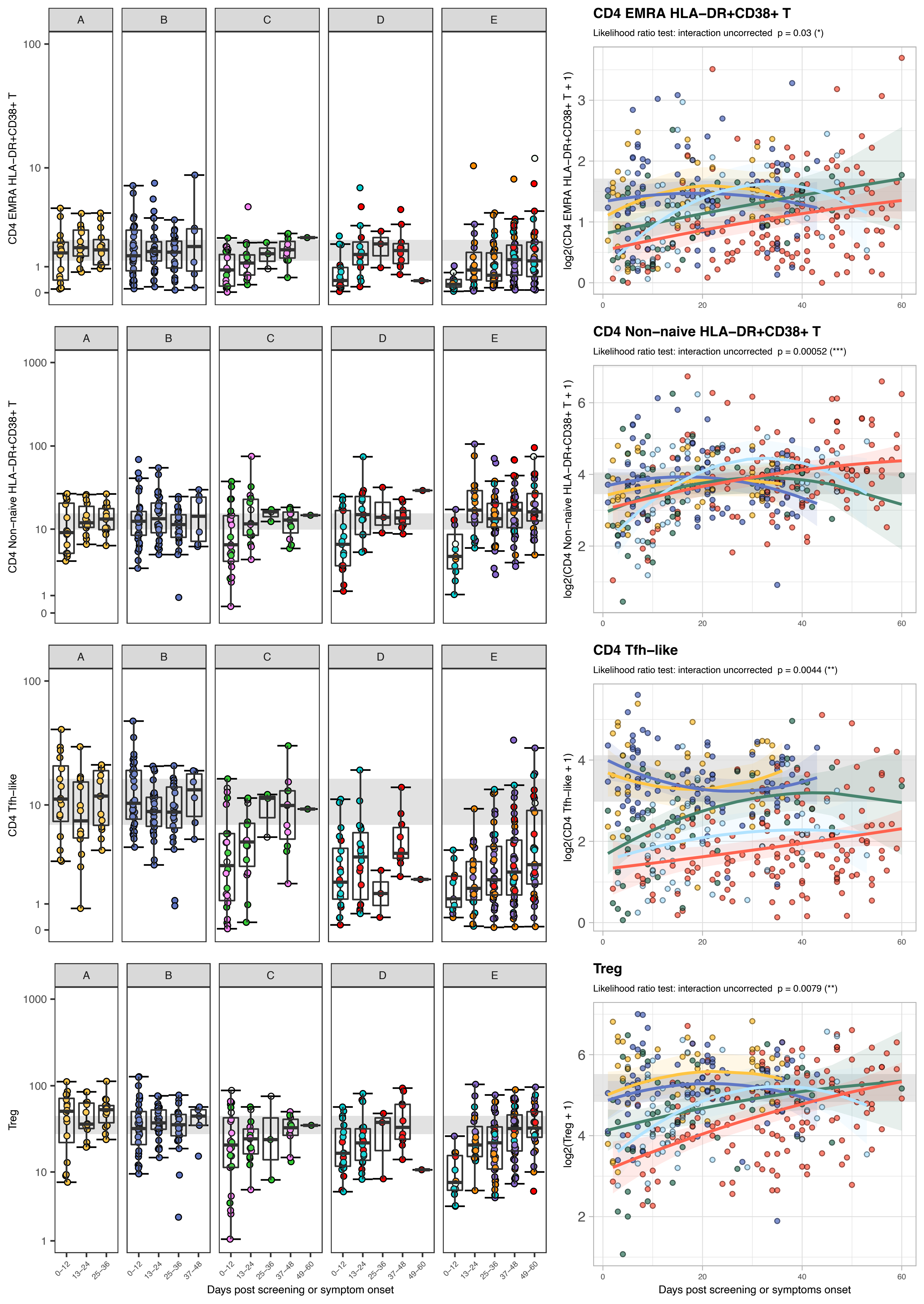

CD4 EMRA HLA−DR+CD38+ T
Likelihood ratio test: interaction uncorrected p = 0.03 (*)
log2(CD4 EMRA HLA−DR+CD38+ T + 1)
CD4 Non−naive HLA−DR+CD38+ T
Likelihood ratio test: interaction uncorrected p = 0.00052 (***)
log2(CD4 Non−naive HLA−DR+CD38+ T + 1)
CD4 Tfh−like
Likelihood ratio test: interaction uncorrected p = 0.0044 (**)
log2(CD4 Tfh−like + 1)
Treg
Likelihood ratio test: interaction uncorrected p = 0.0079 (**)
log2(Treg + 1)
A
B
C
D
E
Treg
0−12
13−24
25−36
37−48
49−60
Days post screening or symptom onset
Days post screening or symptoms onset

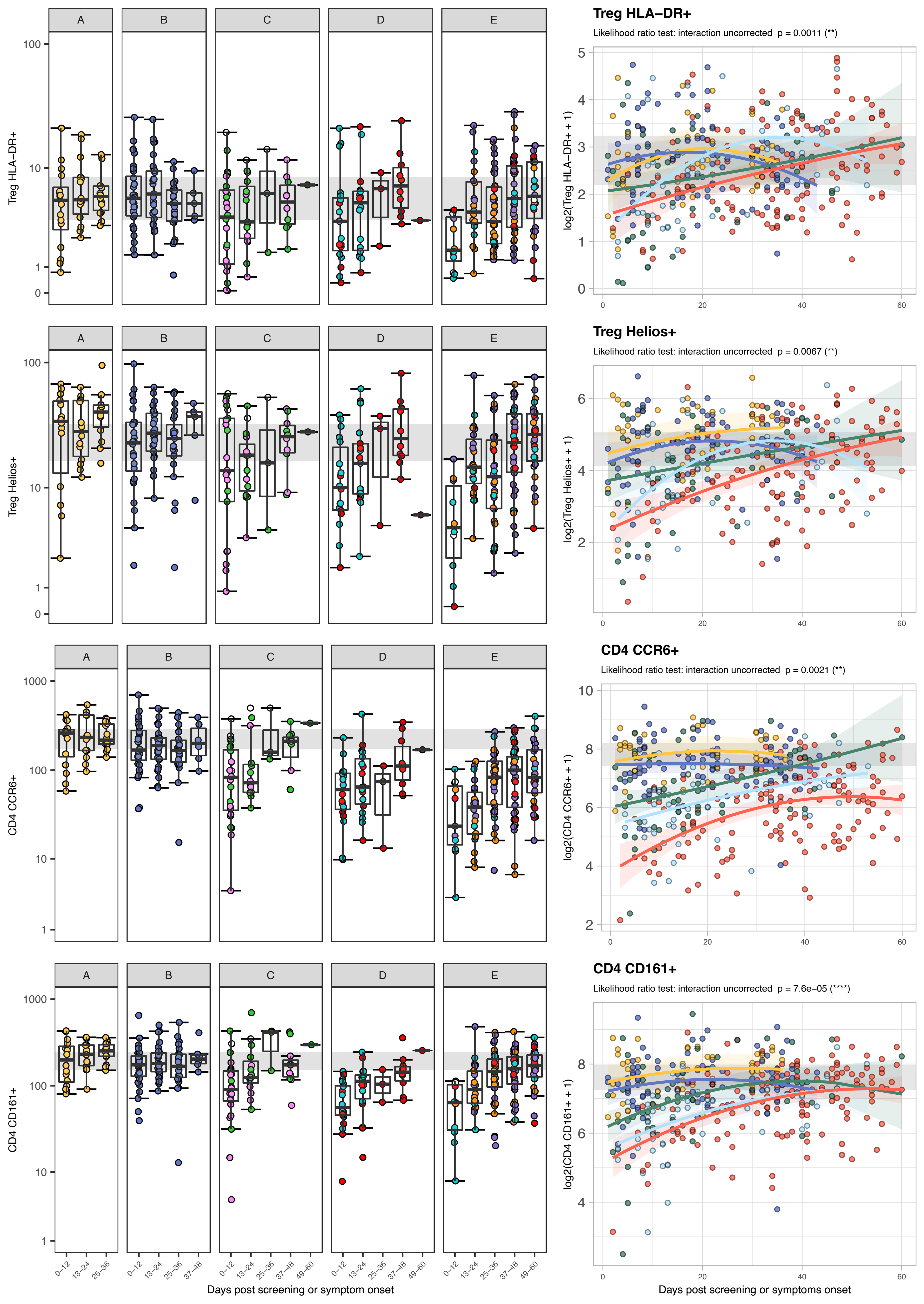

Treg HLA−DR+
Likelihood ratio test: interaction uncorrected p = 0.0011 (**)
Treg Helios+
Likelihood ratio test: interaction uncorrected p = 0.0067 (**)
CD4 CCR6+
Likelihood ratio test: interaction uncorrected p = 0.0021 (**)
CD4 CD161+
Likelihood ratio test: interaction uncorrected p = 7.6e−05 (****)
A
B
C
D
E
Treg HLA−DR+
Treg Helios+
CD4 CCR6+
CD4 CD161+
log2(Treg HLA−DR+ + 1)
log2(Treg Helios+ + 1)
log2(CD4 CCR6+ + 1)
log2(CD4 CD161+ + 1)
0−12
13−24
25−36
37−48
49−60
Days post screening or symptom onset
Days post screening or symptoms onset

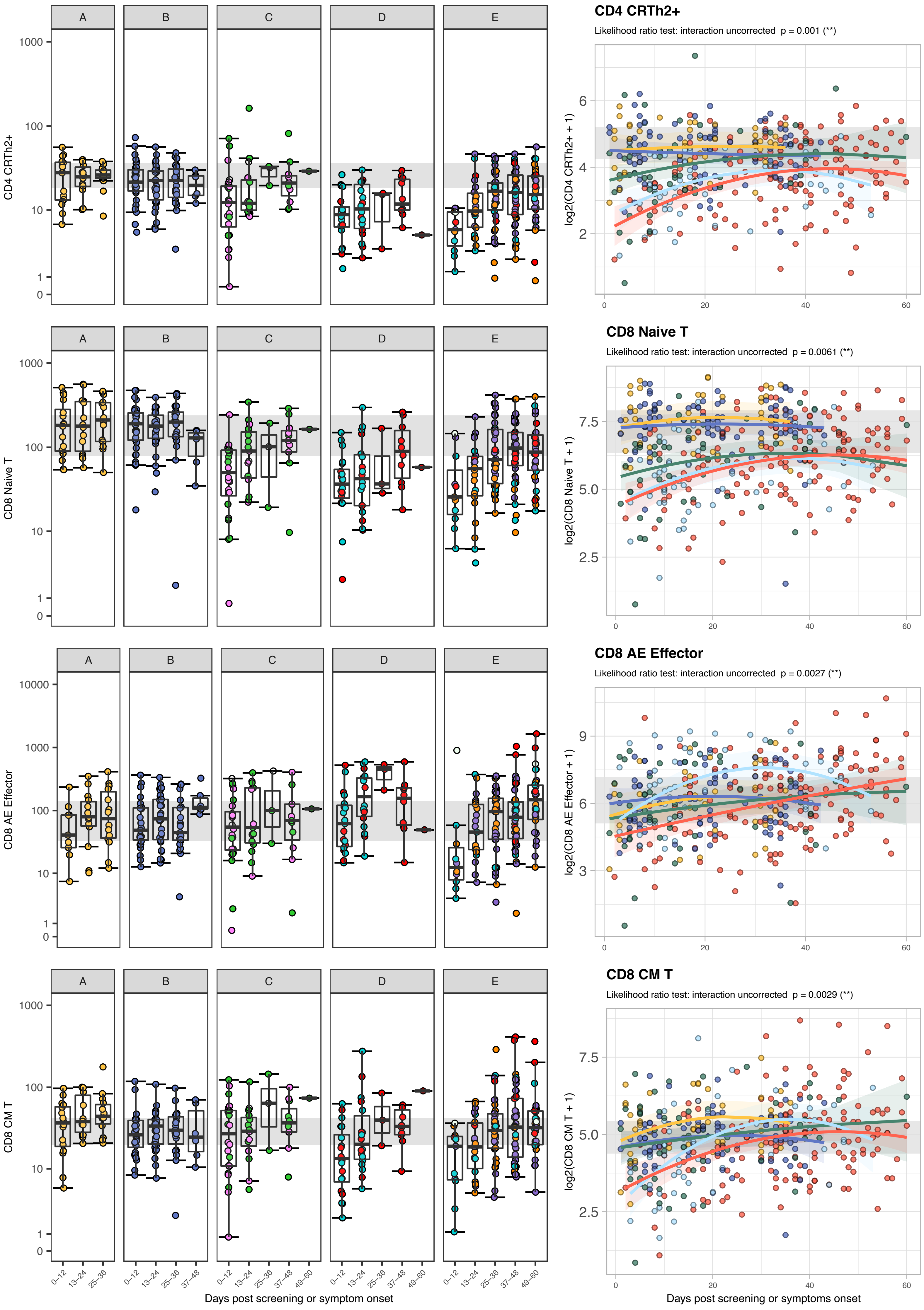

CD4 CRTh2+
Likelihood ratio test: interaction uncorrected p = 0.001 (**)
CD8 Naive T
Likelihood ratio test: interaction uncorrected p = 0.0061 (**)
CD8 AE Effector
Likelihood ratio test: interaction uncorrected p = 0.0027 (**)
CD8 CM T
Likelihood ratio test: interaction uncorrected p = 0.0029 (**)
A
B
C
D
E
CD4 CRTh2+
CD8 Naive T
CD8 AE Effector
CD8 CM T
log2(CD4 CRTh2+ + 1)
log2(CD8 Naive T + 1)
log2(CD8 AE Effector + 1)
log2(CD8 CM T + 1)
0–12
13–24
25–36
37–48
49–60
Days post screening or symptom onset
Days post screening or symptoms onset

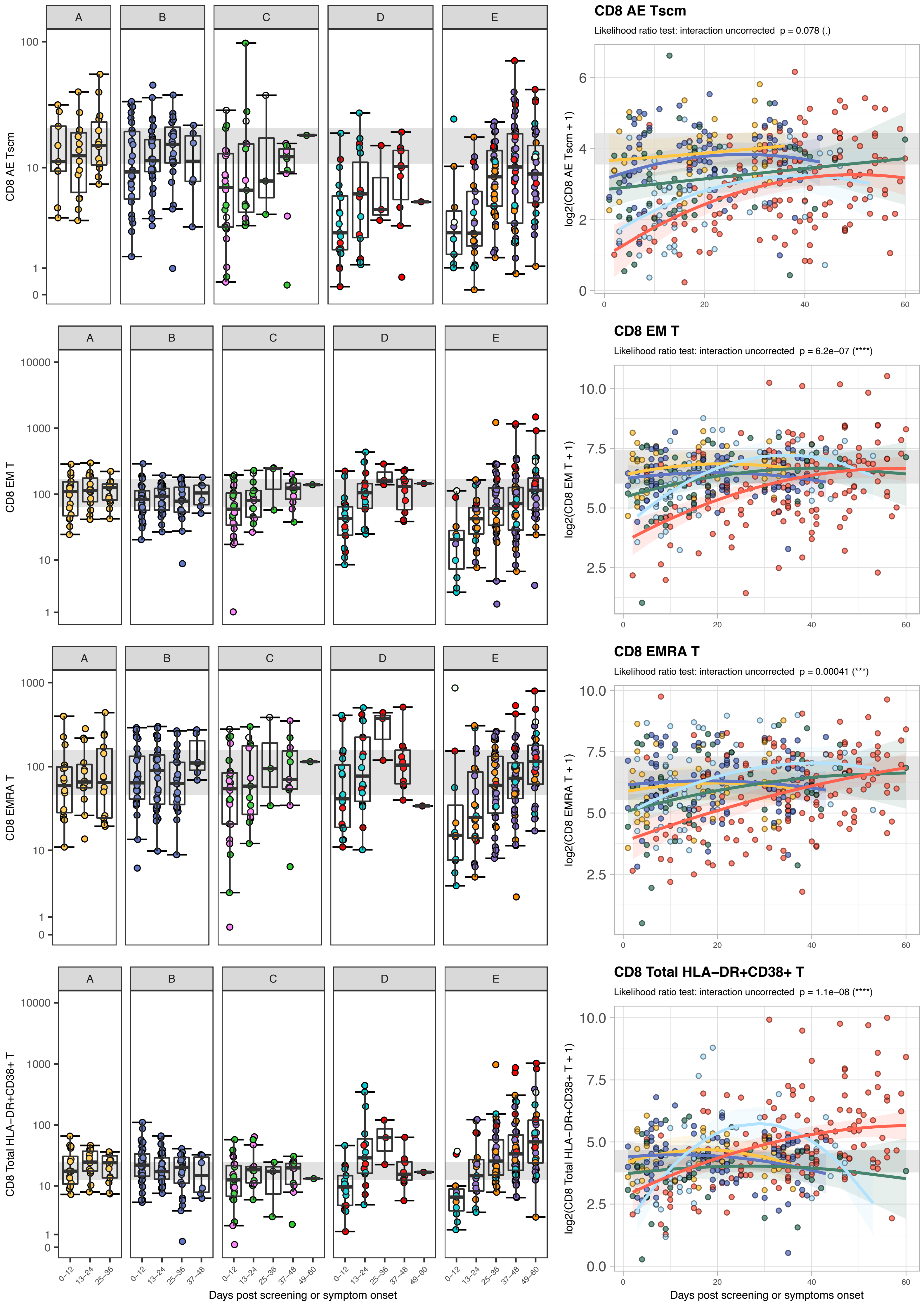

A
B
C
D
E
CD8 AE Tscm
CD8 EM T
CD8 EMRA T
CD8 Total HLA−DR+CD38+ T
CD8 AE Tscm
Likelihood ratio test: interaction uncorrected p = 0.078 (.)
log2(CD8 AE Tscm + 1)
CD8 EM T
Likelihood ratio test: interaction uncorrected p = 6.2e−07 (****)
log2(CD8 EM T + 1)
CD8 EMRA T
Likelihood ratio test: interaction uncorrected p = 0.00041 (***)
log2(CD8 EMRA T + 1)
CD8 Total HLA−DR+CD38+ T
Likelihood ratio test: interaction uncorrected p = 1.1e−08 (****)
log2(CD8 Total HLA−DR+CD38+ T + 1)
0−12
13−24
25−36
37−48
49−60
Days post screening or symptom onset
Days post screening or symptoms onset

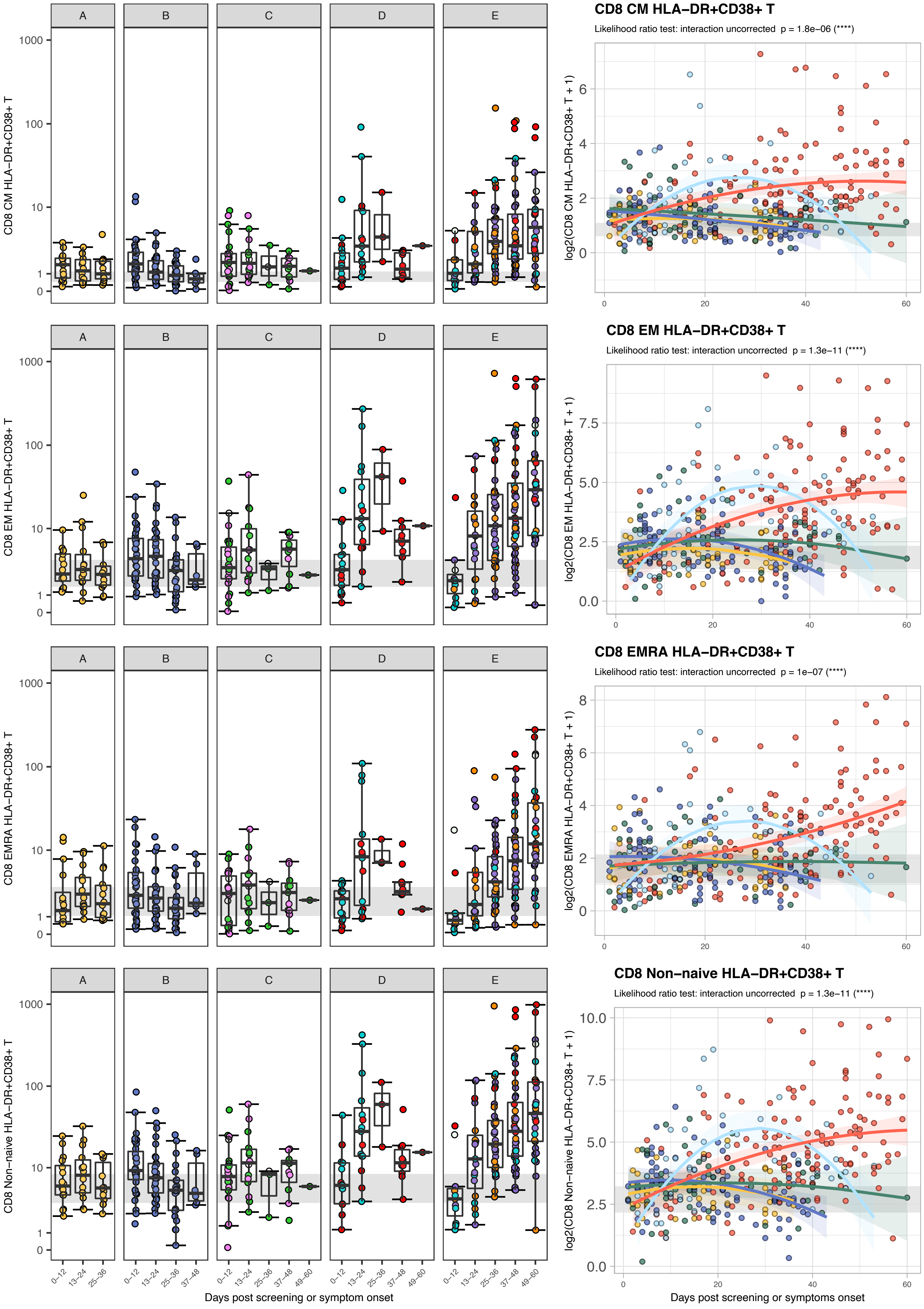

CD8 CM HLA−DR+CD38+ T
Likelihood ratio test: interaction uncorrected p = 1.8e−06 (****)
log2(CD8 CM HLA−DR+CD38+ T + 1)
CD8 EM HLA−DR+CD38+ T
Likelihood ratio test: interaction uncorrected p = 1.3e−11 (****)
log2(CD8 EM HLA−DR+CD38+ T + 1)
CD8 EMRA HLA−DR+CD38+ T
Likelihood ratio test: interaction uncorrected p = 1e−07 (****)
log2(CD8 EMRA HLA−DR+CD38+ T + 1)
CD8 Non−naive HLA−DR+CD38+ T
Likelihood ratio test: interaction uncorrected p = 1.3e−11 (****)
log2(CD8 Non−naive HLA−DR+CD38+ T + 1)
A
B
C
D
E
0−12
13−24
25−36
37−48
49−60
Days post screening or symptom onset
Days post screening or symptoms onset

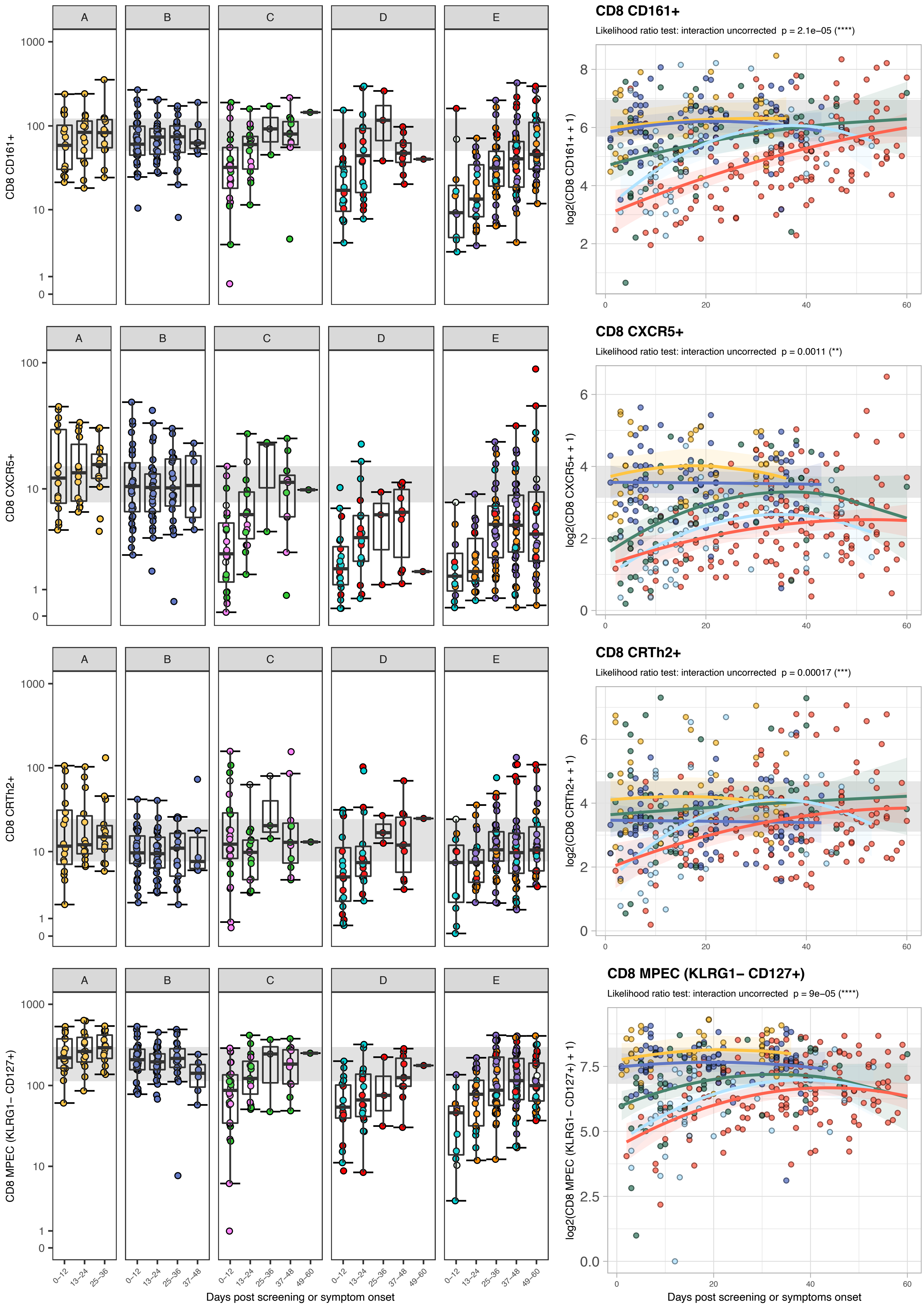

A
B
C
D
E
CD8 CD161+
Likelihood ratio test: interaction uncorrected p = 2.1e−05 (****)
log2(CD8 CD161+ + 1)
CD8 CXCR5+
Likelihood ratio test: interaction uncorrected p = 0.0011 (**)
log2(CD8 CXCR5+ + 1)
CD8 CRTh2+
Likelihood ratio test: interaction uncorrected p = 0.00017 (***)
log2(CD8 CRTh2+ + 1)
CD8 MPEC (KLRG1− CD127+)
Likelihood ratio test: interaction uncorrected p = 9e−05 (****)
log2(CD8 MPEC (KLRG1− CD127+) + 1)
0–12
13–24
25–36
37–48
49–60
Days post screening or symptom onset
Days post screening or symptoms onset

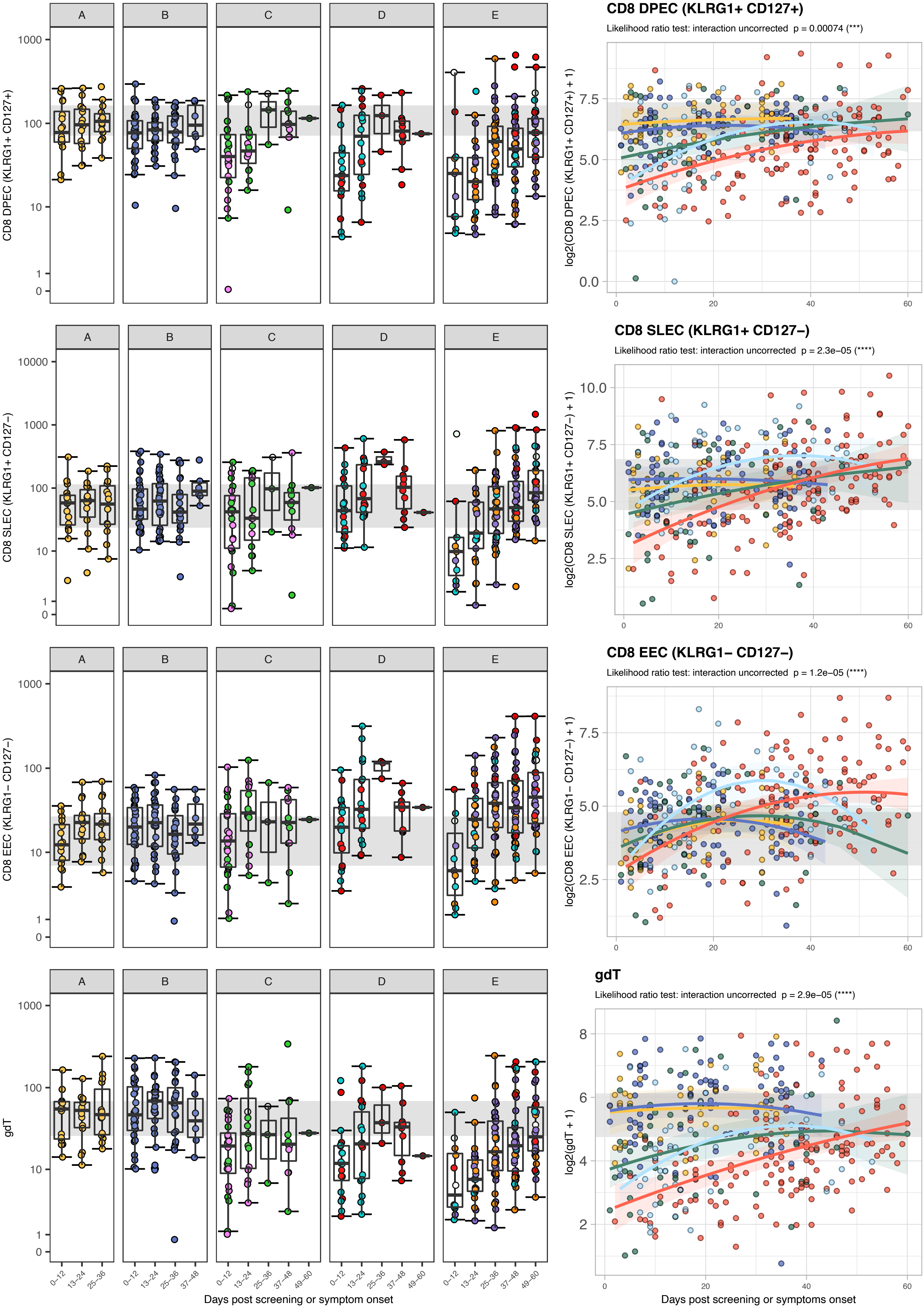

A
B
C
D
E
CD8 DPEC (KLRG1+ CD127+)
log2(CD8 DPEC (KLRG1+ CD127+) + 1)
CD8 DPEC (KLRG1+ CD127+)
Likelihood ratio test: interaction uncorrected p = 0.00074 (***)
CD8 SLEC (KLRG1+ CD127−)
log2(CD8 SLEC (KLRG1+ CD127−) + 1)
CD8 SLEC (KLRG1+ CD127−)
Likelihood ratio test: interaction uncorrected p = 2.3e−05 (****)
CD8 EEC (KLRG1− CD127−)
log2(CD8 EEC (KLRG1− CD127−) + 1)
CD8 EEC (KLRG1− CD127−)
Likelihood ratio test: interaction uncorrected p = 1.2e−05 (****)
gdT
log2(gdT + 1)
gdT
Likelihood ratio test: interaction uncorrected p = 2.9e−05 (****)
0–12
13–24
25–36
37–48
49–60
Days post screening or symptom onset
Days post screening or symptoms onset

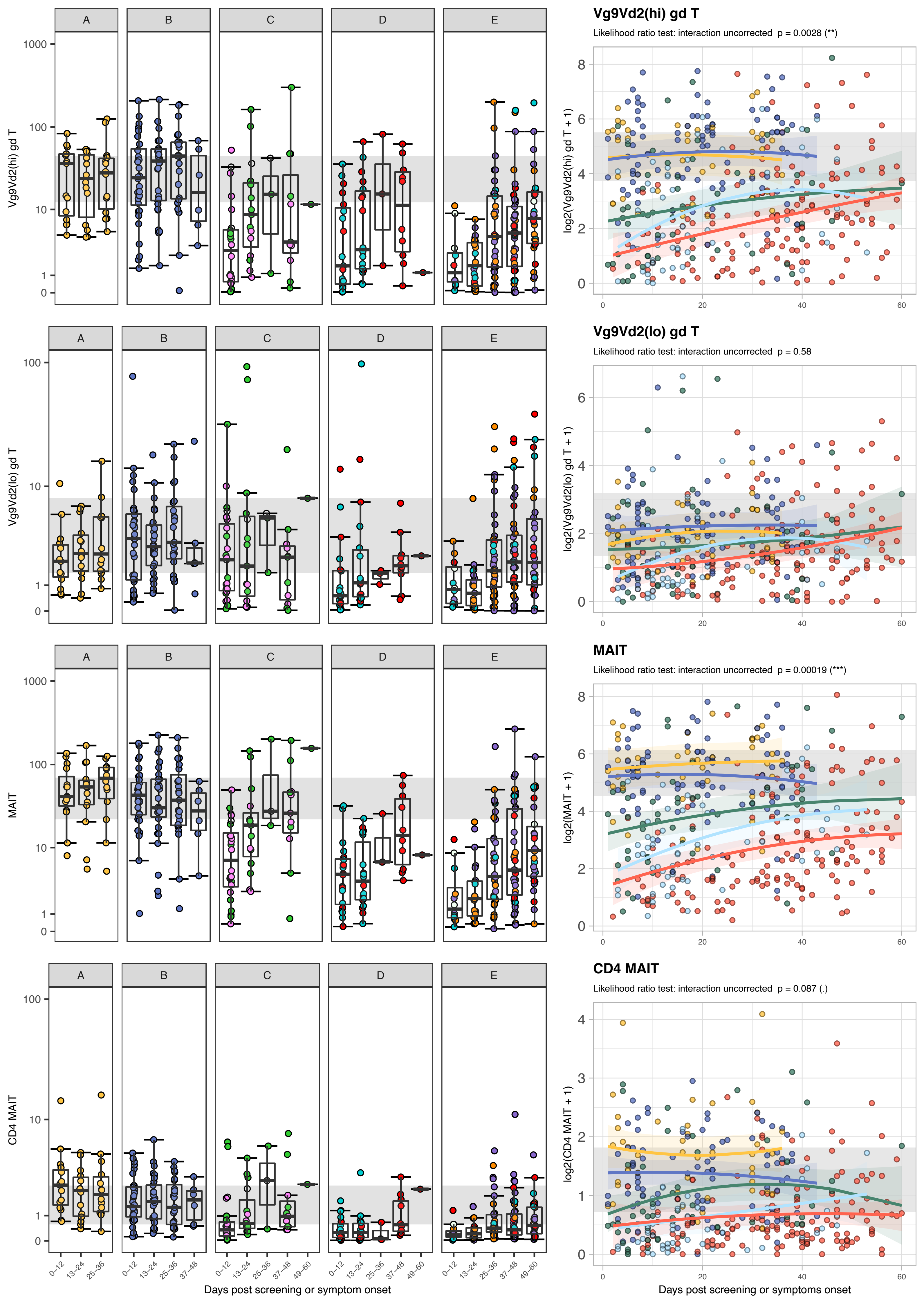

Vg9Vd2(hi) gd T
Likelihood ratio test: interaction uncorrected p = 0.0028 (**)
Vg9Vd2(lo) gd T
Likelihood ratio test: interaction uncorrected p = 0.58
MAIT
Likelihood ratio test: interaction uncorrected p = 0.00019 (***)
CD4 MAIT
Likelihood ratio test: interaction uncorrected p = 0.087 (.)
A
B
C
D
E
Vg9Vd2(hi) gd T
Vg9Vd2(lo) gd T
MAIT
CD4 MAIT
log2(Vg9Vd2(hi) gd T + 1)
log2(Vg9Vd2(lo) gd T + 1)
log2(MAIT + 1)
log2(CD4 MAIT + 1)
0–12
13–24
25–36
37–48
49–60
Days post screening or symptom onset
Days post screening or symptoms onset

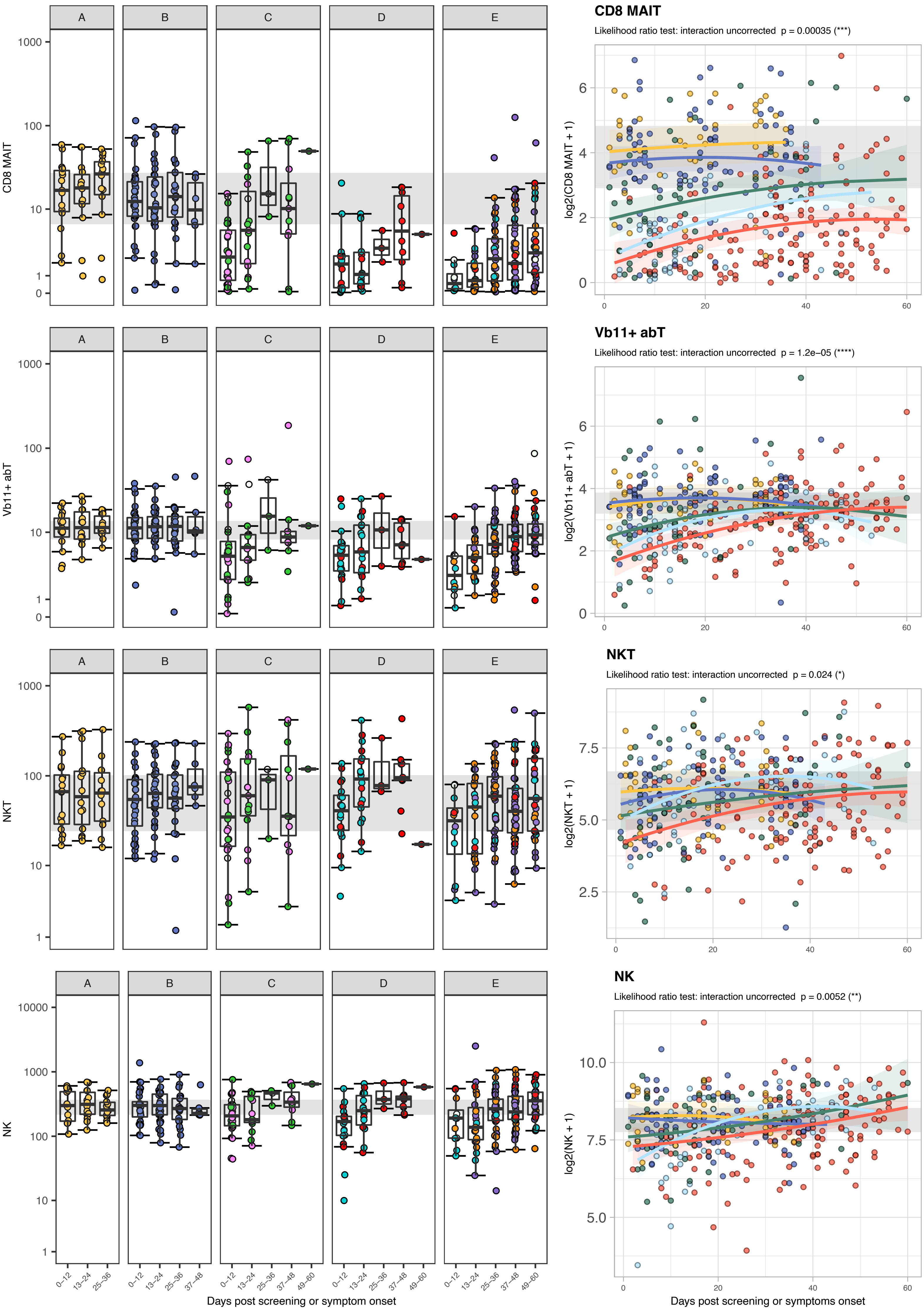

CD8 MAIT
Likelihood ratio test: interaction uncorrected p = 0.00035 (***)
log2(CD8 MAIT + 1)
Vb11+ abT
Likelihood ratio test: interaction uncorrected p = 1.2e−05 (****)
log2(Vb11+ abT + 1)
NKT
Likelihood ratio test: interaction uncorrected p = 0.024 (*)
log2(NKT + 1)
NK
Likelihood ratio test: interaction uncorrected p = 0.0052 (**)
log2(NK + 1)
A
B
C
D
E
CD8 MAIT
Vb11+ abT
NKT
NK
0–12
13–24
25–36
37–48
49–60
Days post screening or symptom onset
Days post screening or symptoms onset

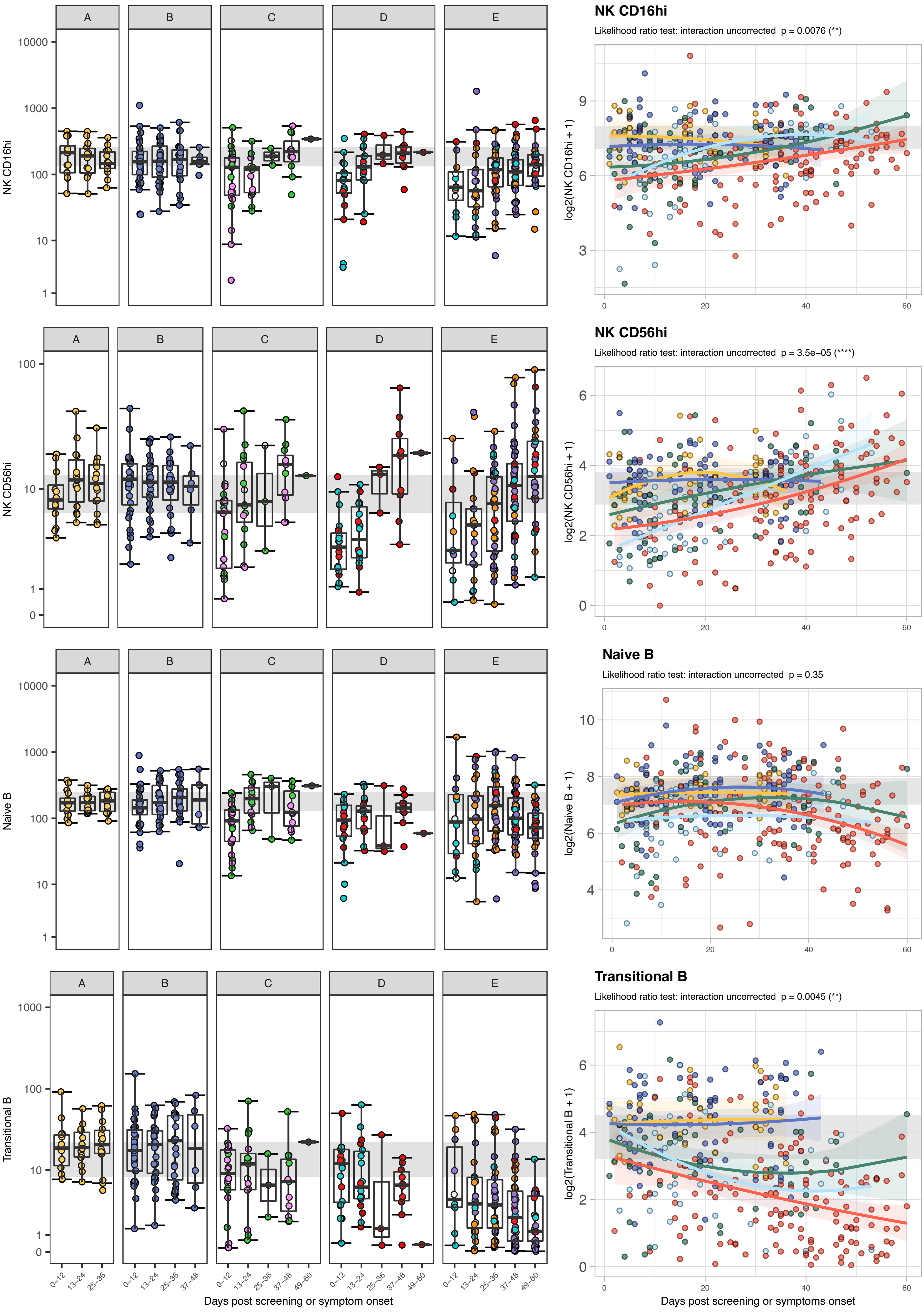

A
B
C
D
E
NK CD16hi
NK CD16hi
Likelihood ratio test: interaction uncorrected p = 0.0076 (**)
log2(NK CD16hi + 1)
NK CD56hi
NK CD56hi
Likelihood ratio test: interaction uncorrected p = 3.5e−05 (****)
log2(NK CD56hi + 1)
Naive B
Naive B
Likelihood ratio test: interaction uncorrected p = 0.35
log2(Naive B + 1)
Transitional B
Transitional B
Likelihood ratio test: interaction uncorrected p = 0.0045 (**)
log2(Transitional B + 1)
0–12
13–24
25–36
37–48
49–60
Days post screening or symptom onset
Days post screening or symptoms onset

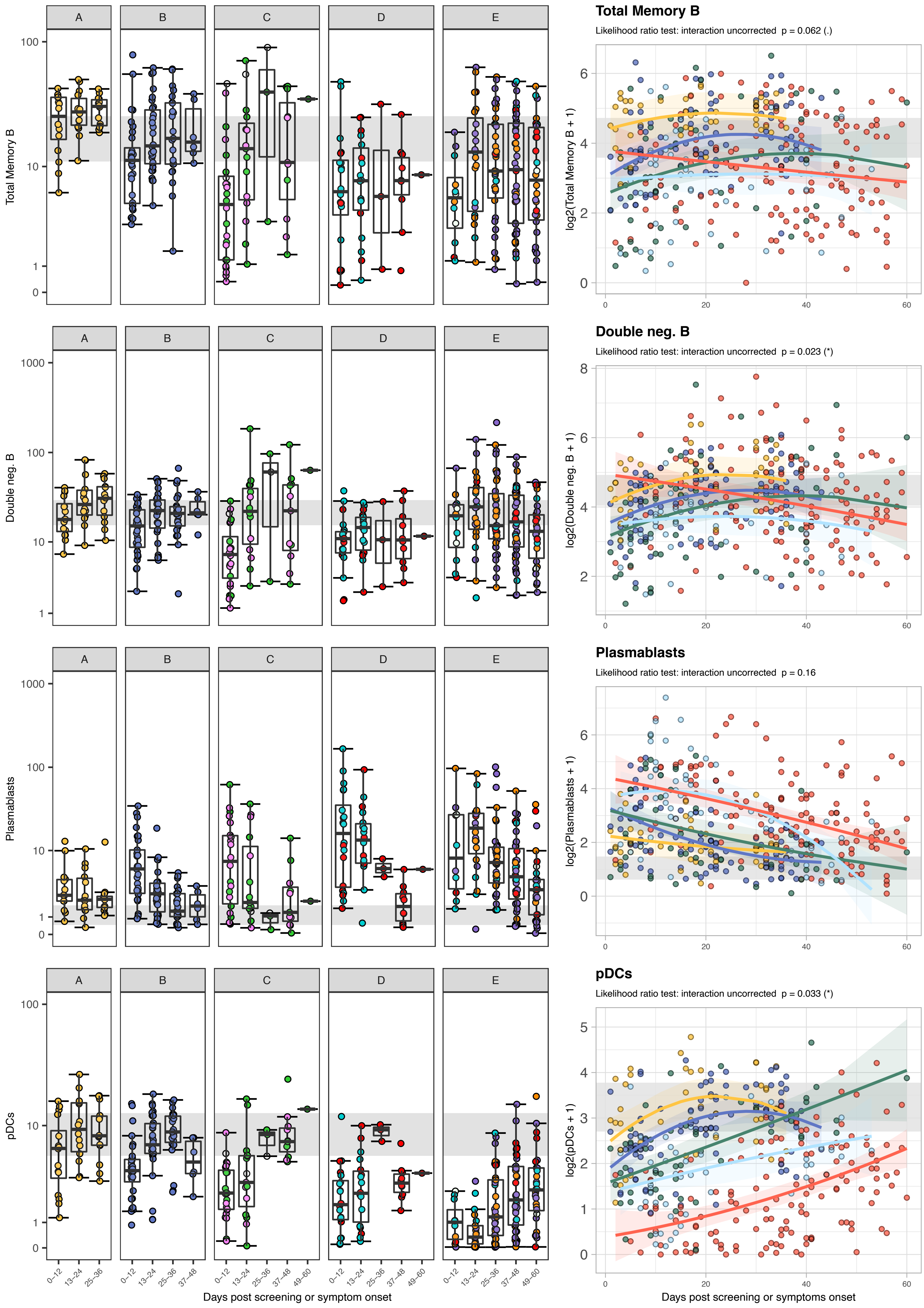
Total Memory B
Likelihood ratio test: interaction uncorrected p = 0.062 (.)
log2(Total Memory B + 1)
Double neg. B
Likelihood ratio test: interaction uncorrected p = 0.023 (*)
log2(Double neg. B + 1)
Plasmablasts
Likelihood ratio test: interaction uncorrected p = 0.16
log2(Plasmablasts + 1)
pDCs
Likelihood ratio test: interaction uncorrected p = 0.033 (*)
log2(pDCs + 1)
A
B
C
D
E
Total Memory B
Double neg. B
Plasmablasts
pDCs
0–12
13–24
25–36
37–48
49–60
Days post screening or symptom onset
Days post screening or symptoms onset

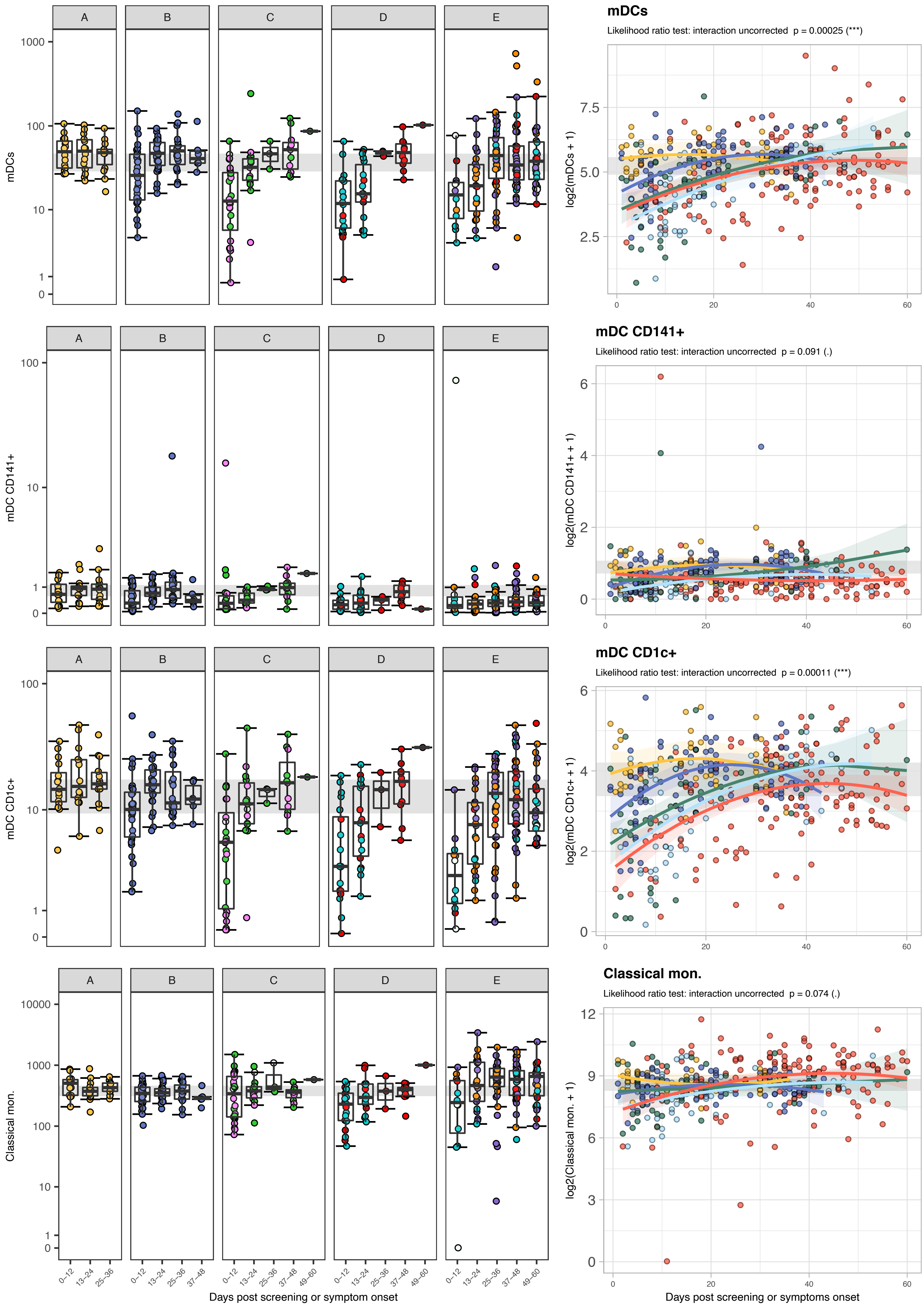
A
B
C
D
E
mDCs
mDC CD141+
mDC CD1c+
Classical mon.
mDCs
Likelihood ratio test: interaction uncorrected p = 0.00025 (***)
log2(mDCs + 1)
mDC CD141+
Likelihood ratio test: interaction uncorrected p = 0.091 (.)
log2(mDC CD141+ + 1)
mDC CD1c+
Likelihood ratio test: interaction uncorrected p = 0.00011 (***)
log2(mDC CD1c+ + 1)
Classical mon.
Likelihood ratio test: interaction uncorrected p = 0.074 (.)
log2(Classical mon. + 1)
0–12
13–24
25–36
37–48
49–60
Days post screening or symptom onset
Days post screening or symptoms onset

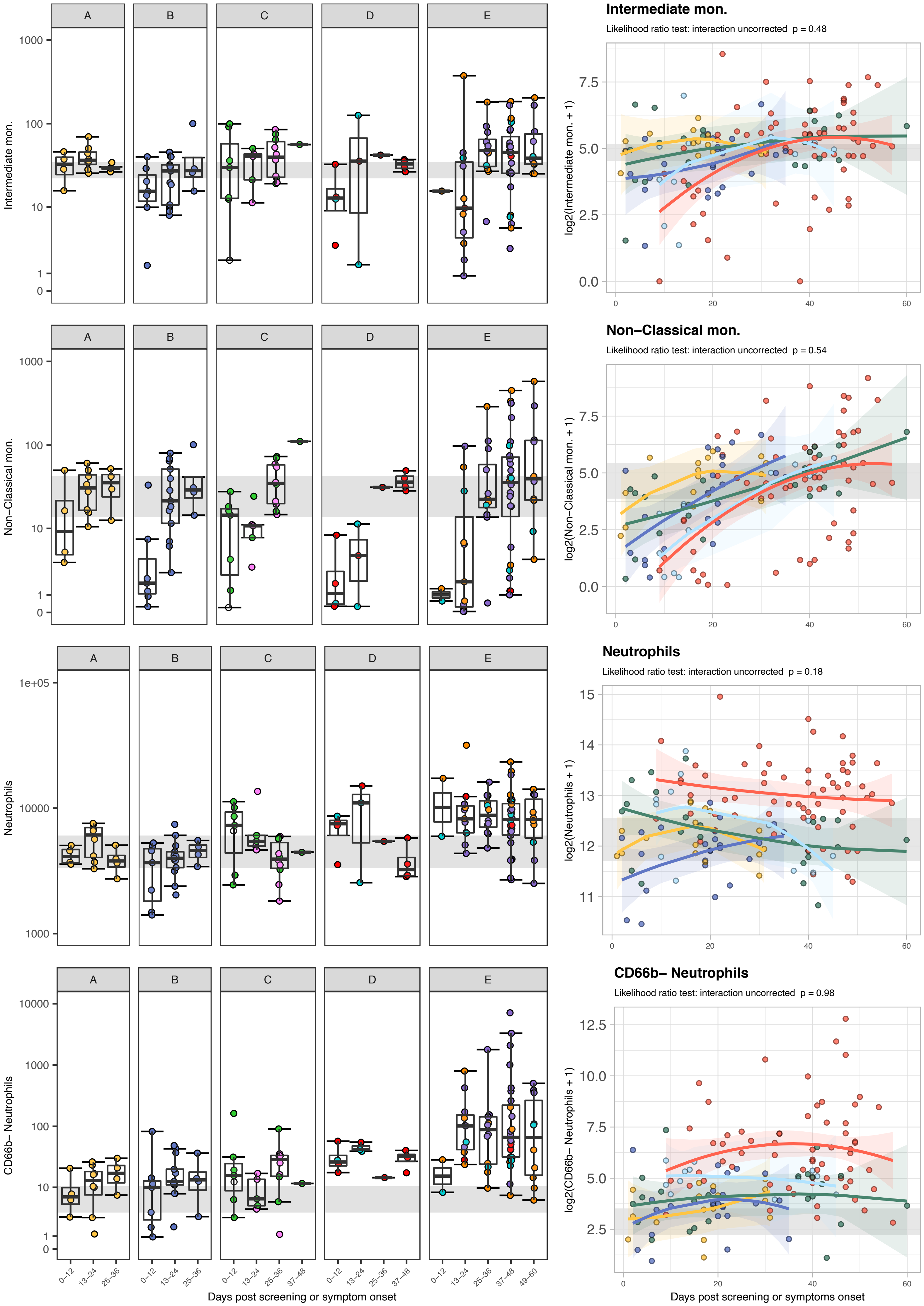
Intermediate mon.
Likelihood ratio test: interaction uncorrected p = 0.48
log2(Intermediate mon. + 1)
Intermediate mon.
Non−Classical mon.
Likelihood ratio test: interaction uncorrected p = 0.54
log2(Non−Classical mon. + 1)
Non−Classical mon.
Neutrophils
Likelihood ratio test: interaction uncorrected p = 0.18
log2(Neutrophils + 1)
Neutrophils
CD66b− Neutrophils
Likelihood ratio test: interaction uncorrected p = 0.98
log2(CD66b− Neutrophils + 1)
CD66b− Neutrophils
A
B
C
D
E
0−12
13−24
25−36
37−48
49−60
Days post screening or symptom onset
Days post screening or symptoms onset

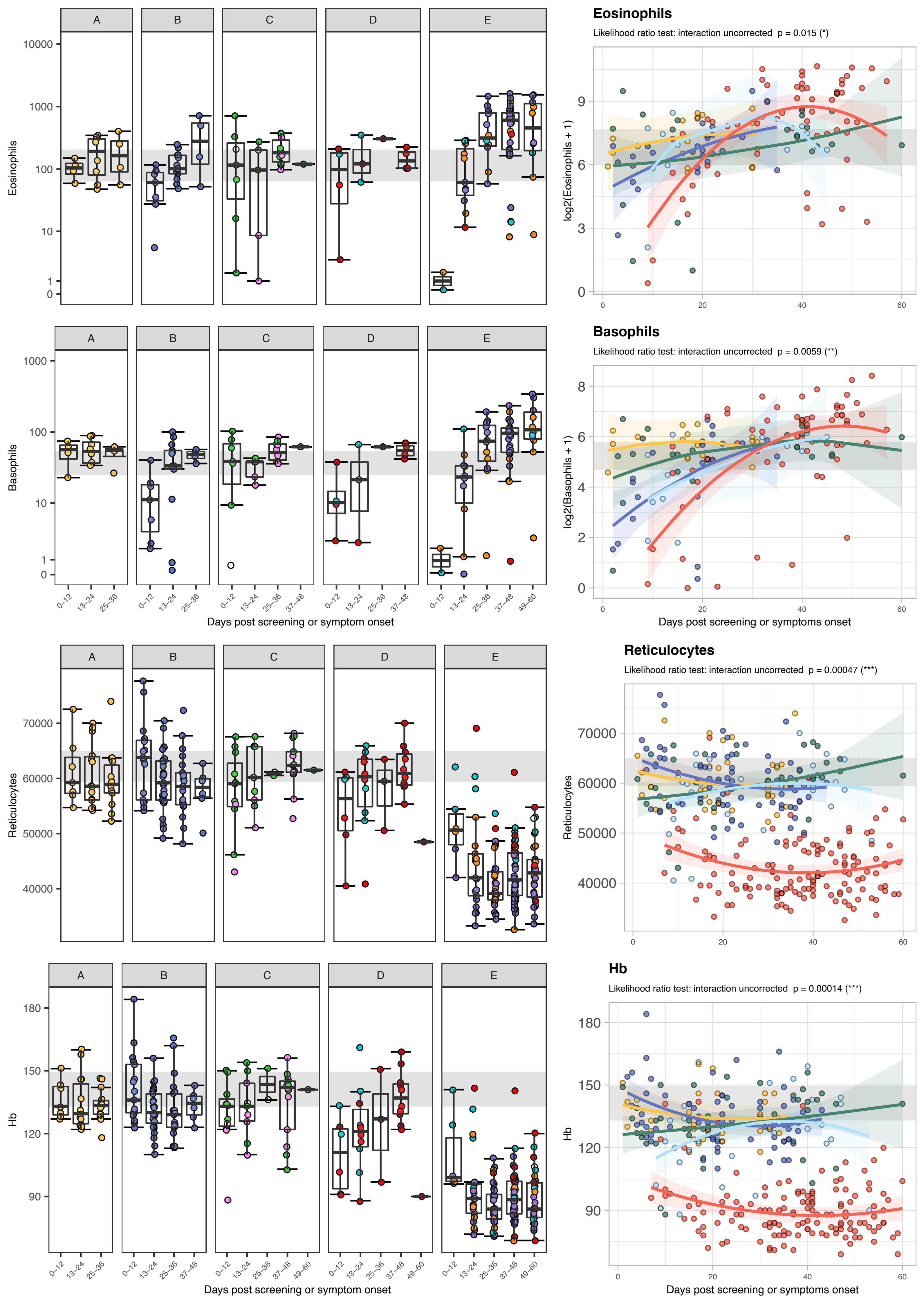
Eosinophils
Likelihood ratio test: interaction uncorrected p = 0.015 (*)
log2(Eosinophils + 1)
Basophils
Likelihood ratio test: interaction uncorrected p = 0.0059 (**)
log2(Basophils + 1)
Days post screening or symptoms onset
Reticulocytes
Likelihood ratio test: interaction uncorrected p = 0.00047 (***)
Hb
Likelihood ratio test: interaction uncorrected p = 0.00014 (***)
Days post screening or symptoms onset
A
B
C
D
E
Eosinophils
Basophils
Days post screening or symptom onset
Reticulocytes
Hb
Days post screening or symptom onset
0–12
13–24
25–36
37–48
49–60
