## Supplementary figures and images for "Delayed bystander CD8 T cell activation, early immune pathology and persistent dysregulation characterise severe COVID-19"

### Item_S3_IMMUNITY_15032021.pdf

##### Item S3: Differences between patients in cluster 1 and cluster 2 defined as figure 4A

A

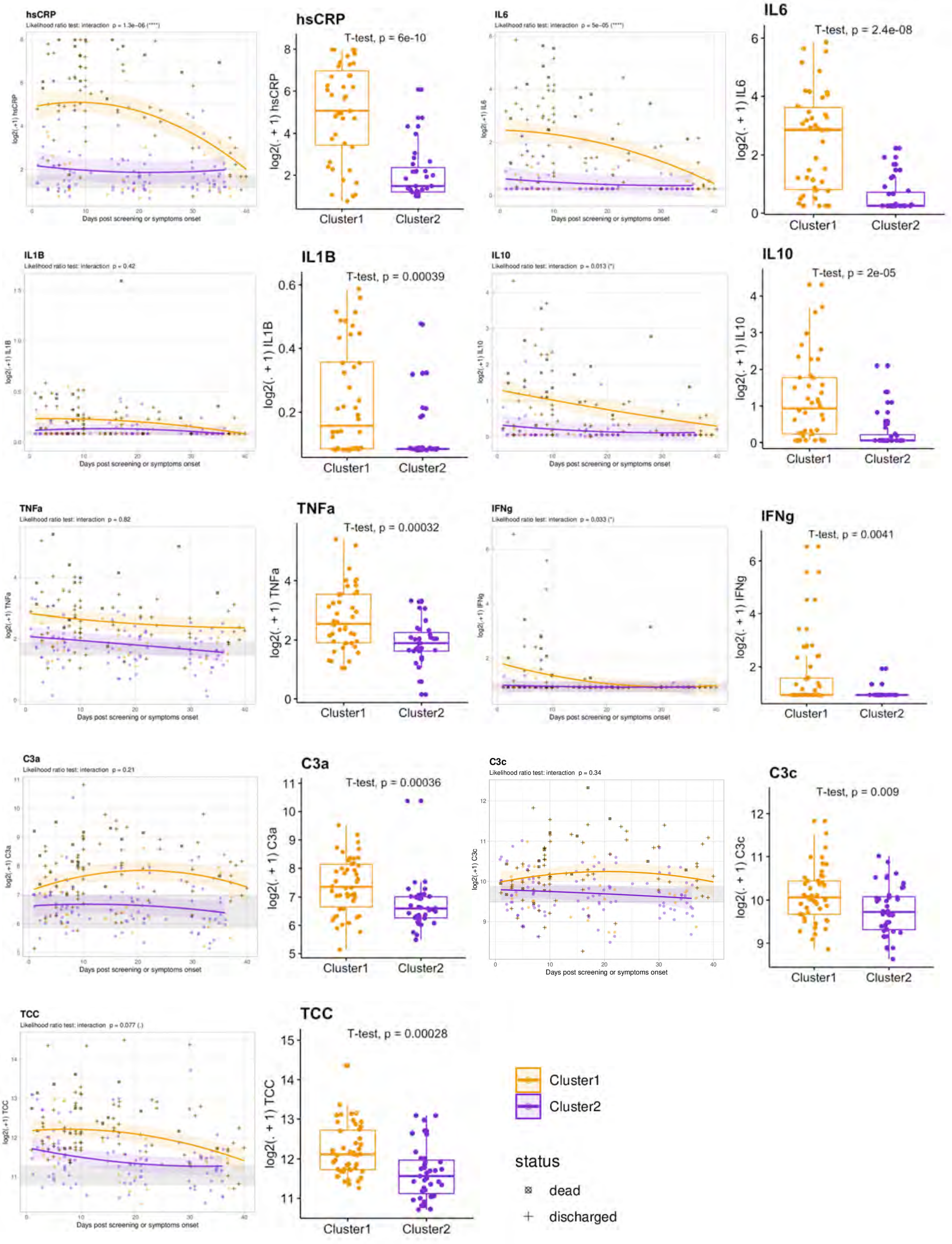

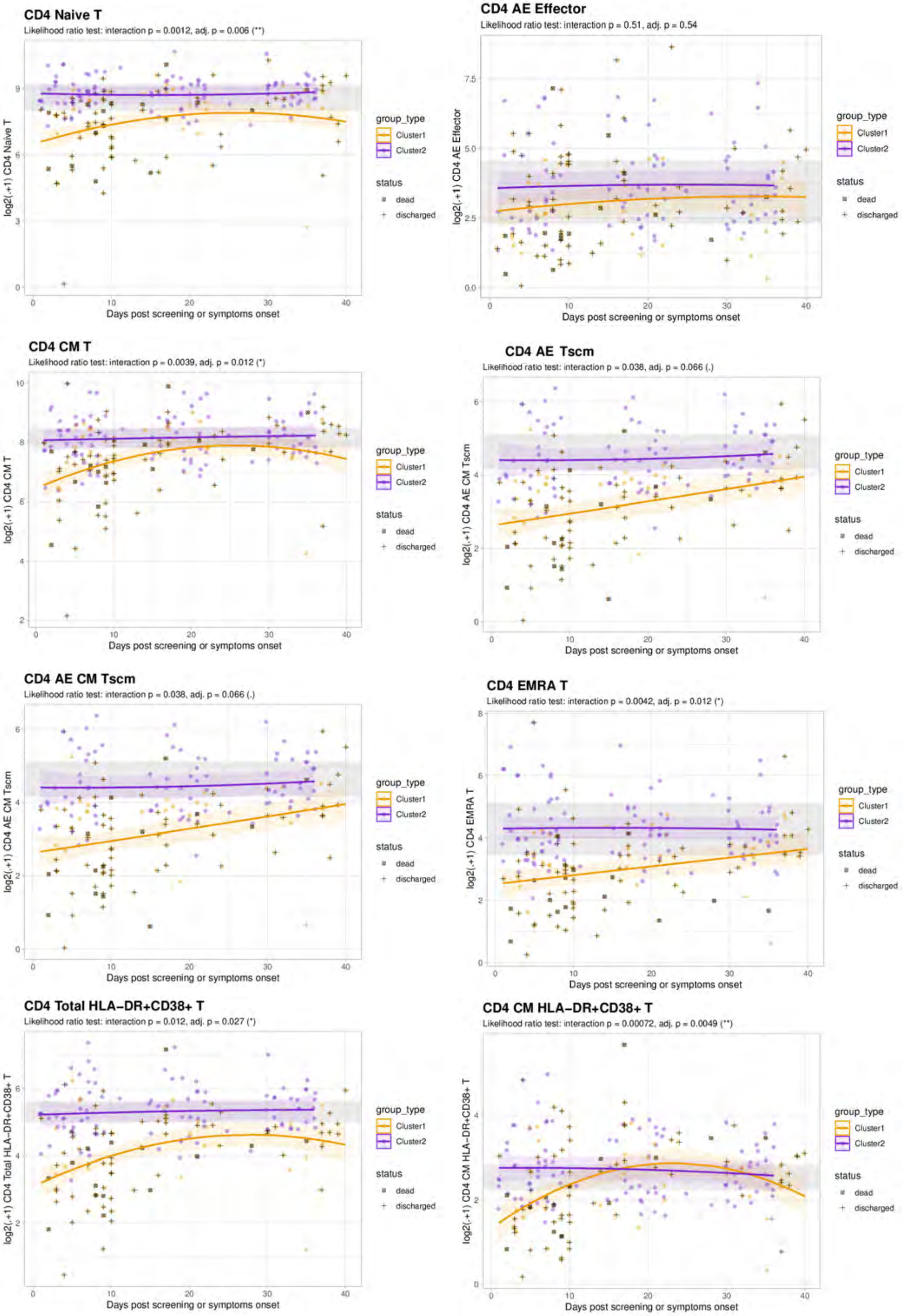

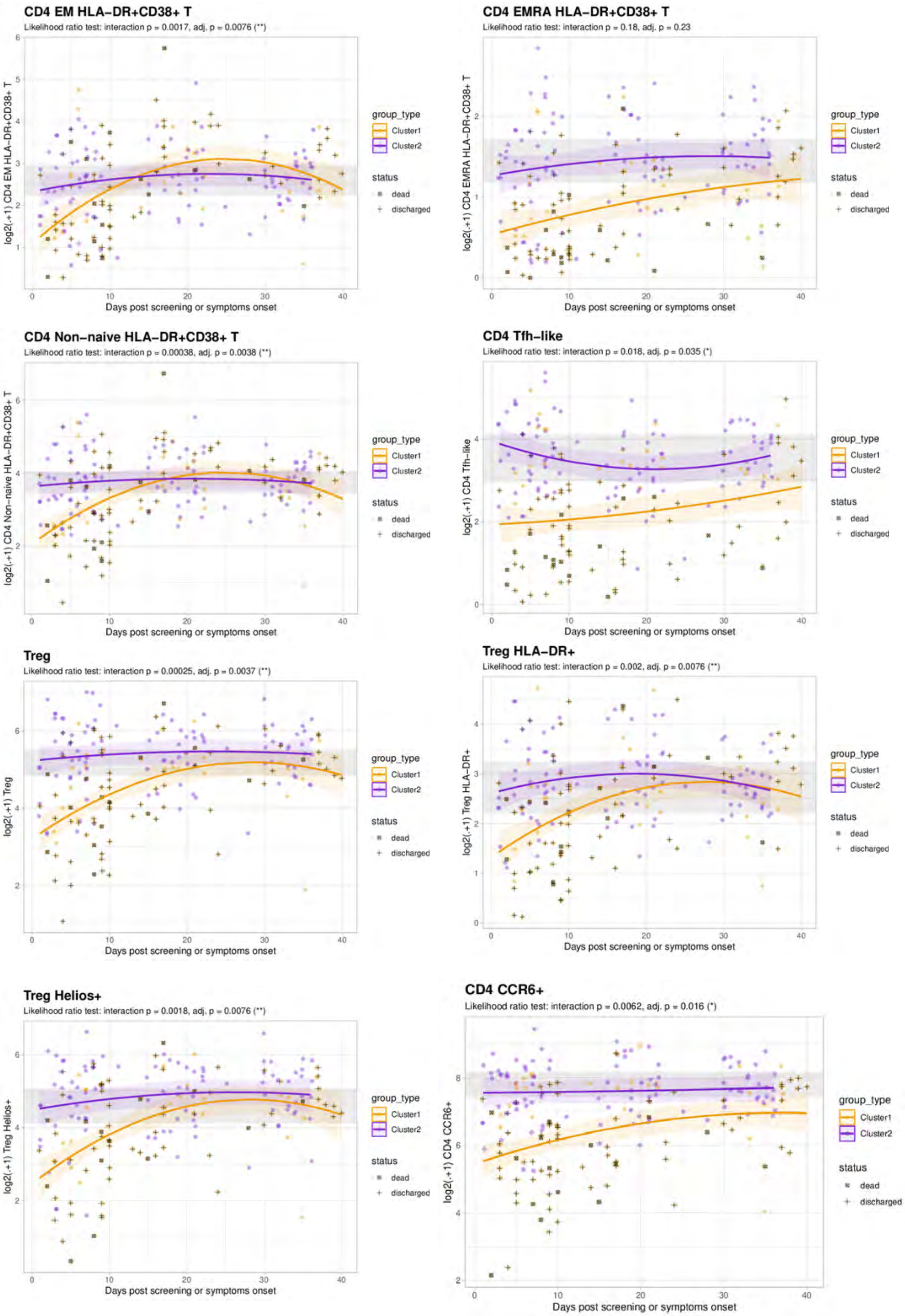

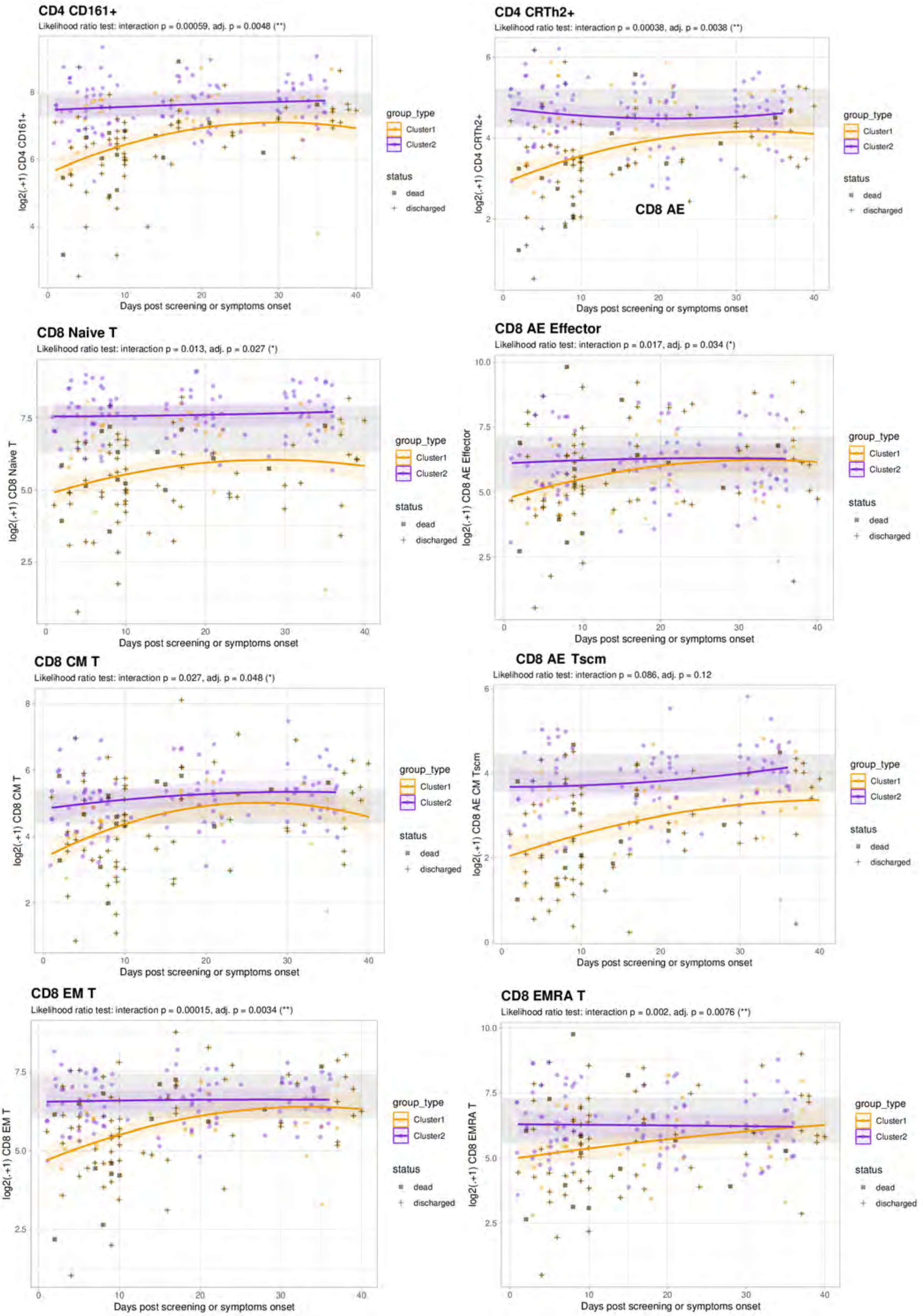

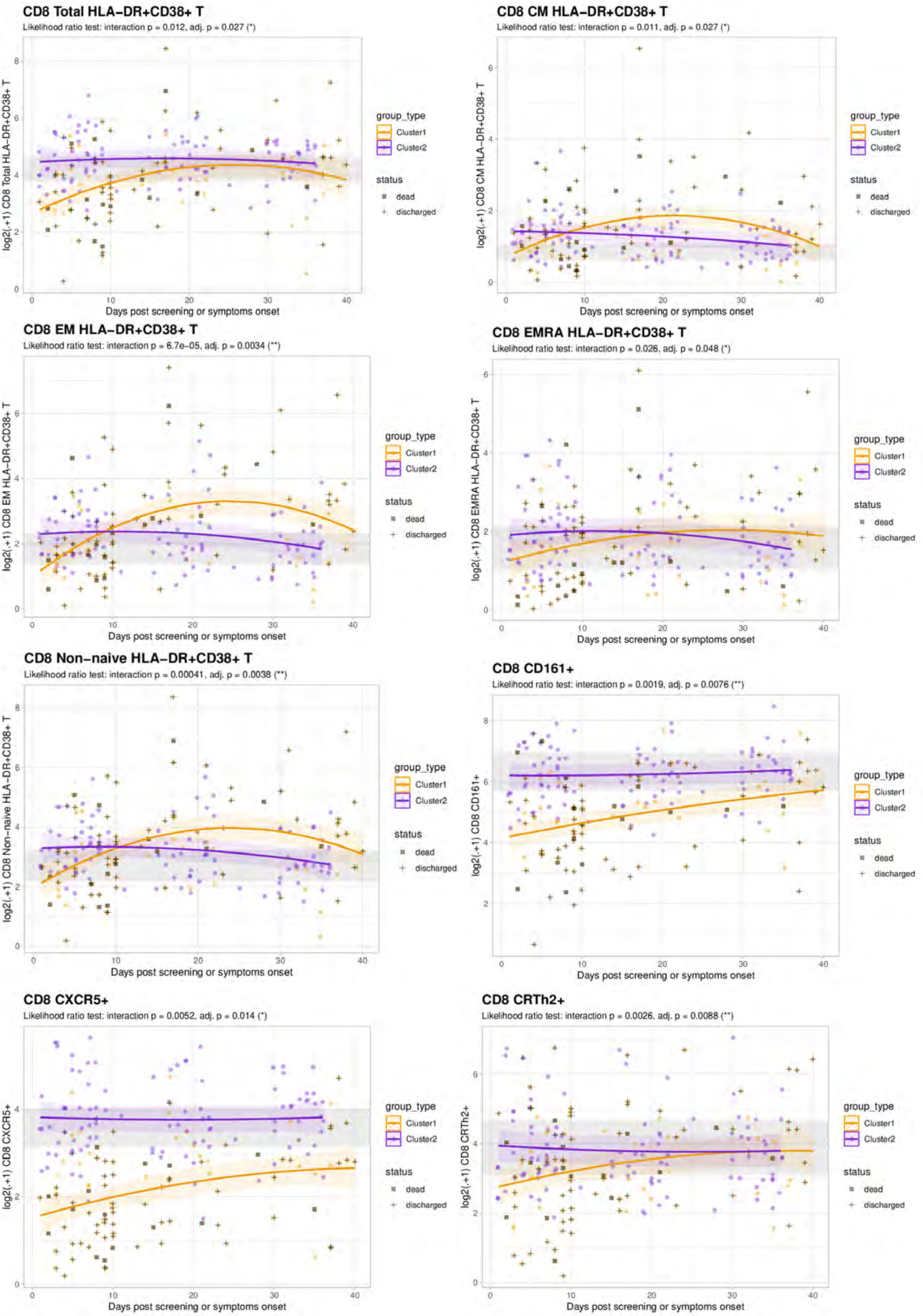

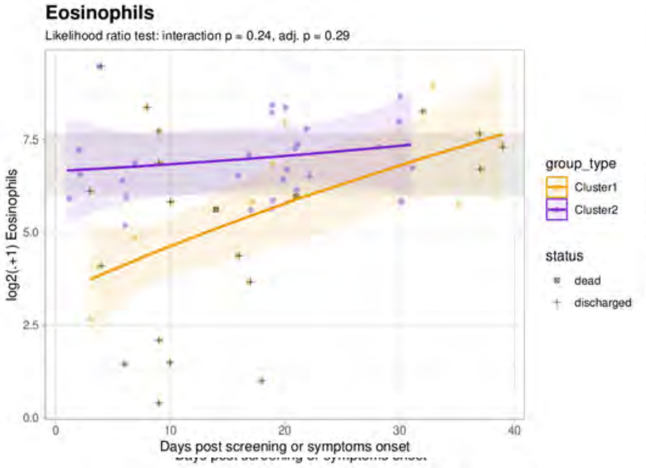

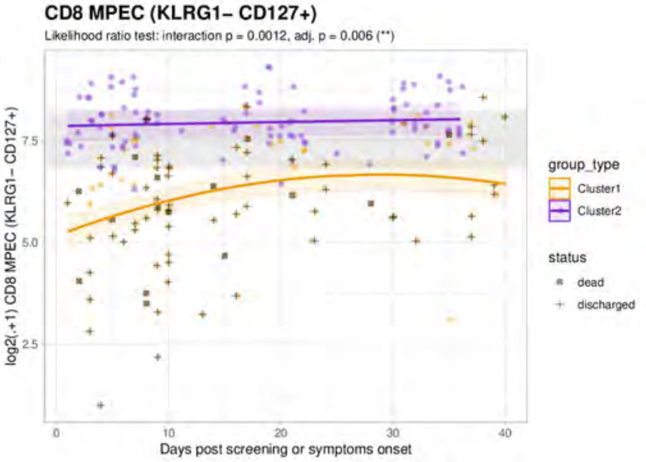

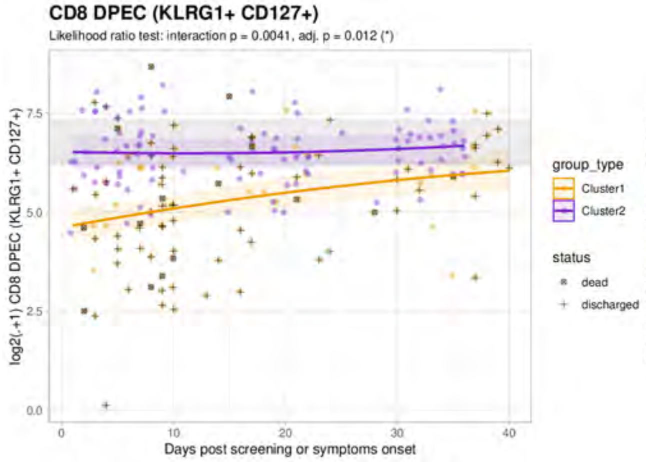

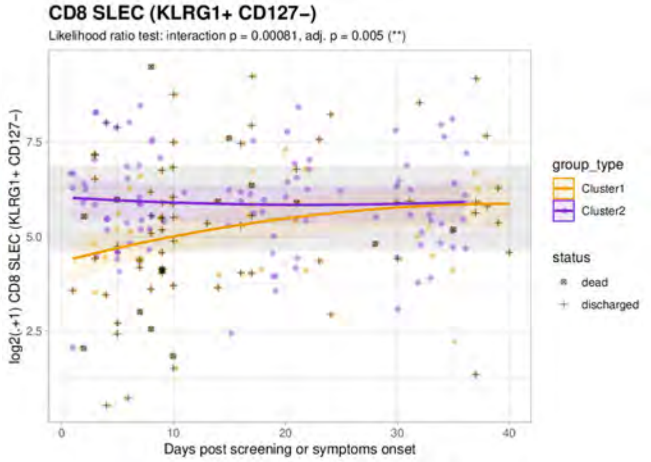

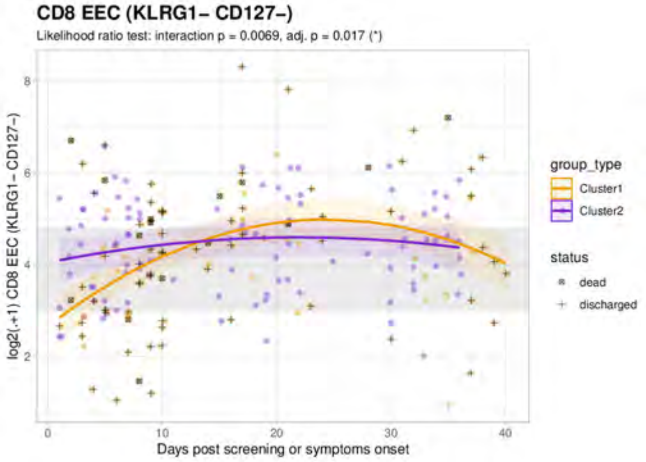

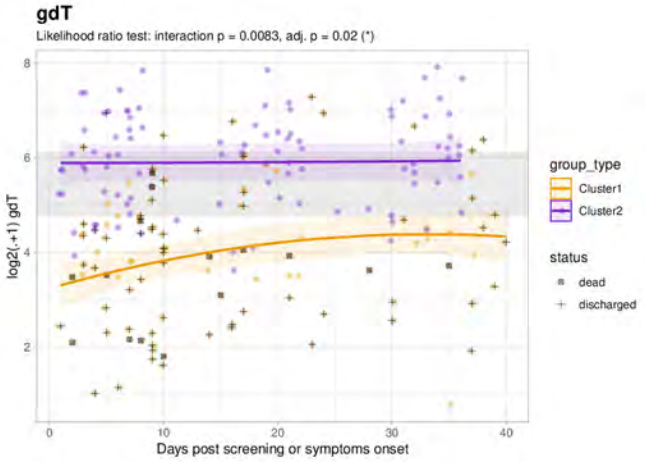

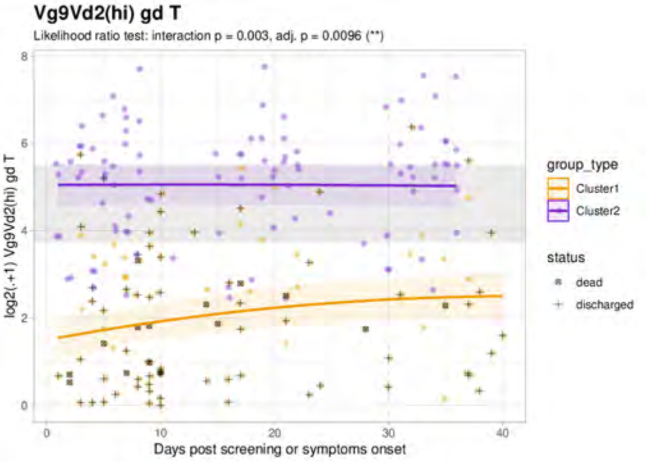

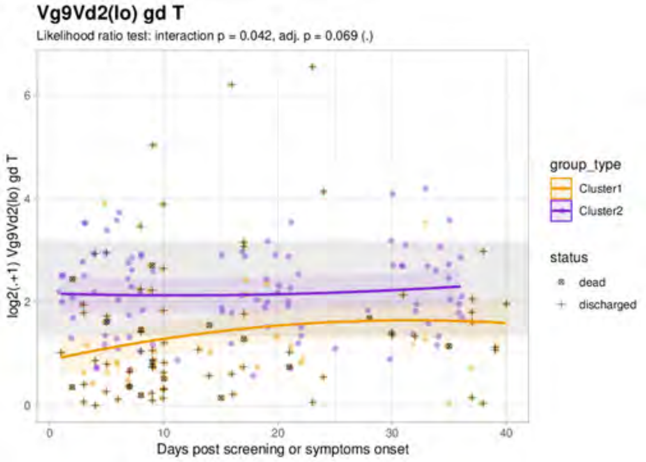

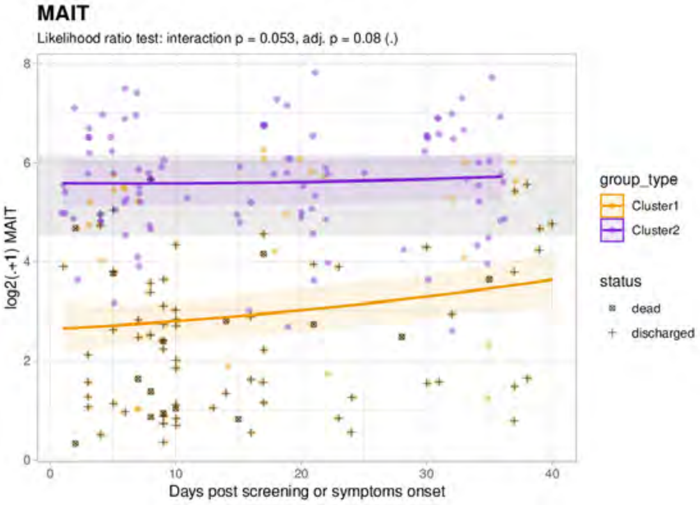

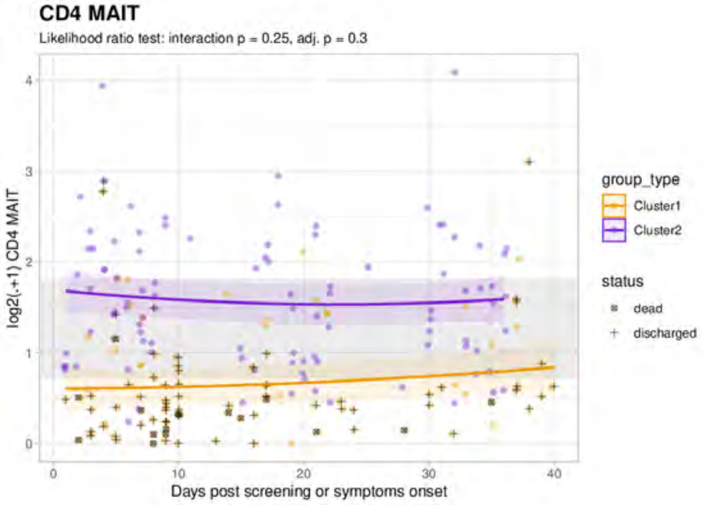

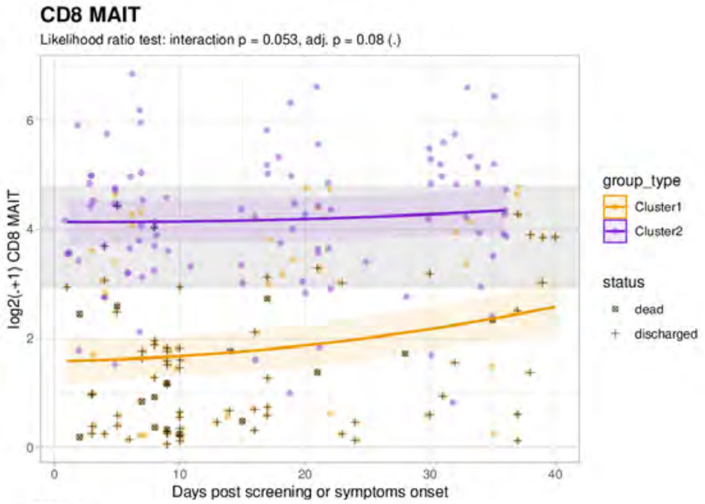

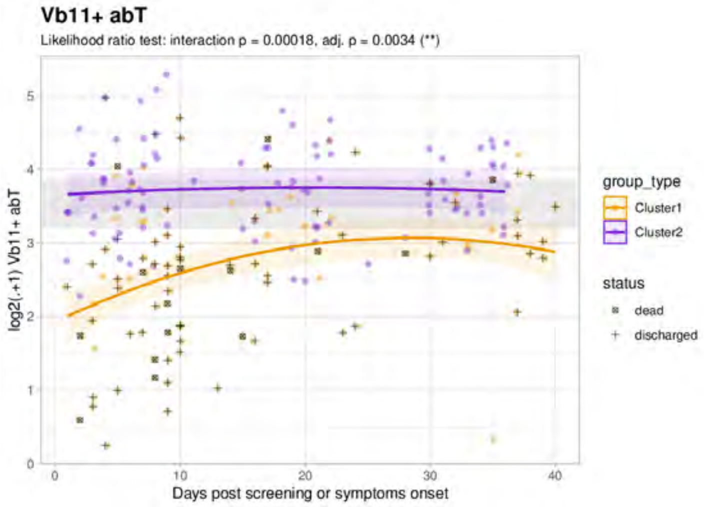

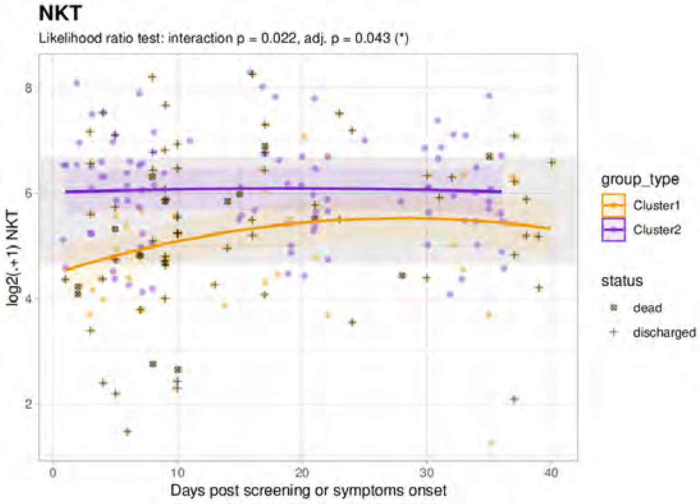

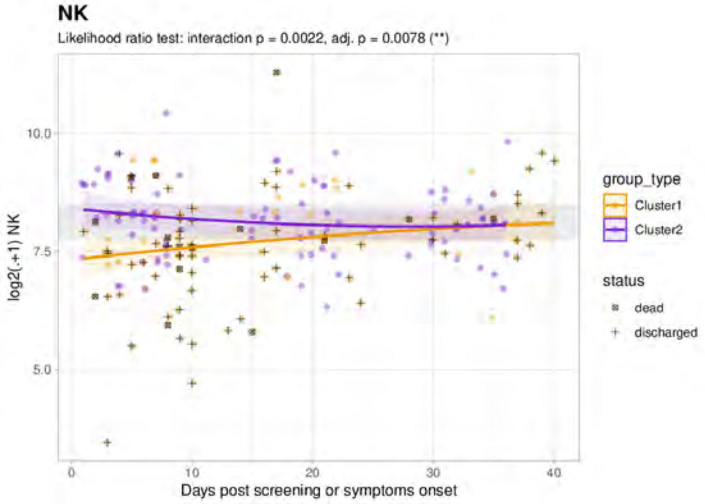

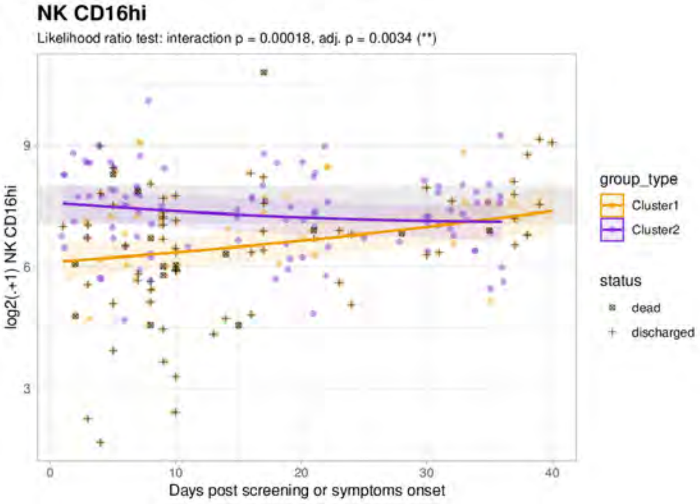

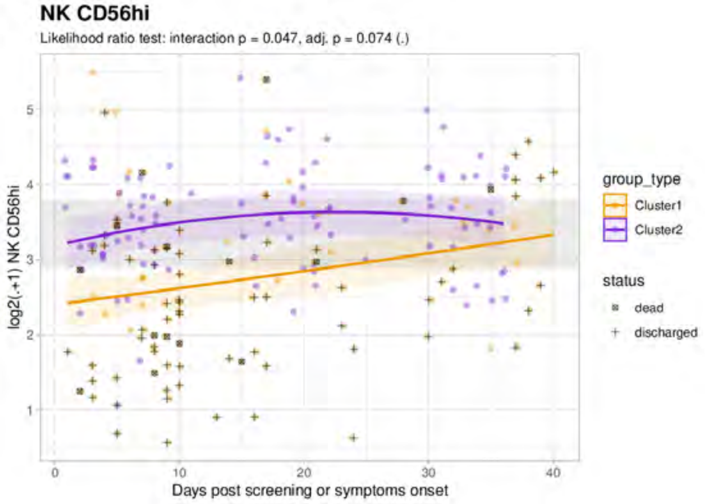

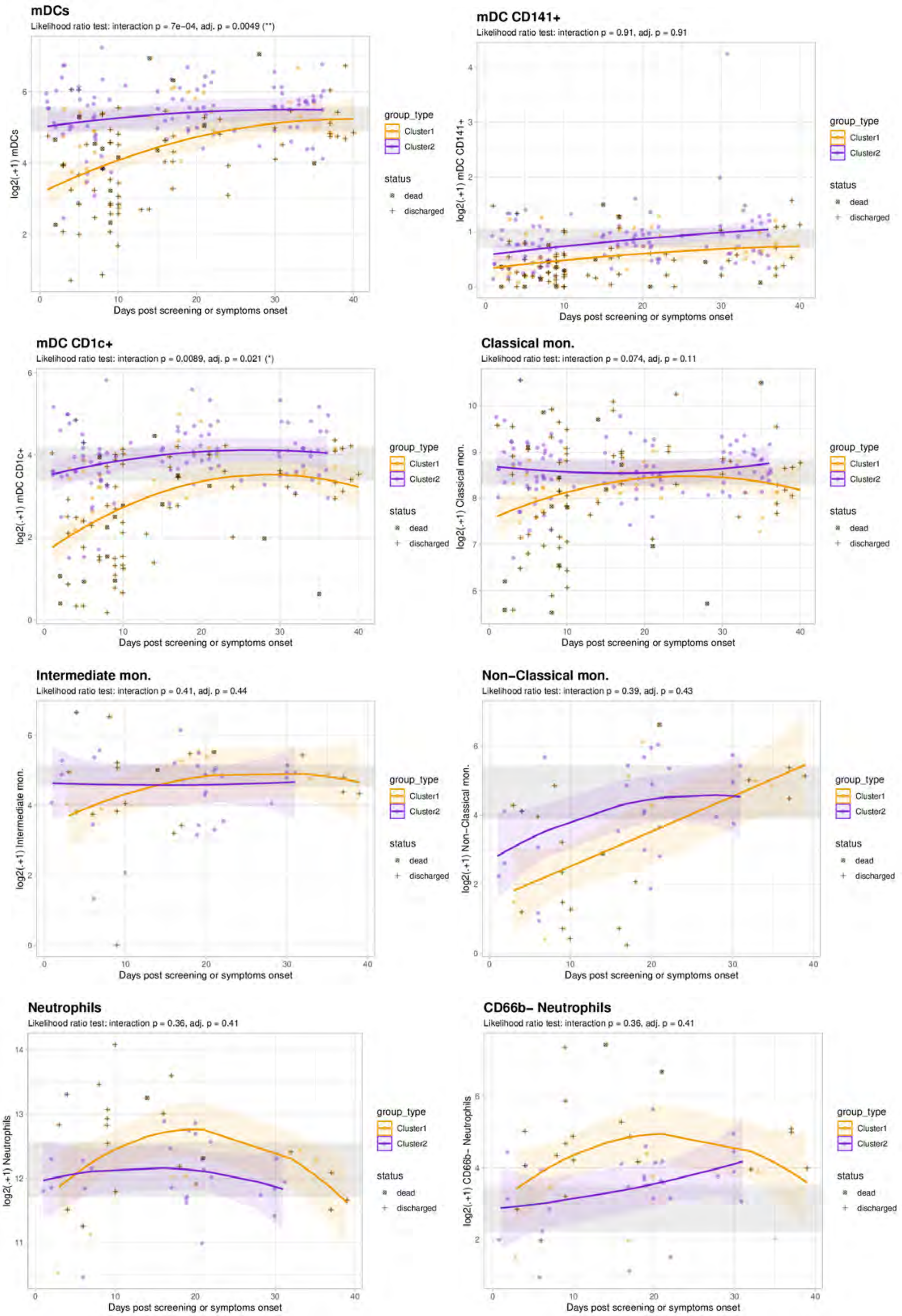

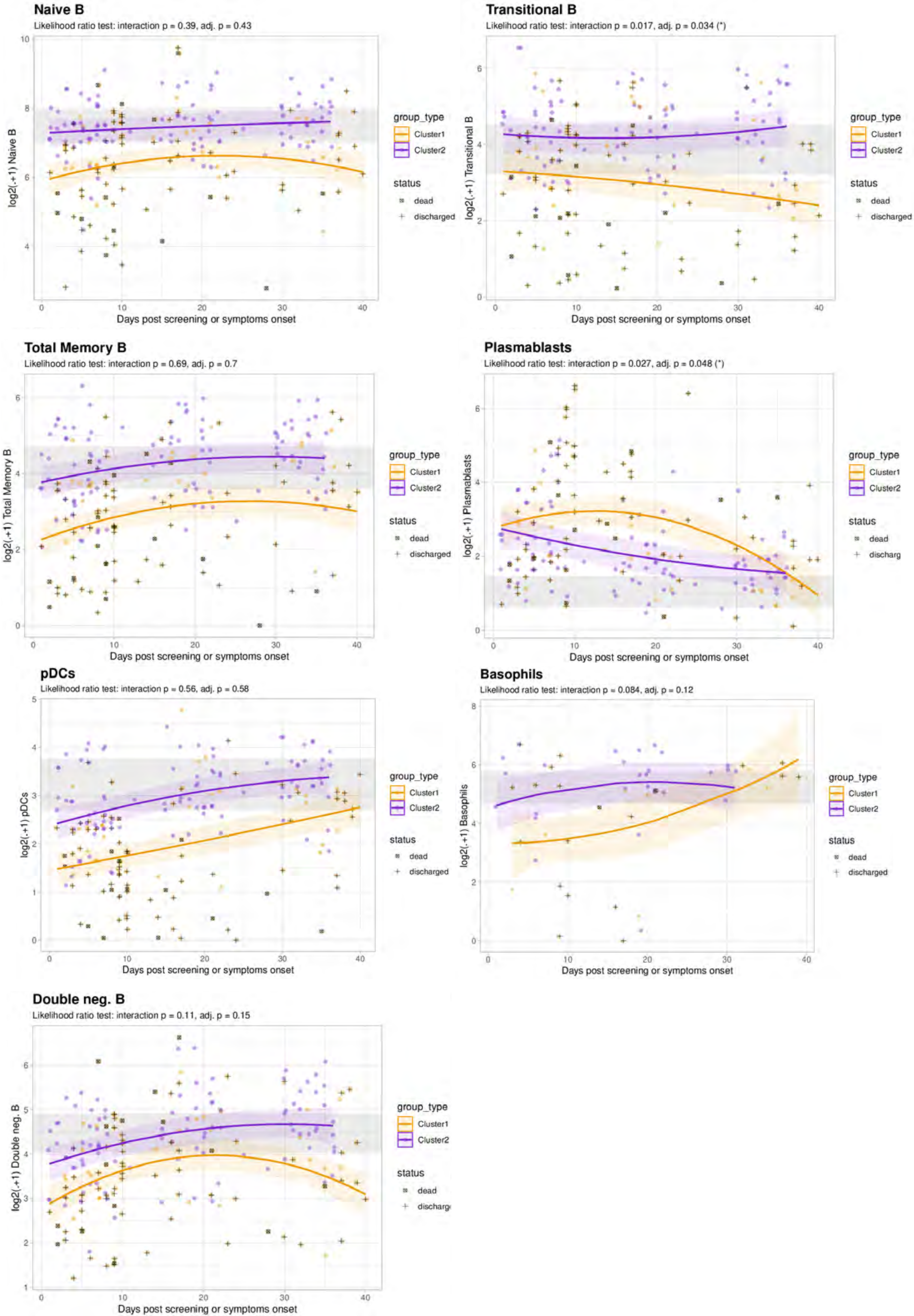

### Item_S4_IMMUNITY_15032021.pdf

#### Item S4: Flow cytometry gating strategy

A

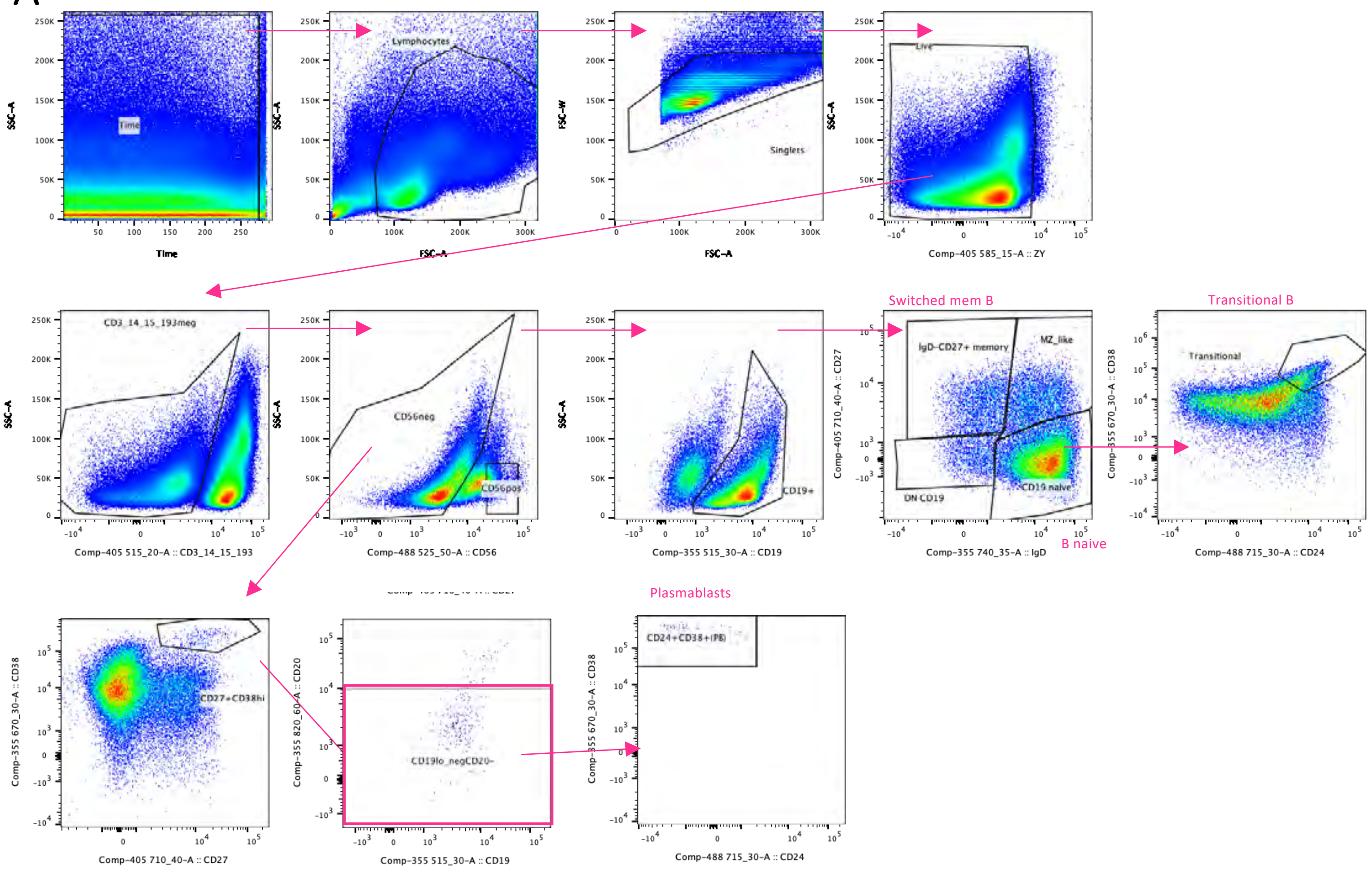


B

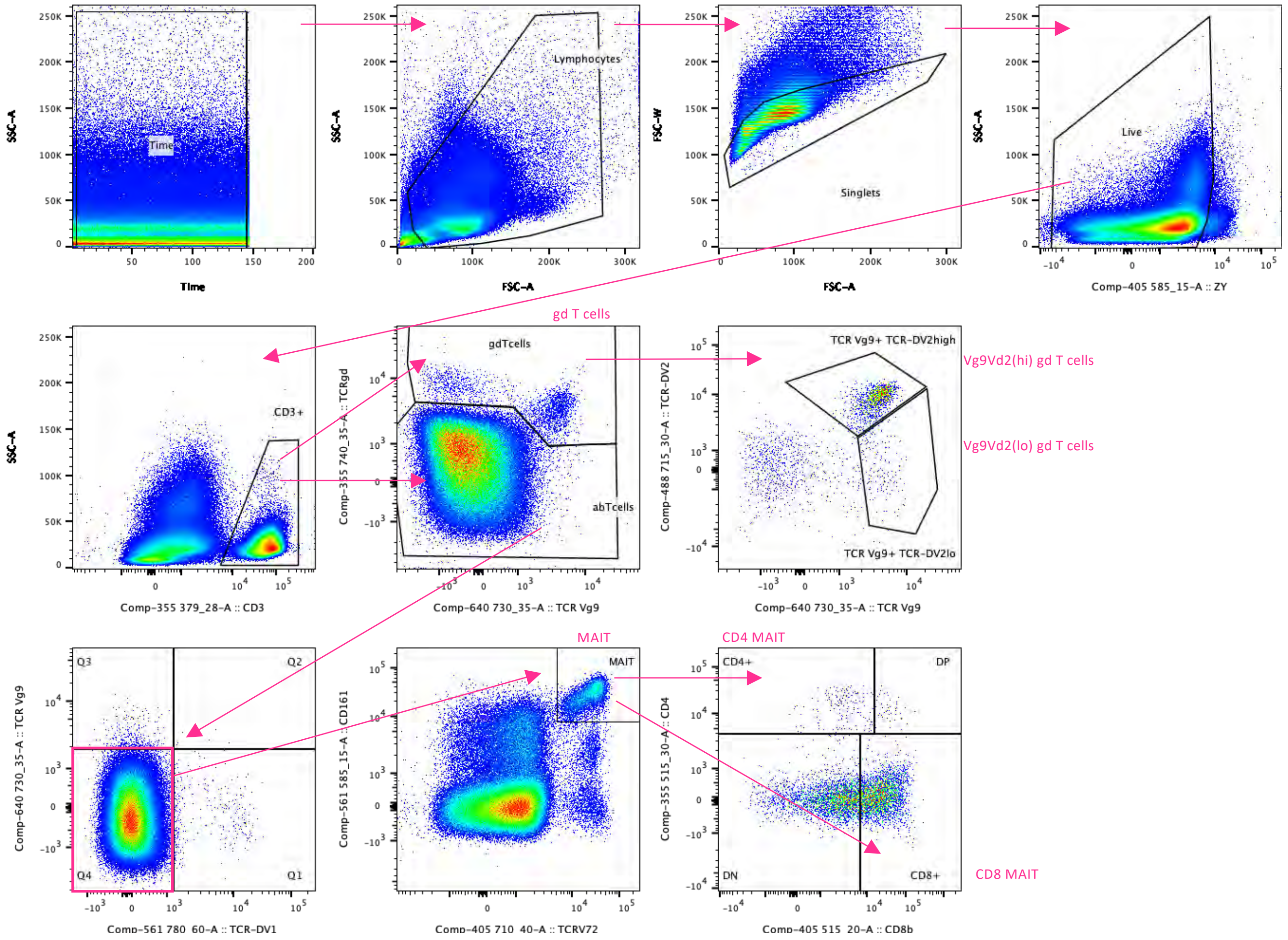

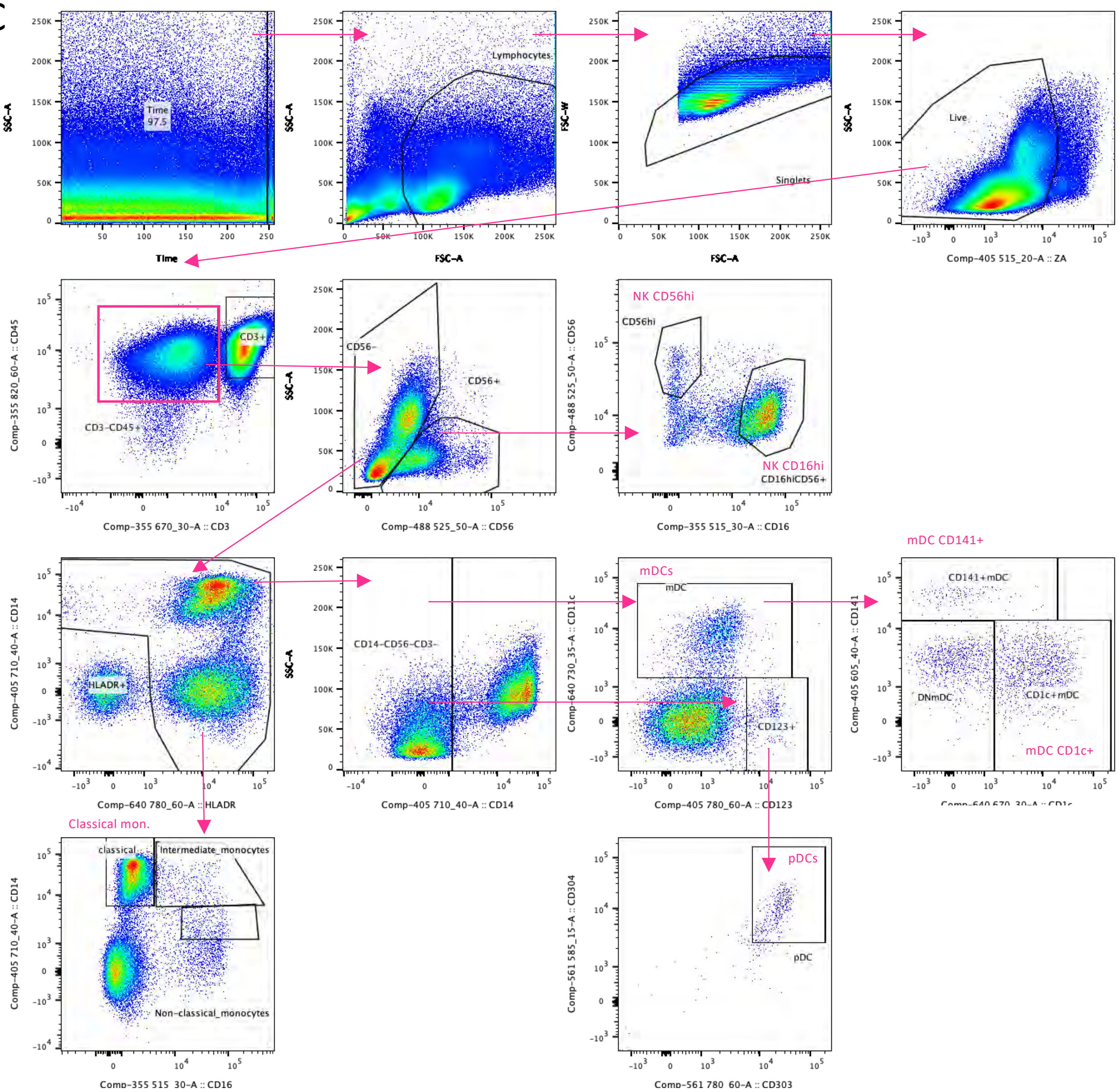

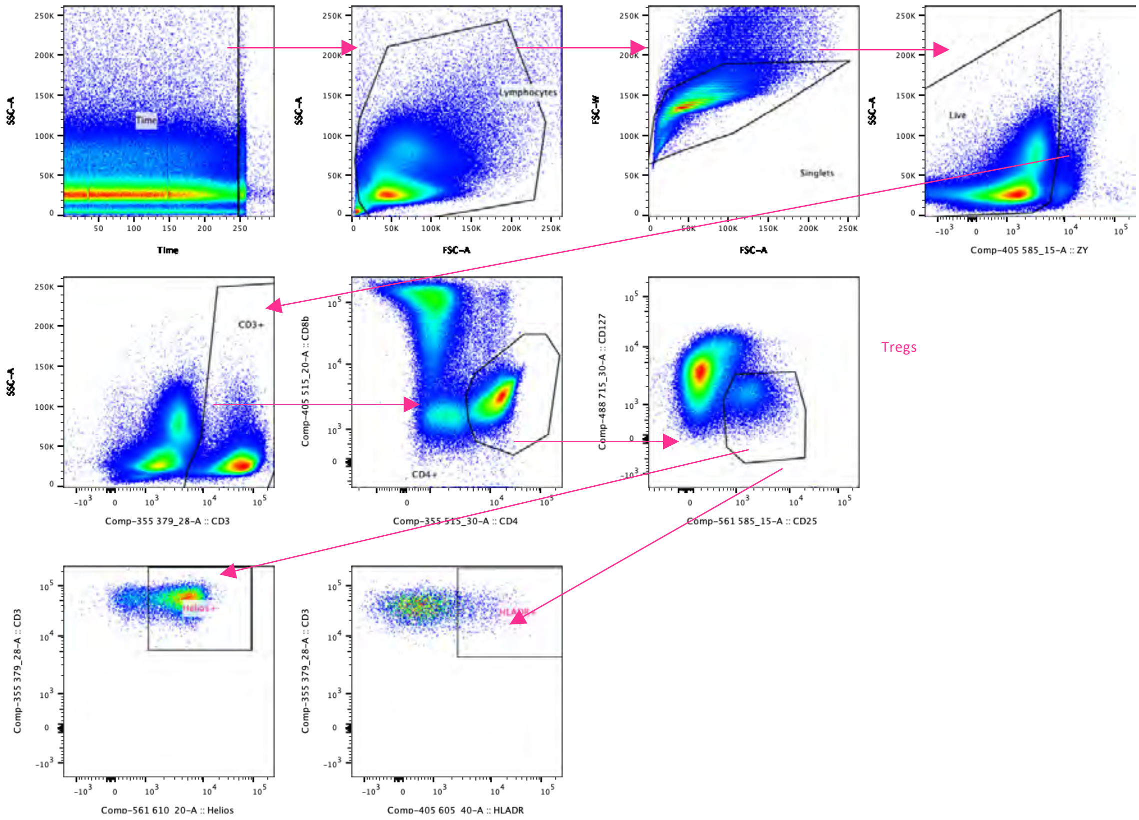

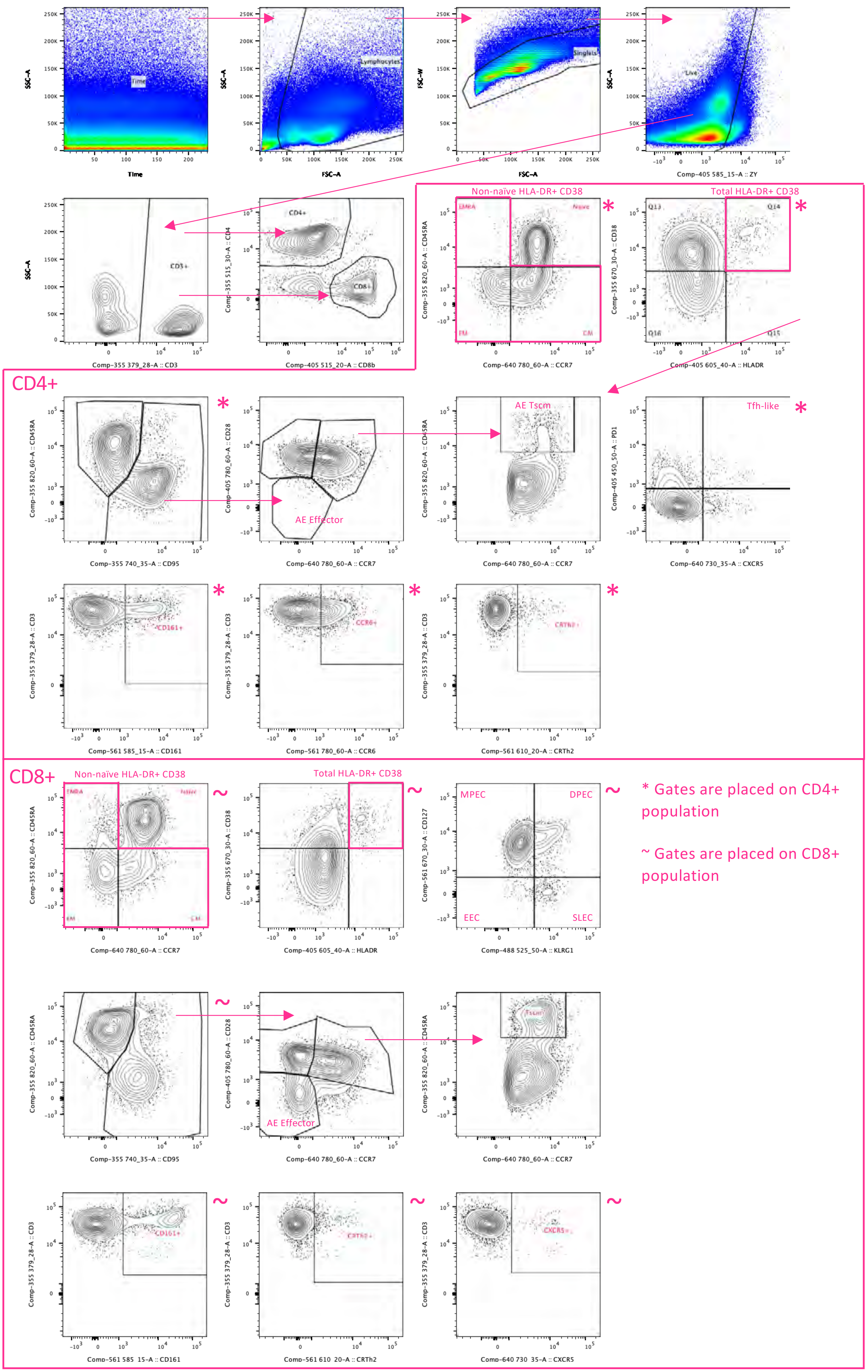
